# The landscape of plasma proteomic links to human organ imaging

**DOI:** 10.1101/2025.01.14.25320532

**Authors:** Zirui Fan, Julio Chirinos, Xiaochen Yang, Juan Shu, Yujue Li, Joan M. O’Brien, Walter Witschey, Daniel J. Rader, Ruben Gur, Bingxin Zhao

## Abstract

Plasma protein levels provide important insights into human disease, yet a comprehensive assessment of plasma proteomics across organs is lacking. Using large-scale multimodal data from the UK Biobank, we integrated plasma proteomics with organ imaging to map their phenotypic and genetic links, analyzing 2,923 proteins and 1,051 imaging traits across multiple organs. We uncovered 5,067 phenotypic protein-imaging associations, identifying both organ-specific and organ-shared proteomic relations, along with their enriched protein-protein interaction networks and biological pathways. By integrating external gene expression data, we observed that plasma proteins associated with the brain, liver, lung, pancreas, and spleen tended to be primarily produced in the corresponding organs, while proteins associated with the heart, body fat, and skeletal muscle were predominantly expressed in the liver. We also mapped key protein predictors of organ structures and showed the effective stratification capability of plasma protein-based prediction models. Furthermore, we identified 8,116 genetic-root putative causal links between proteins and imaging traits across multiple organs. Our study presents the most comprehensive pan-organ imaging proteomics map, bridging molecular and structural biology and offering a valuable resource to contextualize the complex roles of molecular pathways underlying plasma proteomics in organ systems.

## Introduction

Proteins, as the functional outputs of genes, play essential roles in biological mechanisms. Advances in high-throughput proteomics technologies, such as the Olink^1^ and SomaScan^2^ platforms now enable the profiling of thousands of plasma proteins in biobank-scale cohorts^3–9^, providing unprecedented insights into health^4,10^, aging^11–15^, disease^16,17^, and drug discovery^18^. Plasma proteins, originating from tissues across the body^13^, perform context-specific molecular roles within each organ^19^. Mapping the locations of these proteomic links is essential to understanding the organ-specific pathways and mechanisms through which plasma proteins relate to health and clinical outcomes. For example, brain structural phenotypes have been shown to mediate the effects of plasma levels of BCAN, NCAN, and MOG on cognitive ability^20^. Infection-related proteins (such as PIK3CG, PACSIN2, and PRKCB) may contribute to neurodegeneration and Alzheimer’s disease through region-specific brain volume loss^21^. However, a comprehensive, integrated view of plasma protein links across organs is lacking, limiting our ability to fully contextualize their roles in health and disease.

Medical imaging, such as magnetic resonance imaging (MRI), provides non-invasive measures of organ structure and function^22–26^. Imaging-derived phenotypes (IDPs), which capture variations in tissue composition, organ morphology, and functional activity/connectivity, have been extensively linked to physiological and pathological processes in a wide range of diseases, such as heart failure^27^, chronic liver diseases^28^, Alzheimer’s disease^29^, and glaucoma^30^. Integrating IDPs with plasma proteomics holds significant potential for advancing our understanding of organ-specific proteomic biology^31^. However, few large-scale studies have collected both multi-organ imaging and plasma protein data within the same cohort. Similar to many omics mapping strategies of complex traits and diseases^32–34^, such data constraints have led existing studies to rely on separate cohorts for imaging and (prote)omics data^35^, limiting analyses solely to genetics-driven associations^36,37^. Genetic-based, separate-cohort approaches are known to face challenges^35^, including demographic mismatches between cohorts, which can introduce bias, and limited power to detect proteomic effects influenced by non-genetic factor, which limits the insight into mechanisms. Moreover, existing separate-cohort studies have typically focused on single-organ or single-modality IDPs^21,36,37^. These limitations underscore the need for large-scale, single-cohort datasets that directly integrate multi-organ imaging and proteomic data to comprehensively investigate the biological roles of proteins in each organ^31^.

Leveraging multi-organ imaging^26^ and plasma protein^4^ data from the UK Biobank (UKB), we conducted the largest pan-organ imaging proteomics analysis to date. We mapped proteomic links across 2,923 Olink plasma proteins and 1,051 IDPs, encompassing a wide range of organs and tissues, including the brain, heart, aorta, liver, kidney, lung, pancreas, spleen, body fat, and muscle composition (average *n* = 4,896 participants with both imaging and protein data). We developed an atlas of phenotypic protein-imaging associations, revealing organ-specific proteomic networks and enriched biological pathways. To investigate the origins of these proteomic associations, we integrated external gene expression data^38^ to examine whether proteins may be produced in other organs and act remotely, or originated from the same organ. Additionally, we generated a chart for the predictive ability of plasma proteins on organ structure and function, identifying key protein profiles that strongly predict specific IDPs for each organ. Together with UKB genetic data (average *n* = 40,682 participants for imaging and 34,566 for protein after removing overlapping imaging subjects), we further evaluated potential genetic causal associations across organs. An overview of the study design is presented in **Figure 1**.

**Fig. 1.**
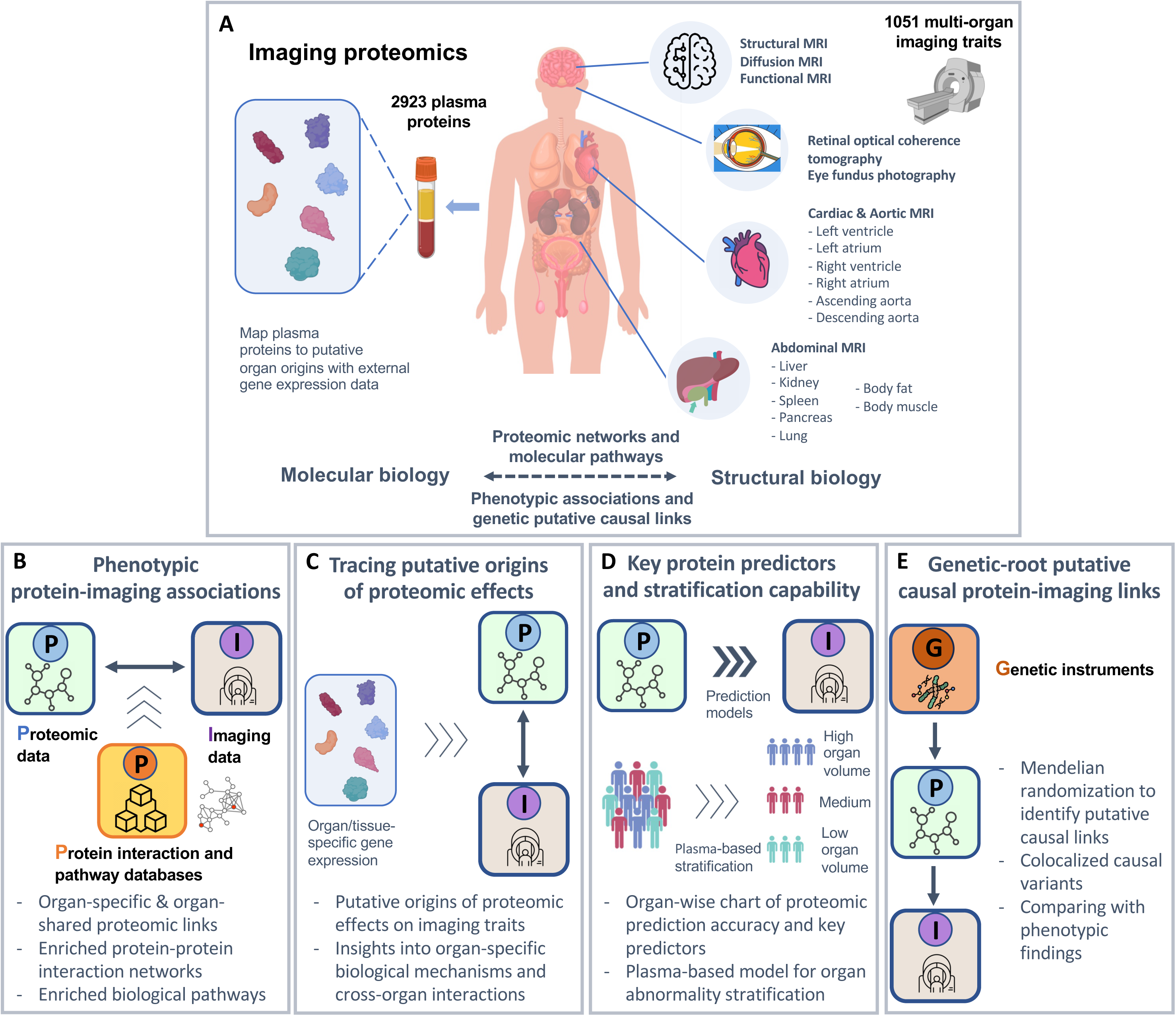
Study overview. **(A)** Our study aimed to explore the phenotypic and genetic connections between plasma proteome and organ structure and function, using 2,923 proteins and 1,051 imaging traits spanning multiple organs from the UK Biobank study. Building on a diverse range of imaging modalities, the study included brain imaging traits such as structural MRI, diffusion MRI, and functional MRI; cardiac MRI traits derived from short-axis, long-axis, and aortic cine images; abdominal MRI traits capturing measurements related to the liver, kidney, lung, pancreas, spleen, and body fat and muscle composition; and eye imaging traits such as derived retinal optical coherence tomography and fundus photography images**. (B)-(E)** Overview of the major analyses, data resources, and key scientific questions addressed in this study.

## RESULTS

### Overview of phenotypic protein-imaging associations

We investigated the phenotypic associations between 2,923 plasma proteins from the UKB pharma proteomics project (UKB-PPP; **Table S1**) and a diverse set of brain and body IDPs. These included 258 brain structural MRI (sMRI) traits, 432 diffusion MRI (dMRI) traits, 82 brain resting-state functional MRI (fMRI) traits, 82 cardiac MRI traits, 41 abdominal MRI traits, as well as 46 optical coherence tomography (OCT) and 110 fundus imaging traits of the eye (**Table S2**). For discovery, we used data from unrelated white British participants (average *n* = 4,383), with effects of age, sex, age-sex interaction, genetic principal components, height, weight, and body mass index removed. Associations were replicated in an independent hold-out sample of white non-British individuals (average *n* = 513; **Methods**). We identified 5,067 associations that were significant in the discovery sample after Bonferroni correction (*P* < 1.63× 10^−8^) and remained nominally significant (*P* < 0.05) in the replication sample with concordant effect signs (**Figs. 2A** and **S1**, **Table S3**). To assess the impact of disease status on protein-imaging associations, we performed sensitivity analyses by additionally adjusting for organ-related diseases (**Methods**). The results showed that protein effect sizes remained highly consistent between models with and without disease status adjustment, with a correlation exceeding 0.99 (**Fig. S2** and **Table S4**).

**Fig. 2.**
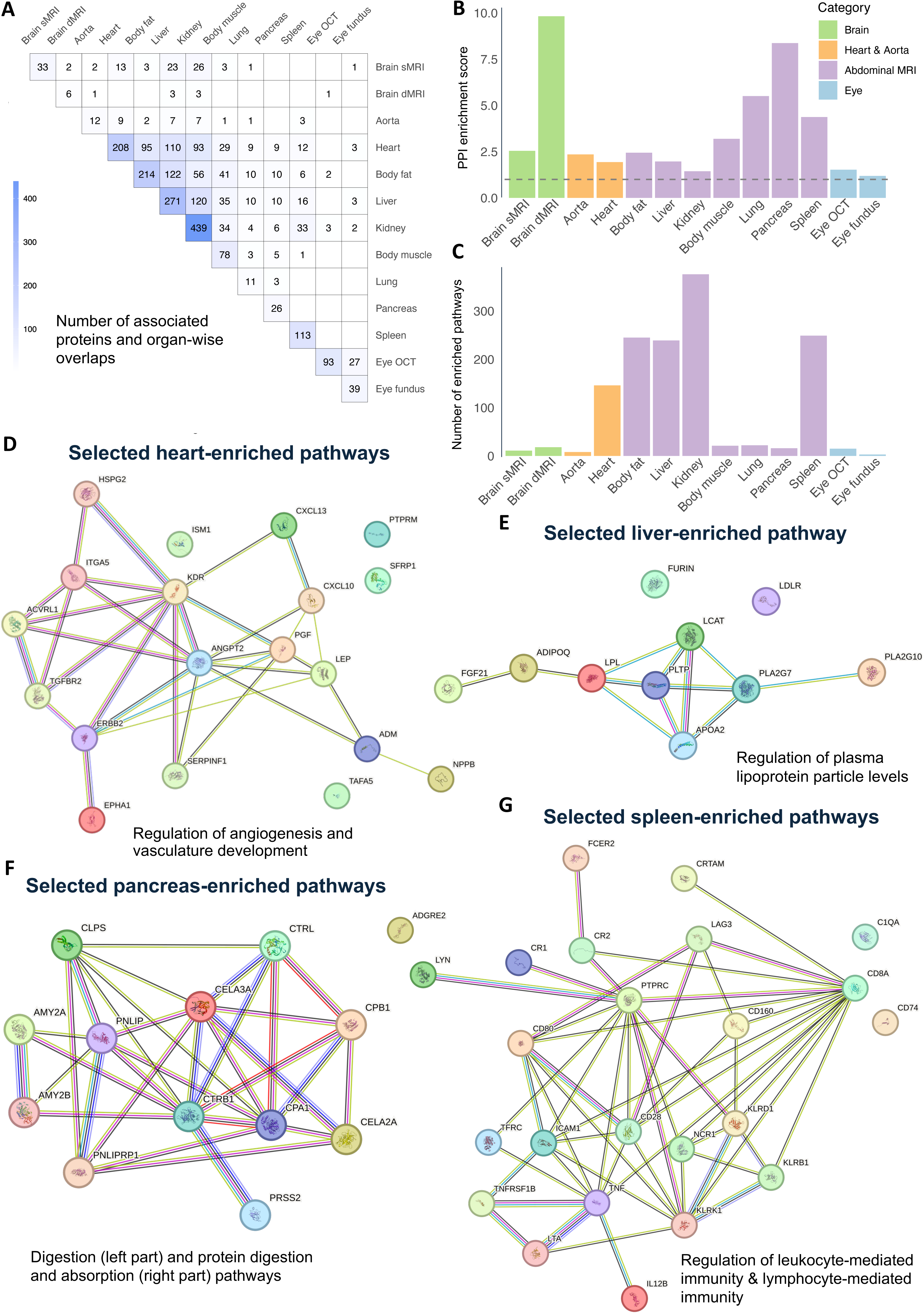
Phenotypic protein-imaging associations and selected enriched pathways. **(A)** The number of significant proteins associated with each organ after the Bonferroni correction (*P* < 1.63× 10^−8^) and the overlapping pattern across organs. **(B)** Enrichment of protein-protein interaction (PPI) scores using the STRING database was assessed. Enrichment scores were calculated for the set of associated proteins for each organ, and statistical significance was tested using a one-sided Wilcoxon rank-sum test. **(C)** Overview of the number of enriched biological pathways in each organ. **(D)** Selected biological pathways enriched among proteins associated with heart imaging traits, including the regulation of angiogenesis and vasculature development (multiple testing-adjusted *P* < 1.53 × 10^−7^). **(E)** Selected biological pathway enriched among proteins associated with liver imaging traits, highlighting the regulation of plasma lipoprotein particle levels pathway (adjusted *P* = 5.36× 10^−5^). **(F)** Selected biological pathways enriched among proteins associated with pancreas imaging traits, including digestion and protein digestion and absorption (adjusted *P* < 1.03 × 10^−7^). **(G)** Selected biological pathways enriched among proteins associated with spleen volume, including lymphocyte mediated immunity and regulation of leukocyte mediated immunity (adjusted *P* < 6.67× 10^−15^).

We found that plasma proteins associated with organ structure and function tended to exhibit high levels of interaction, with significant enrichment observed for all organs using the STRING protein-protein interaction (PPI) database^39^ (*P* < 7.31 × 10^−3^, **Fig. 2B** and **Table S5**). Since proteins often function collaboratively within biological pathways, these findings suggest the presence of proteomic modules and associated network biology within each organ. Further pathway enrichment analysis identified 1,429 enriched biological pathways^40^ (**Fig. 2C** and **Table S6**) after multiple testing adjustments (**Methods**). Some protein clusters and pathways had highly organ-specific associations, while others suggested cross-organ links. In the following sections, we highlight key proteins associated with each organ, along with the proteomic interaction networks and biological pathways they are involved in.

### Plasma proteomic links and enriched biological pathways in the human heart

We observed numerous connections between plasma proteins and body IDPs, spanning the heart, liver, body fat, and kidney (**Fig. S3**). Specifically, we identified 208 proteins associated with cardiac MRI traits of the heart, particularly end-diastolic volume, end-systolic volume, and stroke volume of left/right ventricles (|β| range = (0.07, 0.26), *P* < 1.52 × 10^−8^). Most (over 94%) protein-heart associations were negative, except few proteins such as IGFBP1, IGFBP2, leptin (LEP), and NTPROBNP. These positively associated proteins often serve as markers of adverse cardiac conditions. For example, NTPROBNP is a well-known biomarker associated with increased cardiac filling pressures, the presence of heart failure as well as outcomes in established heart failure^41^. Additionally, LEP promotes oxidative stress, inflammation, and atherogenesis, linking elevated levels to cardiovascular diseases such as coronary artery disease, stroke, and diabetes-related complications^42,43^. Many of the identified heart-associated plasma proteins have well-known biological functions on the heart. For example, GDF15 may protect the heart by activating SMAD2/3 signaling to attenuate hypertrophy and preserve ventricular function under pathological conditions^44^. Plasma levels of ICAM1 and SELE were elevated in individuals with coronary heart disease (CHD) and carotid artery atherosclerosis, independently predicting the risk of both conditions, making them potential molecular markers for atherosclerosis and CHD development^45^.

Moreover, proteins associated with the heart were enriched in various biological pathways. For example, 19 heart-associated proteins were significantly enriched in regulation of angiogenesis and vasculature development (multiple testing-adjusted^40^ *P* range = (1.53× 10^−7^, 1.14× 10^−7^), **Fig. 2D**). Many proteins were growth factors (such as PGF, ADM, ANGPT2) or growth factor receptors (such as KDR, ERBB2, TGFBR2, ACVRL1), which played key roles in regulating vascular development^46–49^. There are other heart-associated proteins that were not growth factors but were also critical in angiogenesis. For example, EPHA1 promotes tumor angiogenesis by regulating endothelial tubulogenesis and recruiting endothelial progenitor cells, with its inhibition reducing tumor angiogenesis and growth^50^. Given the complex and multifaceted role of angiogenesis in cardiovascular diseases, these identified proteins enriched in the angiogenesis pathways could serve as biomarkers of disease state or context-specific therapeutic targets^51,52^. The associated cardiac MRI traits provide valuable insights into their underlying biologic pathways and mechanisms. Furthermore, heart-associated proteins were also enriched in the movement of cells or organisms in response to chemical stimuli, such as taxis, chemotaxis, and positive regulation of leukocyte migration (adjusted *P* range = (5.89 × 10^−7^, 3.06 × 10^−7^), **Figs. S4-S5**). A large proportion of proteins exclusively enriched in taxis and chemotaxis were chemokines and cytokines, including CXCL10, CXCL13, CCL22, and CCL27. These proteins recruit immune cells and play critical roles in inflammatory, infectious and immune responses^53–56^ and are promising therapeutic targets for multiple cancer types.

We identified 12 proteins associated with cardiac MRI traits of the aorta (|β| range = (0.07, 0.14), *P* < 1.49× 10^−8^, **Fig. S6**), primarily linked to the descending aorta. Of these, nine proteins overlapped with those associated with the heart, and most of the aorta-protein associations were also negative. Positive associations were only found with two proteins IGFBP1 and Renin (REN) (β range = (0.07, 0.08), *P* < 1.19× 10^−8^). REN, encoded by *REN*, is secreted by the kidney and initiates the renin-angiotensin-aldosterone system cascade, regulating blood pressure and volume. Dysregulation of this system contributes to cardiovascular and renal disorders, making renin an important therapeutic target^57^.

### Shared proteomic associations and systemic processes in abdominal organs

In addition to the heart, we found many associations between plasma proteins and IDPs of the liver, body fat, and kidney, with these four organs/tissues showing the highest proportion of overlapping associated proteins among all the organs examined (**Fig. 2A**). Plasma proteins shared by these organs widely participate in systemic processes such as metabolic regulation, inflammation, and vascular health, highlighting potential targets for understanding systemic diseases and multi-organ interactions.

We identified 270 proteins associated with the liver (|β| range = (0.08, 0.25), *P* range = (1.61× 10^−8^, 6.62× 10^−67^)), primarily linked to liver volume, fat fraction, and liver iron-corrected T1 (a marker of inflammation and fibrosis). Proteins associated with liver volume differed from those linked to fat fraction, inflammation, and fibrosis, with limited overlap, suggesting that these processes are driven by distinct molecular mechanisms. Liver-associated proteins enriched in many biological pathways. In addition to those shared with heart, such as chemotaxis and leukocyte migration, proteins associated with the liver were also enriched in the regulation of plasma lipoprotein particle levels (adjusted *P* = 5.36 × 10^−5^, **Fig. 2E**) and cholesterol/sterol transport (adjusted *P* = 0.02), underscoring the liver’s central role in lipid metabolism. Many of the involved proteins, such as LCAT, FGF21, APOA2, and FURIN, are produced by the liver and are key regulators of lipid metabolism. For example, LCAT is essential for cholesterol transport, promotes the formation of high-density lipoprotein, potentially reducing atherosclerosis^58^. Reduced LCAT activity has also been observed in individuals with liver disease^59^. FGF21 is critical for maintaining energy balance, regulating glucose and lipid metabolism, and shows promise in treating obesity, type 2 diabetes, and non-alcoholic steatohepatitis^60^. Additionally, liver-associated proteins were uniquely enriched in pathways related to viral processes and the viral life cycle (**Fig. S7**). Many of these proteins act as viral entry receptors or facilitate viral invasion, including ICAM1, a receptor for human rhinovirus^61^; ACE2 and NRP1, which serve as receptors for SARS-CoV-2^62,63^, and FURIN, which cleaves the SARS-CoV-2 spike protein to enable host cell entry^64^. CTSL further supports viral invasion by activating SARS-CoV spike protein-mediated membrane fusion under acidic conditions^65^.

We identified 214 proteins associated with body fat (|β| range = (0.08, 0.37), *P* range = (1.52 × 10^−8^, 8.90 × 10^−160^)), predominantly linked to visceral adipose tissue (VAT) volume and total trunk fat volume. Similar with the liver-associated proteins, body fat-associated proteins were also enriched in biological pathways related to lipid metabolism, including the regulation of plasma lipoprotein particle levels (adjusted *P* = 3.68× 10^−7^), with substantial overlap with liver. Beyond the liver-derived proteins mentioned earlier, additional examples include adiponectin (ADIPOQ) and LPL. ADIPOQ, primarily produced by adipocytes, promotes fatty acid oxidation and improves insulin sensitivity^66^. LPL hydrolyzes triglycerides in chylomicrons and very low-density lipoprotein into fatty acids for cellular uptake, with reduced activity observed in poorly controlled diabetes^67^. These findings suggest the crucial roles of the liver and body fat in systemic lipid regulation and their metabolic interactions with other tissues and diseases. Furthermore, the kidney had the highest number of associations with plasma proteins among all organs, with 439 proteins linked to kidney parenchyma and kidney volume (|β| range = (0.08, 0.38), *P* range = (1.57× 10^−8^, 4.33× 10^−167^)). Nearly all (98.98%, 1,751 out of 1,769 associated pairs) these associations were negative. In addition, 11 proteins were associated with the lung (|β| range = (0.08, 0.17), *P* < 1.53× 10^−8^), many of which were shared with the heart, liver, and body fat as well (**Fig. 2A**).

### Organ-specific proteomic associations with the pancreas and spleen

In contrast to the abdominal organs and tissues with many shared proteomic links, the majority of proteins associated with pancreas and spleen were specific to these two organs (**Fig. 2A**). We observed 26 proteins associated with pancreas fat fraction and volume (|β| range = (0.08, 0.34), *P* range *=* (8.95× 10^−9^, 1.35× 10^−136^), **Fig. S8**). These proteins had a high degree of interaction, with a mean PPI score of 0.22 (enrichment *P* < 2.2 × 10^−16^), and were enriched in digestion-related biological pathways (**Fig. 2F**). Enriched proteins include pancreatic amylases AMY2A and AMY2B, which facilitate carbohydrate breakdown, and lipases such as PNLIP and PNLIPRP1, which digest triglycerides into free fatty acids and monoglycerides. Additionally, proteins such as CTRB1, CTRL, and CPA1 were proteases involved in protein digestion. In addition to digestion, pancreas-associated proteins also enriched in biological pathways related to insulin response. Proteins such as IGFBP2, GHR, and PLA2G1B have been linked to insulin sensitivity and implicated in the regulation of insulin signaling in preclinical studies and animal models^68–70^. These findings highlight the critical roles of pancreas in producing enzymes required for nutrient breakdown and regulating systemic insulin sensitivity and metabolic responses through these pancreas-associated proteins.

Moreover, 113 proteins associated with spleen volume (|β| range = (0.08, 0.39), *P* range *=* (1.61× 10^−8^, 4.64× 10^−145^)). These proteins had enriched interaction (mean PPI score = 0.12, enrichment *P* < 2.2× 10^−16^) and pathway enrichment analysis underscored the spleen’s role in modulating both innate and adaptive immunity. Spleen-associated proteins were significantly enriched in the regulation of leukocyte-mediated immunity (adjusted *P* = 1.43 × 10^−16^) and lymphocyte-mediated immunity (adjusted *P* = 6.67× 10^−15^, **Fig. 2G**). These proteins play diverse roles in immune activation, migration, and function, particularly involving T cells, B cells, and natural killer (NK) cells. For example, LAG3 functions as an immune checkpoint to prevent T cell overactivation and maintain immune homeostasis^71^. IL12B and TNF were pro-inflammatory cytokines that regulate T cell and NK cell activity^72,73^, while ICAM1 mediate leukocyte adhesion to endothelial cells^74^. In addition, CR1 and CR2 modulate B cell activation and are implicated in autoimmune diseases such as systemic lupus erythematosus and rheumatoid arthritis^75,76^.

### Plasma proteomic insights into the brain and eye

We identified 33 plasma proteins that were associated with global and regional brain volumes measured by brain sMRI. These proteins were widely enriched in biological pathways that were crucial to neural development and nervous system function, such as axonogenesis, axon guidance, synapse maturation, and neuron projection guidance (adjusted *P* < 0.046). Most of these proteins were linked to total brain volume, white matter volume, and gray matter volume (|β| range = (0.08, 0.24), *P* < 1.55× 10^−8^). Some of them were also linked to volumes of localized regions. For example, increased level of NCAN and BCAN were associated with increased volumes of the limbic regions, such as frontal pole, insular cortex, hippocampus, and amygdala (β range = (0.11, 0.19), *P* range = (1.27× 10^−14^, 6.75× 10^−40^)). NCAN and BCAN were highly interacted proteins (PPI score = 0.96, **Fig. S9**). They are both chondroitin sulfate proteoglycans, key components of the extracellular matrix and are enriched in critical pathways such as the perineuronal net, perisynaptic extracellular matrix, and synapse-associated extracellular matrix (adjusted *P* < 0.03). The links between NCAN and BCAN and brain volumes were reported before and were associated with cognitive ability^20^. Additionally, elevated level of MOG and SLITRK1 were also associated with higher volumes of limbic system for emotion regulation, such as the frontal pole, thalamus, insular cortex, hippocampus, and amygdala (β range = (0.11, 0.18), *P* range = (4.55× 10^−10^, 4.18× 10^−32^)). SLITRK1 is highly expressed in the brain and regulates excitatory synapse formation and neural connectivity, particularly in hippocampal neurons^77^. It was also implicated in the pathogenesis of Tourette syndrome^78,79^ and has been found in schizophrenia patients^80^. MOG was expressed exclusively in central nervous system (CNS) myelin^81^ and can cause myelin oligodendrocyte glycoprotein antibody-associated disease (MOGAD) when it was attacked by the immune system. Brain structural and functional changes have been reported in individuals with MOGAD, including gray matter atrophy of frontal and temporal lobe, insula, thalamus, and hippocampus^82^. Most of the observed associations between brain volumes and plasma proteins were positive, with only few exceptions such as SOST, FABP3, GDF15, and LEP, whose increased levels in plasma associated with decreased volume of certain brain regions.

Six proteins were associated with white matter microstructural measures of brain dMRI, with two (BACN and NCAN) demonstrating overlap with sMRI. Additional proteins include AHSP, GFAP, OMG, and TF (|β| range = (0.08, 0.13), *P* < 1.45× 10^−8^, **Fig. S10**). Notably, over 70% of the dMRI-protein associations (26 out of 37) involved GFAP, which showed broad associations with multiple white matter tracts (|β| range = (0.08, 0.13), *P* < 1.37× 10^−8^). GFAP, a key astrocyte protein, regulates CNS homeostasis and astrocyte responses to stress and neurological disease^83^. Blood GFAP levels serve as a sensitive biomarker for CNS injuries and diseases, aiding in diagnosis, severity assessment, and prognostication in conditions such as traumatic brain injury and multiple sclerosis^84^. These dMRI-associated proteins were enriched in pathways that were critical for the formation, maintenance, and functional integrity of brain white matter, including glial cell differentiation, positive regulation of neurogenesis, nervous system development, and neuroblast proliferation (adjusted *P* < 0.046, **Fig. S11**). No significant associations were found between plasma proteins and brain resting state fMRI traits with the conservative Bonferroni correction. With a less stringent false discovery rate correction (*P* < 9.33× 10^−4^), several plasma proteins were identified as likely being associated with fMRI functional activity traits (|β| range = (0.04, 0.07), *P* < 9.11× 10^−4^, **Fig. S12**), with IGFBP2 emerging as a key protein linked to multiple networks (|β| range = (0.04, 0.06), *P* < 5.89× 10^−4^).

Eye OCT measures of the retina, a component of the CNS closely connected to the brain^85^, showed substantial associations with plasma proteins. Specifically, 93 plasma proteins linked to the average thickness of ganglion cell-inner plexiform layer (GCIPL, |β| range = (0.08, 0.12), *P* < 1.52× 10^−8^) and the retinal nerve fiber layer (RNFL, |β| range = (0.08, 0.13), *P* < 1.54× 10^−8^). Some of these proteins have been highlighted in previous eye research. For example, SOD1 deficiency leads to oxidative stress, resulting in retinal pigment epithelial damage and features of age-related macular degeneration^86^. Similarly, CA1 regulates intraocular pressure by influencing aqueous humor production and has been implicated in glaucoma^87^. Most of the eye OCT-associated proteins (87 of 93, over 94%) were specific to the eye, with minimal overlap observed with other organs. Most proteins were positively associated with GCIPL thickness and negatively associated with RNFL thickness, except for OMG, which showed the opposite effect and was also the only plasma protein shared between eye OCT traits and brain IDPs. In addition, 39 proteins are associated with eye fundus images features (|β| range = (0.07, 0.10), *P* < 1.62× 10^−8^, **Fig. S13**). The majority (27/39) of these proteins overlapped with those associated with eye OCT measures, emphasizing the shared proteomic links of these ocular imaging traits.

### Tracing the putative origins of proteomic effects on imaging traits

Plasma proteins are produced by various organs and tissues throughout the body. Understanding the origins of the identified proteomic effects on IDPs can provide deeper insights into their biological roles and cross-organ interactions, particularly when proteins related to one organ imaging trait may originate from another. The putative origins and organ-specific plasma proteins can be inferred using external organ/tissue-specific RNA sequencing data^38^. Specifically, plasma proteins can be considered organ-specific^13–15^ if their gene expression in a particular organ is at least four times higher than in any other organ^19^. Following this definition, 19% of plasma proteins (557 out of 2,923) could be traced back to a single organ, accounting for 28% of the identified phenotypic protein-imaging associations (1,415 out of 5,067). For each organ, we evaluated whether the identified imaging-associated proteins were enriched among these organ-specific plasma proteins (**Table S7**). We found that, in some organs, proteins linked to imaging traits were enriched in the set of highly expressed genes from the same organ. Conversely, in other cases, imaging-associated proteins in one organ were enriched in genes highly expressed in a different organ (**Figs. 3A-3B**). This raises the possibility of distant biologic effects, although alternative explanations exist. For example, these relationships may result from organ cross-talk through other mechanisms (neurogenic) or reflect overarching causative factors affecting imaging traits in one organ and protein expression in a different organ without a direct biologic effect the identified proteins across organs. The overall enrichment patterns are detailed below.

**Fig. 3.**
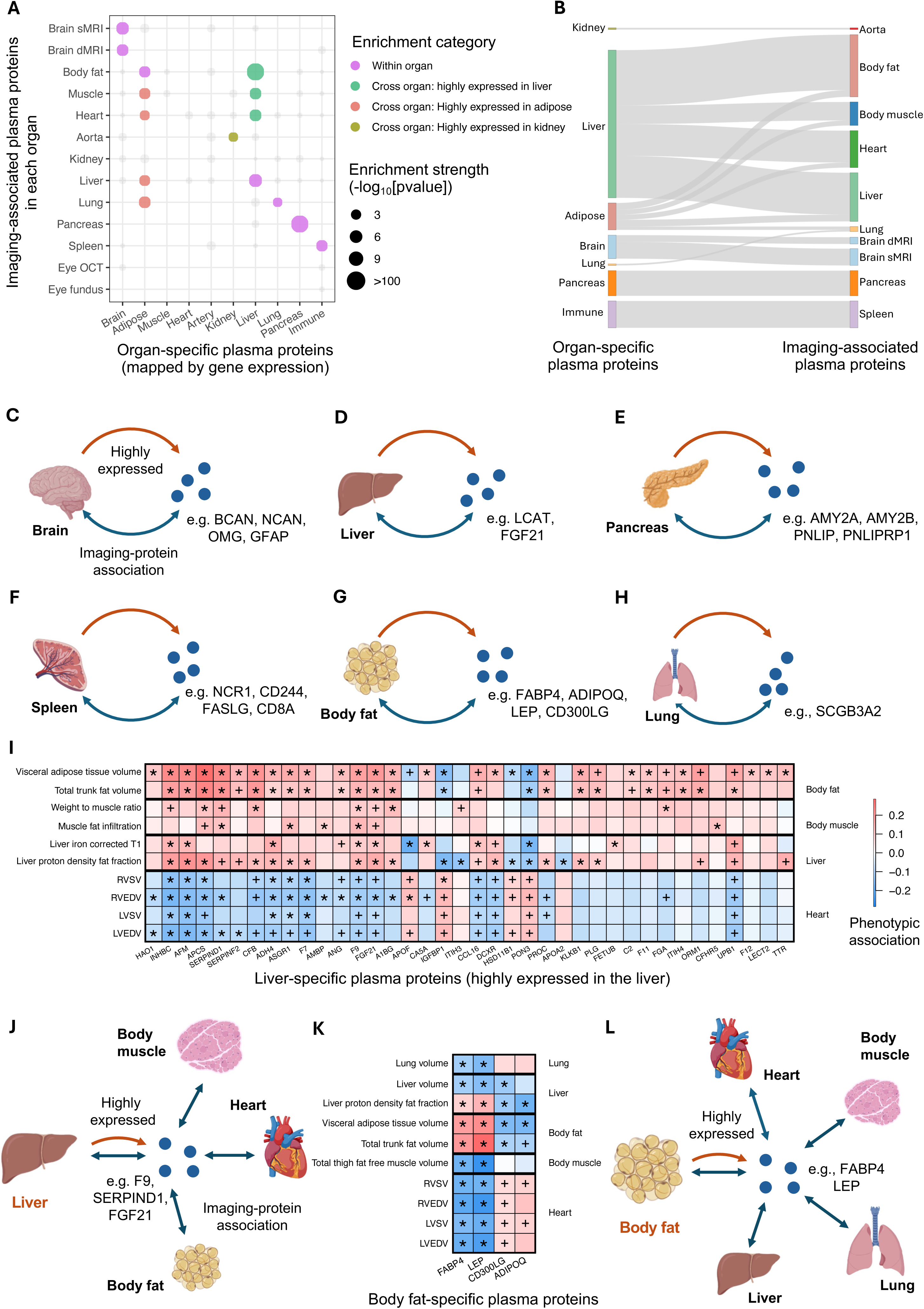
Tracing putative origins of proteomic effects on imaging traits. **(A)** Bubble plot that highlights whether proteins associated with imaging traits of an organ (*y*-axis) were significantly enriched among genes highly expressed in the same or another organ (*x*-axis). Bubble size reflects the enrichment significance level and bubble colors differentiate between within-organ or cross-organ enrichments. Only significant enrichments were highlighted in colors. For example, proteins associated with imaging data from four organs/tissues were significantly enriched among those with high gene expression in the liver. This included the liver itself (within-organ) as well as adipose tissue, muscle, and heart (cross-organ). **(B)** Overview of putative origins of proteomic effects on imaging traits. The left axis represents the organs where proteins exhibit phenotypic associations with imaging traits, while the right axis indicates the proteins’ original organs mapped by gene expression data. The thickness of the connections between the axes reflects the relative number of proteins contributing to each connection. For example, proteins with high gene expression in the liver (right axis) were associated with IDPs of the liver, heart, body muscle, and body fat (left axis). **(C)-(H)** Example proteins demonstrating putative within-organ proteomic associations in each of the six organs/tissues showing significant within-organ enrichment in **(A)**. For example, in **(C)**, BCAN, NCAN, OMG, and GFAP were associated with brain sMRI and dMRI traits and exhibited high gene expression in the brain. **(I)-(J)** Example of putative cross-organ associations. In **(I)**, we show correlation coefficients between plasma proteins highly expressed in the liver (*x*-axis) and imaging traits of the heart, liver, body muscle, and body fat with which these proteins were phenotypically associated (*y-*axis). The color represents correlation estimates. Coefficients that passed Bonferroni correction (*P* < 1.63× 10^−8^) and replicated were marked with asterisk, while those that passed Bonferroni correction but not replicated were marked with a plus sign. In **(J)**, we illustrate example proteins (F9, SERPIND1, and FGF21) that showed significant associations with the heart, body fat, body muscle, and liver and were highly expressed in the liver. **(K)-(L)** Example of putative cross-organ associations. In **(K)**, we show correlation coefficients between plasma proteins highly expressed in the body fat (*x*-axis) and imaging traits of the lung, liver, body fat, body muscle, and heart with which these proteins were phenotypically associated (*y-*axis). The color represents correlation estimates. Coefficients that passed Bonferroni correction (*P* < 1.63× 10^−8^) and replicated were marked with asterisk, while those that passed Bonferroni correction but not replicated were marked with a plus sign. In **(L)**, we illustrate example proteins (FABP4 and LEP) that showed significant associations with the heart, body muscle, lung, liver, and body fat and were highly expressed in body fat.

Proteins associated with IDPs of the brain (both sMRI and dMRI), liver, pancreas, spleen, body fat, and lung were significantly enriched among genes highly expressed in the same organ (enrichment *P* < 6.95× 10^−4^). For example, 4 of the 6 dMRI-associated proteins (BCAN, NCAN, GFAP, and OMG) had high gene expression in the brain (enrichment *P* = 4.40× 10^−7^), and 10 of the 33 sMRI-associated proteins were also defined to be brain-specific (enrichment *P* = 1.52× 10^−8^, **Fig. 3C**). These plasma proteins, likely originating from brain tissues, play specialized roles in brain structure and function^83,88^. Similarly, 25 of the 271 liver-associated proteins were highly expressed in the liver (enrichment *P* = 2.54× 10^−9^, **Fig. 3D**), almost all of which are known to be primarily produced in the liver (such as LCAT^58^ and FGF21^60^) and involved in key liver functions such as detoxification, metabolism, lipid transport, and blood coagulation. Among pancreas-associated proteins, 15 of the 26 were pancreas-specific (such as AMY2A, AMY2B, PNLIP, and PNLIPRP1, enrichment *P* = 0, **Fig. 3E**), reflecting the pancreas’s specialized role in digestion. Similar enrichment patterns were also observed on spleen, body fat, and lung (enrichment *P* < 6.95 × 10^−4^, **Figs. 3F-H** and **Supplementary Note**). Together, these findings demonstrate that proteins phenotypically associated with specific organs often have high organ-specific gene expression, highlighting their specialized roles in maintaining organ structure and function.

While many proteins associated with imaging data of specific organs are highly expressed in those same organs, some are predominantly expressed in other organs. Specifically, significant enrichments were observed among proteins produced in the liver, adipose tissue, and kidney (**Figs. 3A**), reflecting more interconnected, multi-organ regulatory mechanisms. Specifically, 19 heart-associated proteins (enrichment *P* = 2.80× 10^−7^) and 33 body fat-associated proteins (enrichment *P* = 0) were highly expressed in the liver, reflecting its systemic regulatory role across multiple physiological processes (**Figs. 3I-3J**). Many of these liver-produced heart-associated proteins (such as F7, F9, FGA, SERPIND1, and CFB) are crucial to thrombosis and maintaining heart health. For example, F7 is a coagulation factor that promotes thrombosis. It is also associated with coronary artery disease, which is strongly influenced by cholesterol and triglyceride levels^89^. Specific genetic variants of *F7* were associated with lower F7 levels and lower risk of myocardial infarction^90^. Another coagulation factor, F9, was a risk factor of deep venous thrombosis^91^, and F9 activation was showed in patients with acute coronary syndromes^92^. Higher levels of F9 have been observed in obese individuals^93^. SERPIND1 has protective effect on atherosclerosis and restenosis^94^. Additionally, liver-produced body fat-associated proteins, such as FGF21 and IGFBP2, are involved in energy balance, and lipid and glucose metabolism^60,95^. These findings suggest the connection between liver-produced proteins and their links to cardiovascular and metabolic regulation, highlighting the liver’s pivotal role in heart health and systemic metabolism.

Moreover, many proteins associated with body muscle, liver, and heart were produced in the adipose tissue, such as CD300LG, FABP4, LEP, and ADIPOQ (enrichment *P* < 1.18× 10^−4^, **Figs. 3K-3L**). These proteins tend to have pleiotropic roles across various biological processes. FABP4, secreted by adipocytes, is implicated in cardiometabolic risk, atherosclerosis, coronary artery disease, and cardiac dysfunction, serving as both a potential biomarker and a contributor to cardiac metabolism^96^. It also serves as a predictive marker for the progression of nonalcoholic fatty liver disease to nonalcoholic steatohepatitis, indicating patients at higher risk of liver disease complications^97^. LEP, primarily produced by adipose tissue, plays a key role in regulating cardiac metabolism, preventing cardiac lipotoxicity, and is associated with various cardiovascular complications. Furthermore, it plays a crucial role in liver health by preventing lipid accumulation and facilitating lipid mobilization^98^. These findings highlight the systemic influence of adipose-derived proteins in coordinating metabolic and physiological processes across organs, emphasizing the interconnected roles of adipose tissue in maintaining whole-body homeostasis. Additionally, one aorta-associated protein, REN, was produced by the kidney. REN activates the renin-angiotensin system, with angiotensin II causing blood vessel constriction and increasing blood pressure, which places additional strain on the aorta. This highlights the kidney’s indirect yet critical influence on aortic health via protein regulation^99^.

### Organ-wise chart of proteomic prediction accuracy and key predictors

Given the strong associations between plasma proteins and whole-body IDPs, we assessed their combined predictive power for organ structure and function and identified the top-ranked proteins for IDP prediction. Using an elastic-net model, we resampled half of the subjects for training and testing, repeated this process 200 times, evaluated the mean prediction *R*-squared (*r*^2^) of plasma proteins for IDPs, and selected the top robust predictors^16^ (**Methods**). Abdominal MRI traits showed the highest predictive power, particularly for body fat IDPs (median *r*^2^ range = (0.45, 0.55), **Fig. 4A**). The full set of 2,923 proteins generally outperformed the subset of 557 organ-specific proteins (i.e., proteins whose genes are highly expressed in a specific organ) in predicting IDPs across all organs (**Fig. S14**). Nevertheless, we observed that the top-ranked predictors for IDPs of the brain, body fat, liver, pancreas, and lungs were predominantly organ-specific proteins corresponding to those respective organs. Below we highlight the IDPs for which plasma proteins demonstrated high predictive power. Complete results, including prediction performance and top five predictors for each IDP, are provided in **Table S8**.

**Fig. 4.**
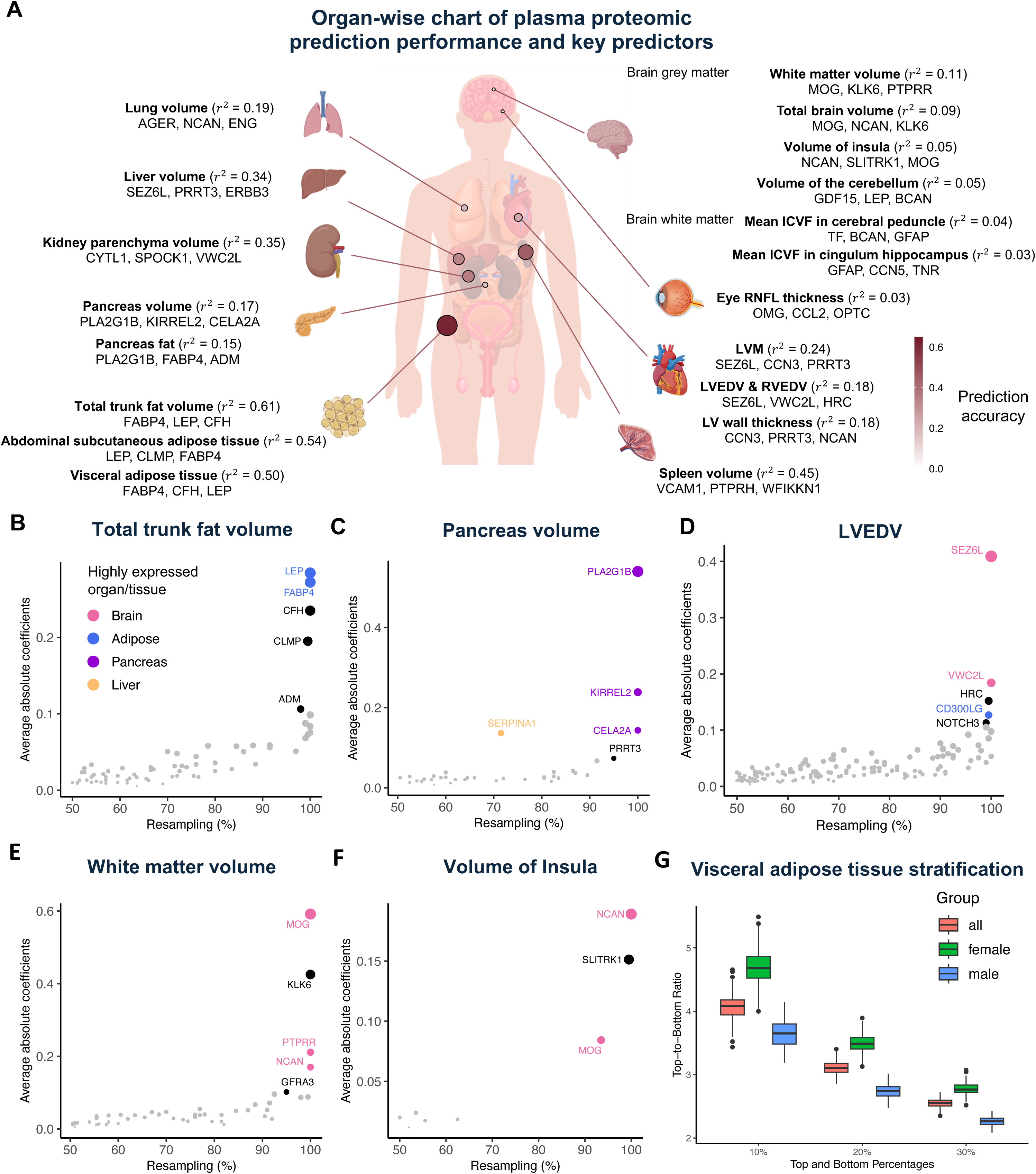
Predictive power of plasma proteins on organ structure and function. **(A)** Predictive accuracy of selected imaging traits across all organs, including the brain, eye, heart, spleen, lung, liver, kidney, pancreas, and body fat, along with the top three plasma protein predictors for each trait. ICVF refers to the intra-cellular volume fraction and RNFL refers to retinal nerve fiber layer. LVM stands for left ventricular myocardial mass, while LVEDV and RVEDV represent left and right ventricular end-diastolic volumes, respectively. **(B)-(F)** Top-ranked plasma proteins for selected imaging traits. The *x*-axis represents the frequency of proteins appearing across 200 resampling iterations, while the *y*-axis represents the average absolute value of coefficients across the 200 resampling iterations. Top proteins are labeled with their names and are further highlighted in color if this protein is highly expressed in certain specific organ (at least 4 times higher than any other organs). We illustrate the results for total trunk fat volume (**B**), pancreas volume (**C**), left ventricular end-diastolic volume (LVEDV, **D**), volume of cerebral white matter (**E**), and volume of the insular cortex (**F**). **(G)** The top-to-bottom ratios of average visceral fat volume for percentile groups representing the top and bottom 10%, 20%, and 30% of predicted imaging traits based on plasma protein-based prediction model. The *x*-axis represents the percentile groups, and the *y*-axis represents the top-to-bottom ratios. Sex groups, including all individuals, females, and males, are distinguished by color.

The highest predictive power was observed for abdominal MRI traits (median *r*^2^ range = (0.10, 0.55), **Fig. S15**). For some organs and tissues, including body fat, liver, pancreas, and lung, top protein predictors for related IDPs were primarily organ-specific. This makes the prediction power of the full set of plasma proteins similar to that of the subset primarily expressed in these organs. Among these, body fat traits showed the highest prediction accuracy, particularly for total trunk fat volume, total abdominal adipose tissue index, and abdominal fat ratio (*r*^2^ range = (0.57, 0.61), **Figs. 4B** and **S16-S17**). Key predictors, such as FABP4 and LEP, which are primarily secreted by adipose tissue, consistently and strongly predicted all body fat imaging traits with high power. CLMP and CFH also demonstrated strong predictive power for body fat. CLMP is involved in adipocyte differentiation and linked to the progression of obesity^100^. Plasma proteins also showed strong prediction performance for liver IDPs. Among these, liver volume had the highest prediction power (*r*^2^ = 0.34), with top predictors, such as SEZ6L and VWC2L, primarily expressed in the brain (**Fig. S18**). Additionally, plasma proteins effectively predicted liver inflammation and fibrosis (*r*^2^ = 0.20, **Fig. S19**), with key predictors, such as APOF and SERPINA6, predominantly expressed in the liver. APOF regulates hepatic lipoprotein metabolism by enhancing low-density lipoprotein triglyceride release and promoting the clearance of lipoprotein remnants and is associated with hepatic steatosis^101^. For pancreas IDPs, plasma proteins predicted pancreas volume and pancreas fat fraction with moderate power (*r*^2^ range = (0.15, 0.17)). Top predictors for pancreas volume included PLA2G1B, CELA2A, and KIRREL2 (**Fig. 4C**), all primarily produced in the pancreas. PLA2G1B and CELA2A, produced by pancreatic acinar cells, are involved in digesting dietary proteins and phospholipids and are linked to insulin sensitivity^70,102^. KIRREL2, primarily expressed in pancreatic beta cells, may play a role in beta cell function and pancreas development^103^. For pancreas fat fraction, PLA2G1B was also the strongest predictor. Additionally, LEP and FABP4, which were key predictors for body and visceral fat, showed strong predictive power for pancreas fat fraction as well (**Fig. S20**). In the lung, the strongest predictor of lung volume was AGER, which is highly expressed in lung tissue. Together with other key proteins such as NCAN, LEP, ENG, and CD93, plasma proteins predicted lung volume with a prediction *r*^2^ of 0.19 (**Fig. S21**).

Plasma proteins also show strong predictive power for IDPs of body muscle, spleen, and kidney, despite the genes of top predictors not being highly expressed in these corresponding organs. Among these, spleen volume showed the highest predictive power ( *r*^2^ = 0.45), with top predictors including VCAM1, CRLF1, SEMA7A, PTPRH, and WFIKKN1 (**Fig. S22**). Many of these proteins are key players in inflammation and immune responses. For example, VCAM1 regulates leukocyte adhesion to blood vessel walls and their passage through the endothelial layer^104^, while SEMA7A, expressed on activated T cells, stimulates macrophages to produce proinflammatory cytokines, driving inflammatory immune responses^105^. For body muscle, weight-to-muscle ratio and muscle fat infiltration had the highest predictive power (*r*^2^ range = (0.35, 0.44), **Figs. S23-S25**). Key proteins such as CLMP, ART3, and RGMA consistently predicted multiple muscle-related traits. Notably, CLMP also demonstrated strong predictive power for body fat traits, highlighting its dual role in muscle and fat-related processes. Similarly, LEP and FABP4, both key predictors of body fat traits, also strongly predicted muscle traits like muscle fat infiltration in the posterior thigh and weight-to-muscle ratio, further emphasizing the overlap in proteins driving fat and muscle composition. Additionally, plasma proteins also showed robust predictive ability for kidney IDPs, particularly kidney parenchyma volume and kidney volume (*r*^2^ range = (0.24, 0.35), **Figs. S26-S27**). Top predictors for kidney traits included SPOCK1, VWC2L, and LRTM2, which are highly expressed in the brain, and CYTL1, which is highly expressed in the aorta.

Similar to the kidney, specific proteins highly expressed in the brain also dominate the prediction of MRI traits of the heart and aorta. Heart IDPs with the highest predictive performance include myocardial mass and ventricular end-diastolic volumes (both left and right) as well as global left ventricular myocardial-wall thickness at end-diastole (*r*^2^ range = (0.18, 0.24), **Fig. S28**). For the aorta, IDPs with high predictive power were related to the descending aorta, particularly descending aorta minimum and maximum areas (*r*^2^ = 0.11). SEZ6L and VWC2L, highly expressed in the brain, consistently predicted IDPs of multiple organs, including left ventricular myocardial mass and end-diastolic volumes, as well as liver volume, emphasizing their significance across multiple systems (**Figs. 4D** and **S29-S32**). Notably, predictors for left ventricular myocardial-wall thickness differed from those of other IDPs. Top predictors for left ventricular myocardial-wall thickness include CCN3, REN, and CHGB (**Fig. S33**), many of which are involved in cardiac remodeling and hypertrophy. For example, REN activates the renin-angiotensin system and contributes to left ventricular hypertrophy^99^. Similarly, CCN3-deficient mice exhibit septal thickening, hypertrophic cardiomyopathy, and ventricular dilation, highlighting CCN3’s role in regulating myocardial-wall thickness and maintaining overall heart health^106^. Additionally, CHGB plays a critical role in catecholamine secretion, and genetic variations in *CHGB* have been associated with increased risks of hypertension^107^, which likely contribute to the relationship between CHGB and left ventricular myocardial-wall thickness.

Among brain IDPs, relatively high predictive performance was observed for global measures, including total brain volume, white matter volume, and grey matter volume (*r*^2^ range = (0.08, 0.10)). Many top predictors (such as MOG, PTPRR, and NCAN) were highly expressed in the brain (**Figs. 4E** and **S34-S36**). These proteins, along with other strong predictors such as KLK6, GFRA3, and SLITRK1, were associated with structural brain IDPs and contributed to brain structure, function, or development. Regional brain volumes generally had smaller prediction power than global ones. For example, plasma proteins predicted volumes of the cerebellum VIIIa/VIIIb, insular cortex, thalamus, amygdala, and hippocampus with moderate accuracy (*r*^2^ range = (0.03, 0.05)). NCAN, MOG, and BCAN, highly expressed in the brain, were the strongest predictors for these regions, except for the cerebellum (**Figs. 4F** and **S37-S39**). In contrast, protein predictors for cerebellum volumes differed and varied slightly across subregions. GDF15 and LEP consistently predicted the volumes of both left and right cerebellum VIIIa/VIIIb (**Fig. S40**). LEP influences neural activity in the posterior cerebellum, modulating brain responses to food-related stimuli and food intake-related plasticity^108^. More results for brain dMRI and eye are summarized in the **Supplementary Note** and **Figures S41-S47**. For example, one of the top predictors of brain dMRI was APCS (**Fig. S46**), which produced exclusively in the liver and has been linked to neurodegenerative diseases (such as Alzheimer’s disease) when it appears in the brain due to compromised blood-brain barrier integrity. APCS is cytotoxic to cerebral neurons, promotes Aβ amyloid formation, highlighting an inter-organ proteomic relationship in disease pathology^109,110^.

### Stratification capability of plasma protein prediction models

The developed plasma protein-based prediction models for IDPs can be used to stratify individuals with abnormal organ structure and function without requiring actual imaging data. Such applications take advantage of the greater accessibility of blood-based measurements compared to imaging modalities like whole-body MRIs. In this section, we evaluated the stratification capability by calculating the ratio of true IDP values between the top and bottom 10% of protein-predicted IDP deciles^111^. Consistent with the cohort-level prediction accuracy, IDPs related to body organ fat and visceral fat exhibited the highest individual-level stratification ratios. The largest difference was observed for VAT, where the true VAT volume in the top 10% of protein-predicted VAT was over four times higher than in the bottom 10% (**Fig. 4G**). This demonstrates that plasma proteins-trained computational models can effectively stratify individuals with high VAT volumes. Other body fat traits, such as total trunk fat volume and total abdominal adipose tissue index, also showed substantial stratification (median ratio > 3.5, **Fig. S48**). Stratifications were similarly effective for organ-associated fat volumes, such as liver fat fraction and pancreas fat fraction (median ratio > 2.8, **Fig. S49**).

Plasma protein prediction models also effectively stratify volumetric traits of multiple organs, with spleen volume exhibiting the highest stratification ratio (median ratio > 2.6, **Fig. S50**). Stratification for other organ volumes, including the kidney, liver, lung, and pancreas, showed consistent patterns (median ratio > 1.4, **Fig. S51**). For cardiac MRI traits, left ventricle end-diastolic volume, left ventricular myocardial mass, and right ventricle end-systolic volume had modest stratification (median ratio > 1.3, **Fig. S52**). For brain IDPs, both global volumes such as cerebral white matter (median ratio > 1.12, **Fig. S53**) and regional volumes such as the amygdala and insular cortex (median ratio > 1.07, **Fig. S54**) demonstrated stratification capability. Importantly, we found that the stratification performance is generally robust over time. Due to the UKB data collection procedures, plasma samples and imaging scans were collected at different visits, with a gap of 3 to 17 years between collections depending on the individual. We found that this time gap has only a minor impact on stratification performance, as illustrated by body and visceral fat traits (**Fig. S55**) and kidney and lung volumes (**Fig. S56**). Overall, plasma protein-based stratification is particularly valuable for whole body images that are not routinely accessible in healthy large-scale cohorts. Computational models with plasma protein levels from blood samples provide a promising tool for evaluating the health of organs and their specific regions.

### Genetic-root putative causal links between plasma proteins and imaging traits

Mendelian randomization (MR) analysis was performed to identify plasma proteins with genetic-driven putative causal links to IDPs (**Methods**). We tested 2,856 proteins using their *cis*-protein quantitative trait loci (pQTL) variants with inverse variance weighted^112^ and Wald-ratio methods^113,114^ (**Table S9**). To ensure robustness, several sensitivity tests^115^ were performed to validate the MR assumptions, and colocalization analysis^33^ were further conducted to identify protein-imaging pairs sharing causal variants (**Table S10**). After Bonferroni correction (*P* < 2.98× 10^−8^) and excluding results that did not pass sensitivity tests, we identified 8,116 significant protein-IDP genetic causal pairs involving 318 proteins and 1,041 IDPs, with 448 of these pairs (98 proteins and 334 IDPs) showing PPH4 > 80% in the colocalization analysis (**Fig. 5A**). The MR associations of these 318 proteins were broadly shared across organs, with 250 influencing more than one IDP (**Fig. 5B**). These findings indicate that genetic-driven proteomic links are often pleiotropic, affecting multiple organs. Below we report the overall patterns and highlight key genetically causal plasma proteins for each organ.

**Fig. 5.**
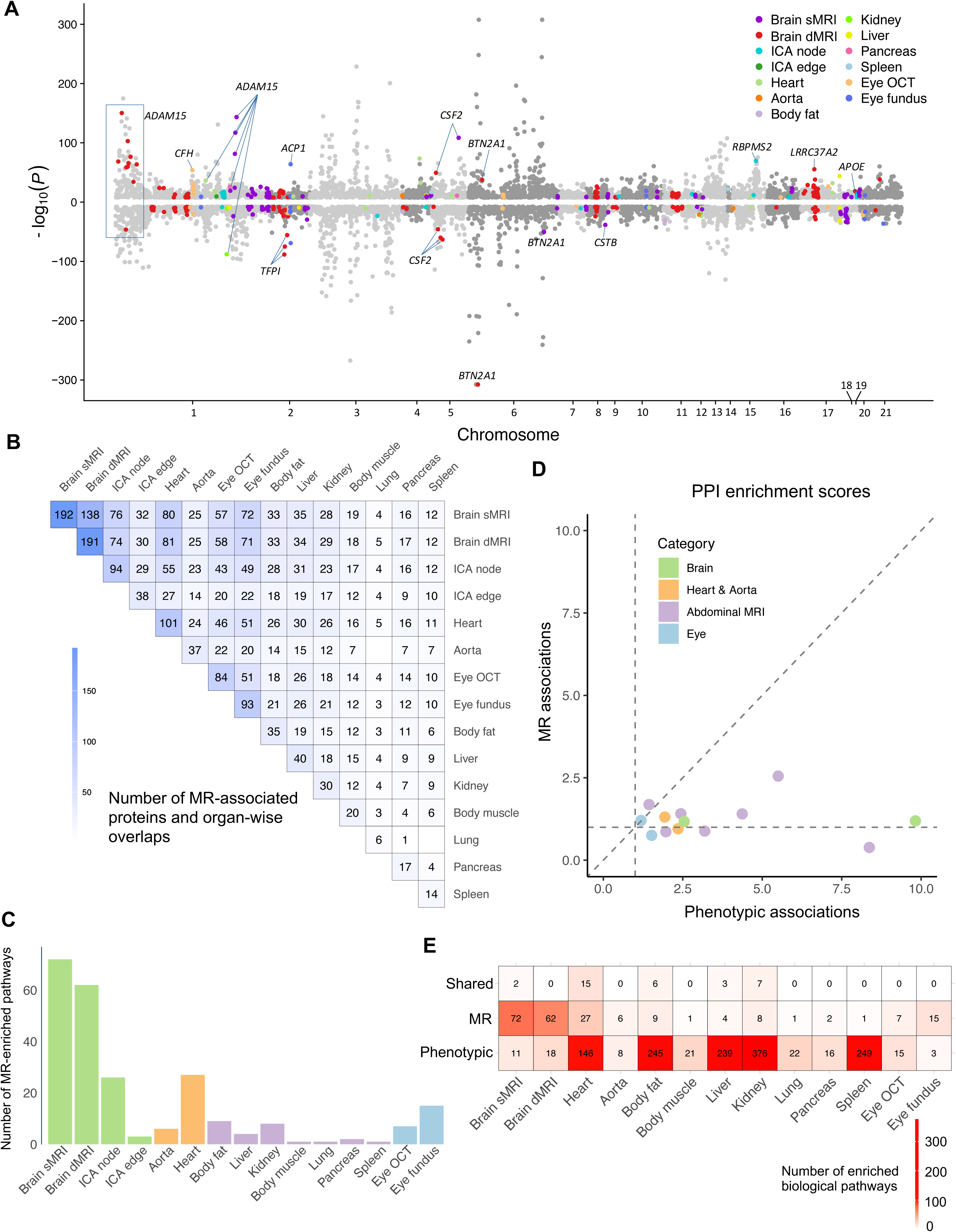
Genetic-root putative causal protein-imaging links. **(A)** Putative Causal associations between plasma proteins and imaging traits identified by Mendelian randomization (MR). The *x*-axis represents the chromosomes where the proteins are located, the *y*-axis represents the -log10(P-value) of protein-imaging associations. Only protein-imaging pairs significant under Bonferroni correction from the MR analysis were plotted (*P* < 2.98× 10^−8^). Pairs with strong evidence of sharing causal variants in Bayesian colocalization analysis (PPH4 > 80%) were highlighted in organ-specific colors. Brain sMRI stands for brain structural MRI, brain dMRI stands for brain diffusion MRI, ICA edge and ICA nodes are brain functional MRI traits generated by independent component analysis, OCT stands for optical coherence tomography. **(B)** The number of significant proteins associated with each organ after the Bonferroni correction (*P* < 2.98× 10^−8^) in MR analysis and the overlapping pattern across organs. **(C)** Overview of the number of MR-enriched biological pathways in each organ. **(D)** Comparison of protein-protein interaction (PPI) enrichment scores between phenotypically and causally associated proteins across organs. PPI scores were calculated using the STRING database for proteins associated with each organ. The *x*-axis shows PPI enrichment scores for phenotypically associated proteins, while the *y*-axis shows scores for MR-associated proteins. Points are colored by organ. Dashed lines mark an enrichment score of one, representing the baseline level of including all proteins. **(E)** Number of enriched pathways for phenotypically associated and MR-associated proteins, as well as the number of pathways shared between the two approaches.

Plasma proteins demonstrated extensive MR associations with brain IDPs, including regional brain volumes, white matter microstructure, and fMRI traits. Such strong MR association power resulted in the identification of numerous enriched biological pathways^40^ in the brain (**Fig. 5C** and **Table S11**). Approximately 60% of the identified proteins (over 190) exhibited genetic causal links to brain sMRI and dMRI IDPs (|β| range = (0.01, 0.67), *P* < 2.98× 10^−8^), with 140 proteins affecting both modalities. Two examples of overlapping proteins are GRN and CTSB. Elevated GRN level was linked to increased thalamus volumes (β range = (0.12, 0.13), *P* < 3.12× 10^−10^, PPH4 range = (98.52%, 99.21%), **Fig. S57**) and reduced mean orientation dispersion index in the fornix cres and stria terminalis (β = -0.15, *P* = 6.32× 10^−12^, PPH4 = 97.53%, **Fig. S58**). Elevated CTSB level was associated with reduced grey matter volume in the left inferior frontal gyrus pars opercularis (β = -0.09, *P* = 3.70× 10^−39^, PPH4 = 80.00%, **Fig. S59**) and impaired white matter integrity in uncinate fasciculus, anterior corona radiata, and cingulum hippocampus tracks (β range = (-0.10, -0.08), *P* range = (3.09× 10^−32^, 1.20× 10^−37^)). Both GRN and CTSB are known therapeutic targets for neurodegenerative diseases such as Alzheimer’s disease and frontotemporal dementia^116,117^. We provide more examples regarding BCAN, OMG, FOXO1, PCDH7 in the **Supplementary Note** and **Figures 6A** and **S60-S66**. In addition, although plasma proteins showed no phenotypic associations with fMRI traits, genetic causal relationships were observed for both functional activity (94 proteins, |β| range = (0.01, 0.57), *P* < 2.95× 10^−8^) and functional connectivity measures (38 proteins, |β| range = (0.02, 0.60), *P* < 2.73× 10^−8^). For example, APOE had a genetic causal link with functional activity (amplitude) in the default mode and central executive networks (β = 0.06, *P* = 5.40 × 10^−9^, PPH4 = 99.71%, **Fig. 6B**). APOE is a well-documented genetic risk factor for Alzheimer’s disease, with the ε4 allele increasing risk by impairing Aβ clearance and promoting aggregation, contributing to Aβ pathology^118^.

**Fig. 6.**
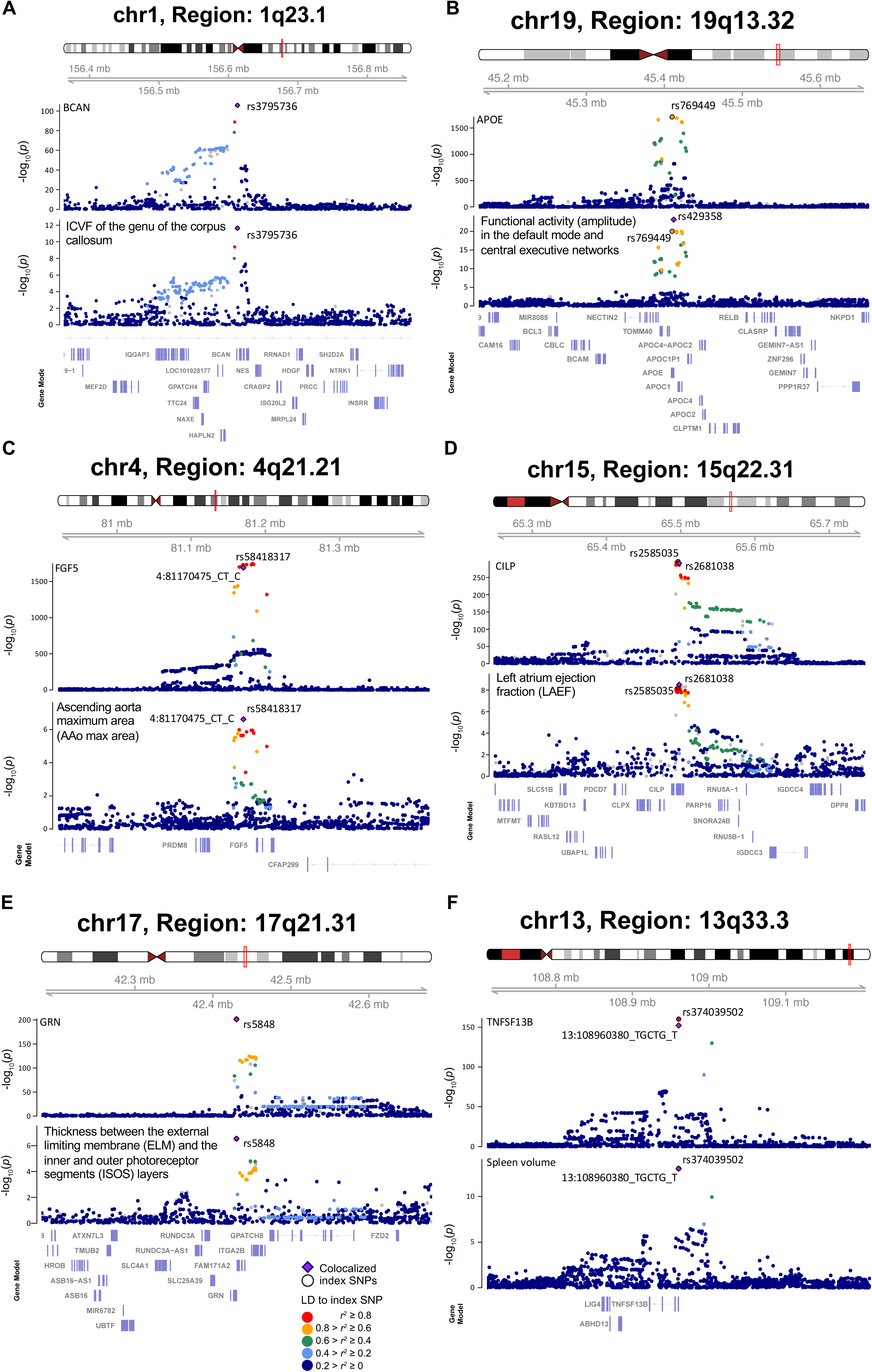
Selected associations identified by Mendelian randomization (MR) with shared causal variants. **(A)** In MR analysis, BCAN was causally associated with mean intracellular volume fraction (ICVF) in the genu of the corpus callosum. The posterior probability of Bayesian colocalization analysis for the shared causal variant hypothesis (PPH4) is 100.00%. **(B)** APOE was causally associated with functional activity (amplitude) trait in the default mode and central executive networks. The PPH4 of Bayesian colocalization analysis is 99.71%. **(C)** FGF5 was causally associated with ascending aorta maximum area (AAo max area). The PPH4 of Bayesian colocalization analysis is 98.72%. **(D)** CILP was causally associated with left atrium ejection fraction (LAEF). The PPH4 of Bayesian colocalization analysis is 98.26%. **(E)** GRN was causally associated with the average thickness between the external limiting membrane (ELM) and the inner and outer photoreceptor segments (ISOS) layers. The PPH4 of Bayesian colocalization analysis is 99.89%. **(F)** TNFSF13B was causally associated with spleen volume. The PPH4 of Bayesian colocalization analysis is 99.90%.

Among non-brain organs, plasma proteins had the strongest causal relationships with the heart and aorta (|β| range = (0.01, 0.59), *P* < 2.94× 10^−8^). A total of 114 plasma proteins (104 for heart and 37 for aorta) were identified to have MR associations with cardiac MRI traits, primarily involving end-diastolic/systolic volumes and ascending/descending aorta features. For example, increased level of FGF5 was causally associated with larger ascending aorta maximum area and minimum area (β = 0.05, *P* < 3.81× 10^−11^, PPH4 range = (98.72%, 99.26%), **Figs. 6C** and **S67**). FGF5 promotes angiogenesis in human aortic endothelial cells by enhancing vascular sprouting^119^ and has been linked to hypertension risk across multiple ethnicities in genetic studies^120,121^. Elevated levels of CILP were causally linked to a reduced left atrium ejection fraction (β = -0.11, *P* = 9.10× 10^−10^, PPH4 = 98.26%, **Fig. 6D**). CILP plays a key role in extracellular matrix remodeling and serves as a marker for cardiac fibrosis^122,123^. EFEMP1, another protein critical for maintaining ECM structure, was positively associated with right atrium stroke volume (β = 0.14, *P* = 2.41× 10^−10^, PPH4 = 93.57%, **Fig. S68**). EFEMP1 promotes proper scar formation after myocardial infarction, preventing cardiac rupture^124^. Additionally, elevated level of PDE5A was causally associated with increased right ventricular end-diastolic volume and stroke volume (β range = (0.12, 0.15), *P* range = (1.84× 10^−33^, 2.83 × 10^−74^), PPH4 range = (89.37%, 92.62%), **Fig. S69**). PDE5A is an approved therapeutic target for hypertension^125^, and its inhibition provides protective effects against cardiac stresses such as ischemia-reperfusion injury, drug toxicity, pressure-induced hypertrophy, and acute stress responses in the heart muscle^126^.

Genetic causal links were identified between plasma proteins and eye IDPs, including both OCT measures (84 proteins, |β| range = (0.01, 0.69), *P* < 2.79× 10^−8^) and fundus images (93 proteins, |β| range = (0.01, 0.48), *P* < 2.87× 10^−8^). Many of these linked proteins play critical roles in eye health, and some also have causal relationships with other organs, particularly the brain and heart. For example, GRN was causally associated with the average thickness between the external limiting membrane and the inner and outer photoreceptor segments layers (β = -0.11, *P* = 5.28× 10^−9^, PPH4 = 99.89%, **Fig. 6E**). GRN promotes the differentiation of retinal precursor cells into photoreceptor cells^127^ and supports retinal ganglion cell survival during development by regulating astrocyte activation^128^, highlighting its essential role in retinal regeneration and development. Similarly, EFEMP1 was causally linked to the vertical cup-to-disc ratio (β = -0.25, *P* = 5.00 × 10^−19^, **Fig. S70**), a trait associated with primary open-angle glaucoma. The Arg345Trp mutation in *EFEMP1* has been linked to drusen formation in Malattia Leventinese, closely resembling age-related macular degeneration pathology^129,130^. Moreover, CFH had a MR association with increased thickness between the inner nuclear layer and retinal pigment epithelium (β = 0.10, *P* = 1.10× 10^−54^, PPH4 = 93.71%). CFH plays a protective role in the eye by regulating complement activation and preventing inflammatory damage. Notably, the Y402H variant in *CFH* has been linked to increased risk of age-related macular degeneration^131,132^.

A total of 79 proteins showed MR associations with abdominal MRI IDPs, with the strongest relationships observed in the liver, spleen, and pancreas. Specifically, in the liver, elevated levels of RNF43 and APOH were causally associated with lower liver iron-corrected T1 (β range = (-0.06, -0.31), *P* range = (3.46× 10^−10^, 3.84× 10^−32^), PPH4 range = (98.87%, 99.84%), **Figs. S71-S72**), a measure of liver inflammation and fibrosis. RNF43 regulates WNT signaling, maintaining hepatocyte proliferation and differentiation balance, ensuring liver homeostasis and preventing liver cancer^133,134^. *APOH*, predominantly expressed in the liver^135^, has been linked to nonalcoholic fatty liver disease in genetic studies^136^. Elevated level of ADH4 was causally associated with increased liver volume (β = 0.25, *P* = 4.81× 10^−12^, PPH4 = 95.08%, **Fig. S73**). ADH4, which plays a role in alcohol metabolism, is downregulated in hepatocellular carcinoma and serves as a potential prognostic biomarker and therapeutic target^137,138^. Additionally, strong MR relationships were identified for spleen volume, with 114 plasma proteins showing significant associations (|β| range = (0.03, 0.42), *P* < 1.90 × 10^−9^)). Among these, TNFSF13B and EGF demonstrated the strongest links, with increased levels of both proteins linked to spleen enlargement (β range = (0.26, 0.42), *P* range = (7.70× 10^−18^, 3.99× 10^−23^), PPH4 range = (94.52%, 99.90%), **Figs. 6F** and **S74**). TNFSF13B, primarily expressed in the spleen, regulates B-cell survival, proliferation, and differentiation^139^, highlighting the spleen as a key site for its immune-regulatory function. EGF is essential for suppressing cellular senescence and sustaining growth in mammals, and inhibition of its receptor has been shown to cause spleen atrophy in mice^140^. MR associations were also observed for pancreas IDPs. For example, ADAM15 was linked to pancreatic iron levels (β = 0.03, *P* = 1.62× 10^−26^, PPH4 = 81.22%, **Fig. S75**). ADAM15 is overexpressed in pancreatic cancer cells, suggesting its role in tumor progression^141^. For the lung, IL4R was causally associated with lung volume (β = -0.05, *P* = 3.52× 10^−11^) and is a known drug target for chronic lung diseases such as asthma and pulmonary fibrosis. IL4R is critical for airway inflammation^142^, and polymorphisms in the IL4 promoter (C-589T) are associated with asthma^143^.

## Discussion

This study represents the largest imaging proteomics analysis to date. Using plasma protein and multi-organ imaging data from the UKB study, we identified novel phenotypic protein-imaging associations, shedding light on the roles plasma proteins play in organ-specific mechanisms and related biological pathways. These associations were largely robust to disease status, with a correlation over 99.8%, which is likely due to the predominantly healthy composition of the UKB cohort. Integration with external gene expression data revealed that some proteins associated with specific organs were secreted or highly expressed in the same organ, while others exhibited cross-organ relationships, notably involving proteins derived from adipose tissue and the liver. This finding underscores the liver’s central role in regulating systemic and metabolic processes. The predictive power of plasma proteins varied across organs, with abdominal tissues and organs showing the highest predictive accuracy. Genetic causal associations between plasma proteins and IDPs were identified using genetic instrumental variables, with approximately 5.5% further passing colocalization tests, indicating strong evidence of shared causal variant. As the most comprehensive pan-organ imaging proteomics analysis conducted within a single large-scale cohort, this study offers novel insights into the interplay between plasma proteins and organ-specific biology.

Building on these novel findings, our study bridges molecular-level biological processes with structural biology observed through whole-body imaging, revealing mechanisms underlying structural and functional changes in organs. For example, in the brain, consistent links were identified between specific proteins, such as BCAN, NCAN, MOG, and SLITRK1 and key brain regions, including the frontal pole, hippocampus, and amygdala, which are critical hubs of the limbic system involved in emotion regulation and the fight-or-flight response. The thalamus also emerged consistently, indicating the involvement in the reward systems. These proteins not only associate with these brain regions but also demonstrate strong predictive abilities, highlighting their potential to reveal molecular pathways underlying emotion regulation and motivational behavior. Furthermore, our results suggest that plasma proteins serve as robust non-invasive biomarkers for the heart and various abnormal organs, enabling early detection of organ dysfunction, large-cohort subject stratification, and the development of protein-targeted therapies for organ-specific and systemic diseases. Together, these findings enhance our understanding of molecular and structural biology and provide a foundation for future proteomic applications in precision medicine.

The phenotypic and genetic associations between proteins and imaging traits reveal distinct yet complementary patterns across different organs. Phenotypic association analysis identified more signals in non-brain organs, with particularly strong and extensive associations observed between plasma proteins and abdominal MRI traits. Notably, the set of proteins identified through phenotypic associations displayed higher levels of interaction (**Table S5**), greater enrichment in biological pathways (**Table S6**), and elevated gene expression within the same or other specific organs (**Table S7**). These findings provide deep insights into the proteomic networks and mechanisms underlying protein-imaging associations, highlighting how phenotypic analyses capture both organ-specific and organ-shared biological influences, potentially driven by environmental and non-genetic factors. These findings are consistent with previous phenotypic analyses of omics data, which have identified co-regulated clusters and protein modules enriched in network biology^7,144^.

In contrast, genetic causal proteins identified through the MR approach showed minimal overlap with phenotypically associated proteins. Notably, MR demonstrated greater power in the brain, identifying more putative causal proteins for brain IDPs, including fMRI traits that lacked power of identifying phenotypic associations. Of note, the set of proteins identified by MR consistently exhibited lower protein interaction scores (**Table S12**) compared to phenotypically associated proteins across most organs (**Fig. 5D**). Similarly, except for the brain and eye, MR-identified proteins had lower number of enriched biological pathways than phenotypically associated proteins (**Fig. 5E**). Specifically, there was minimal overlap between pathways linked to phenotypic and MR associations, except for the heart, body fat, liver, and kidney, where pathways enriched by phenotypically associated proteins largely encompassed those enriched by MR-associated proteins. Moreover, MR-identified proteins in most organs did not show significant enrichment in organ-specific gene expression, with the only exception of the lung (**Table S13**). These findings suggest that, compared to phenotypic analyses, MR-based genetic mapping may prioritize isolated proteins rather than interconnected protein co-regulated clusters and may be less connected to organ-specific biological mechanisms reflected in gene expression.

Notably, on the other hand, many MR-identified proteins are either approved or in-development drug targets^145^, with the associated diseases aligning with the organ of the imaging data. For example, APOE, GRN, BCHE, CTSB, and APCS, which have genetic causal associations with brain imaging traits, are drug targets for neurodegenerative diseases such as Alzheimer’s disease, dementia, cognitive impairment, and Parkinson’s disease. Similarly, proteins genetically linked to heart imaging traits serve as drug targets for cardiovascular diseases, including PLAU for myocardial infarction, AOC3, PDE5A, and SELE for hypertension, and LCAT for coronary artery disease. It is well established that genetic evidence is valuable for drug development and clinical trial success^146^. Our MR analysis provides key genetic insights into potential drug targets and highlights opportunities for leveraging imaging data in therapeutic target discovery process. Together, the differences between phenotypic and genetic results underscore the unique strengths of each approach, which may be helpful in different downstream applications. By combining phenotypic and genetic analyses, the present study provides a more comprehensive resource for understanding protein-imaging relationships.

## Data Availability

The individual-level data used in this study can be obtained from UK Biobank (https://www.ukbiobank.ac.uk/). The GWAS summary statistics of Olink plasma proteins (removing imaging subjects) produced in this study will be deposited in Zenodo upon publication. Other datasets in this paper include: the STRING database (https://string-db.org/), the Therapeutic Target Database (https://idrblab.net/ttd/), and the GTEx dataset v8 (https://gtexportal.org/).

## Acknowledgements

Research reported in this publication was supported by the National Institute On Aging under Award Numbers RF1AG082938 and 1R01AG085581. The content is solely the responsibility of the authors and does not necessarily represent the official views of the National Institutes of Health. The study has also been partially supported by funding from the Purdue University Statistics Department, Department of Statistics and Data Science at the University of Pennsylvania, Wharton Dean’s Research Fund, Analytics at Wharton, Wharton AI & Analytics Initiative, Perelman School of Medicine CCEB Innovation Center Grant, and the University Research Foundation Grant. The individual-level data UK Biobank used in this study were obtained under application 76139 subject to a data transfer agreement. We would like to thank the individuals who represented themselves in the UK Biobank for their participation and the research teams for their efforts in collecting, processing, and disseminating these datasets. We would like to thank the research computing and IT groups at the Wharton School of the University of Pennsylvania and the Rosen Center for Advanced Computing at the Purdue University for providing computational resources and support that have contributed to these research results.

## Author Contributions Statement

Z.F. and B.Z. designed the study. Z.F. analyzed the data and generated the results. X.Y., J.S., and Y.L. helped with data preprocessing and analysis. J.C., J.M.O., W.W., D.J.R., and R.G. provided feedback on study design and helped results interpretations. Z.F. and B.Z. wrote the manuscript with feedback from all authors.

## Methods

### Image-derived phenotypes of the brain and body

Image-derived phenotypes (IDPs) used in our study were based on data from the UKB study, which recruited approximately half a million participants aged 40 to 69 between 2006 and 2010^147^. The ethics approval of the UKB study was from the North West Multicentre Research Ethics Committee (approval number: 11/NW/0382) and informed consent was obtained by participants. Overall, we used 258 structural MRI (sMRI) traits capturing regional and total brain volumes derived from T1-weighted structural images, 432 diffusion MRI (dMRI) traits capturing white matter integrity through microstructural and tract-specific measures derived using tract-based spatial statistics (TBSS), 76 resting functional MRI (fMRI)-derived node amplitude traits and 6 global functional connectivity traits summarizing the pairwise coactivity of nodes, 82 cardiac and aortic MRI traits capturing global and regional metrics of four heart chambers and two aortic sections, and 46 derived OCT measures of retinal structure and 110 fundus image features extracted using transfer learning models, as well as 41 abdominal MRI traits capturing body fat and muscle composition, along with kidney, liver, lung, spleen, and pancreas characteristics. These IDPs captured the structural and functional characteristics of multiple human organs and tissues, including the brain, eye, heart, aorta, body fat, body muscle, liver, kidney, lung, pancreas, and spleen.

Brain IDPs used in this study were processed and generated by Alfaro-Almagro, et al. ^148^. Briefly, we used sMRI traits extracted by three pipelines, FMRIB’s automated segmentation tool^149^ (FAST), FMRIB’s integrated registration and segmentation tool^150^ (FIRST), and FreeSurfer’s aseg tool^151^. The FAST pipeline generates 139 regional grey matter volumes by segmenting T1-weighted brain images and combining these results with predefined regions of interest that cover both cortical and subcortical regions. The FIRST pipeline models and measures the shape and volume of 15 subcortical structures and outputs volumes of 15 subcortical structures. The aseg tool segments and measures the volumes of subcortical structures using T1-weighted images. In total, we used 248 sMRI traits generated from the three pipelines, including 139 IDPs processed by the FAST pipeline, 14 IDPs processed by the FIRST pipeline, and 95 IDPs processed by the aseg pipeline. We additionally included 10 global brain volume measures, both normalized for head size and non-normalized, including combined grey and white matter volume, total white matter volume, total grey matter volume, peripheral cortical grey matter volume, and ventricular cerebrospinal fluid volume. For brain dMRI, diffusion-tensor imaging (DTI) fitting was performed using the DTIFIT tool, generating DTI outputs including fractional anisotropy (FA), mean diffusivity (MD), mode of anisotropy (MO), axial diffusivity (L1), and radial diffusivities (L2 and L3). In addition to DTIFIT, voxelwise microstructural parameters, such as intra-cellular volume fraction (ICVF), isotropic volume fraction (ISOVF), and orientation dispersion index (OD), were derived using Neurite Orientation Dispersion and Density Imaging (NODDI) modelling with the AMICO tool^152,153^. These DTI and NODDI outputs were then processed using TBSS^154^, which aligns and skeletonizes the data to generate measures for 48 different white matter tracts for each DTI/NODDI output, resulting in 432 dMRI IDPs. For fMRI traits, we used 76 node amplitude traits reflecting neuronal activity and 6 global functional connectivity measures summarizing the coactivity between node pairs. These global measures were derived using a combined principal component analysis and independent component analysis approach applied to all pairwise functional connectivity traits of the 76 nodes^22,155,156^.

We analyzed three main categories of non-brain IDPs: heart and aorta, eye, and abdominal organs/tissues. For the heart and aorta, we used 76 cardiac and 6 aortic MRI traits derived from short-axis, long-axis, and aortic cine images^24,157^. The heart IDPs include global and regional metrics for four cardiac chambers: 64 traits for the left ventricle, 4 for the right ventricle, 4 for the left atrium, and 4 for the right atrium. The aortic traits consist of 3 traits for the ascending aorta and 3 for the descending aorta. For the eye, we analyzed 156 retinal imaging traits^85^, comprising 46 measures derived from optical coherence tomography (OCT) images and 110 features from fundus photographs. The derived OCT measures include retinal thickness across specific layers, vertical cup-to-disc ratio, and disc diameter. The fundus image traits were generated using 11 pre-trained transfer learning models built on ImageNet, extracting the top 10 principal components (PCs) from each model’s final layer, resulting in 110 fundus image features. Abdominal

IDPs comprised 41 MRI measures, covering body fat, body muscle, and organ-specific features. These included 8 body fat traits assessing abdominal fat distribution and visceral/subcutaneous fat volumes, and 12 body muscle traits measuring lean tissue volumes, fat-free muscle volumes in the thighs, and muscle fat infiltration as an indicator of muscle quality. Organ-specific measures included 6 kidney traits (volume and parenchyma), 10 liver traits (fat and iron content, with liver iron corrected T1 reflecting inflammation and fibrosis), 1 lung volume trait, 1 spleen volume trait, and 3 pancreas traits (volume, fat, and iron content). In total, there were 1051 IDPs across 8 imaging modalities (**Table S2**).

### UK Biobank pharma proteomics project

UK Biobank pharma proteomics project (UKB-PPP) profiled the plasma proteomes of 54,219 UKB participants using Olink Explore 3072, measuring 2,941 protein analytes representing 2,923 unique proteins across eight panels focused on inflammation, cardiometabolic health, neurology, and oncology. The project included 46,595 randomly selected participants at baseline, 6,376 consortium-selected participants, and 1,268 from the COVID-19 repeat imaging study. Protein signals were quantified as normalized protein expression values, processed using Olink’s MyData Cloud Software. Details on data collection, processing, and quality control are documented in previous study^4^.

### Phenotypic plasma protein-imaging associations

To examine the phenotypic associations between plasma proteins and IDPs, we fitted linear regression models using unrelated white British subjects^158^ as discovery samples (average *n* = 4,383). The analysis was adjusted for covariates including age (at protein assessment and imaging visit), age squared, sex, age-sex interaction, age-squared-sex interaction, top ten genetic PCs, height, weight, and body mass index. For fMRI IDPs, we additionally adjusted for volumetric scaling, head motion, and brain position^22,155^. For regional brain volumes, we additionally adjusted for total brain volume to account for global effects. *P*-values were derived from two-sided *t*-tests (R version 4.1.0) and multiple testing was corrected using Bonferroni adjustment. The analysis was replicated in an independent hold-out set of white non-British subjects (average *n* = 513). Associations were considered significant if they passed the Bonferroni threshold in the discovery sample (*P* < 1.63× 10^−8^), had a *P*-value < 0.05 in the replication sample, and exhibited concordant effect directions between the two models.

### Protein-protein interaction enrichment analysis

Protein-protein interaction (PPI) was obtained from the STRING database^39^, which integrates evidence from various interaction sources, including textmining, experiments, databases, co-expression, neighborhood, gene fusion, co-occurrence, to assign interaction scores. For a given list of proteins, the mean PPI score was calculated by averaging the pairwise PPI scores between all protein pairs. Additionally, to assess whether the observed interactions among the proteins in the list were significantly enriched, a statistical test was performed using a one-sided Wilcoxon rank-sum test. The test compared the PPI scores of the proteins in the query list against the PPI scores of all 2,923 proteins in our study, serving as the background. This analysis determined whether the query protein set exhibited significantly higher interaction compared to the overall background set of proteins.

### Pathway enrichment analysis

To identify biological pathways and functional annotations enriched among the identified proteins, we performed a pathway enrichment analysis using g:Profiler^159^. The protein list was uploaded to the g:Profiler web tool, which applies its built-in multiple testing correction method ‘g:SCS threshold’, ensuring rigorous control of false discovery rates. We selected data resources including Gene Ontology molecular function, cellular component, and biological process, as well as KEGG and Reactome pathways, to evaluate potential functional enrichment. To focus on biologically relevant terms, we restricted enriched terms to a size range of 10 to 500, ensuring that the enriched terms were neither too broad nor too narrow. This approach allowed us to identify meaningful biological pathways and processes in which the associated proteins with each organ were involved, offering deeper insights into the molecular mechanisms underlying the protein-imaging trait associations.

### Sensitivity analysis

To evaluate the potential impact of disease status on the associations between plasma proteins and IDPs, we performed a sensitivity analysis by including relevant disease statuses as covariates in the regression models. Diseases for each organ were defined based on the following ICD-10 categories: brain-related diseases (F and G), including mental and behavioral disorders, as well as diseases of the nervous system; heart and aorta-related diseases (I), encompassing ischemic heart disease, heart failure, and arrhythmias; body fat-related disorders (E65-E78), covering obesity, hyperlipidemia, and metabolic syndromes; kidney-related diseases (N00-N29), including nephrotic syndromes and chronic kidney diseases; liver-related diseases (K70-K77), such as alcoholic liver disease, nonalcoholic fatty liver disease, and cirrhosis; pancreas-related diseases (K85-K87), including acute and chronic pancreatitis; eye-related diseases (H00-H59), such as glaucoma, cataracts, and retinal disorders; spleen-related diseases (D73), including splenomegaly and other splenic disorders; and muscle-related diseases (M60-M62 and M79), including myopathies, muscle inflammation, and fibromyalgia. To ensure robust statistical power, diseases with fewer than 500 cases were excluded from the analysis. Beta coefficients and *P*-values for the associations were reported after adjusting for disease status.

### Organ-specific plasma proteins

We assigned organ-specific labels to plasma proteins following the approach described by Oh, et al. ^13^. Briefly, gene expression data were obtained from the GTEx^38^, and tissue gene expression levels were normalized. A gene was defined as organ-specific, which served as the organ label of the corresponding protein, if its expression level in one organ was at least four times higher than in any other organ, consistent with the definition proposed by the Human Protein Atlas^19^. To ensure consistency, tissues from the same organ were grouped, and the maximum expression level among sub tissues was used to represent the expression level for that organ. Following Oh, et al. ^13^, the immune organ was defined as combined gene expression in blood and spleen tissues. Using this approach, 557 out of 2,923 proteins were assigned organ-specific labels based on their gene expression profiles. To evaluate whether a list of proteins was enriched for proteins labeled with that organ based on gene expression, we performed a hypergeometric test across all organs. The test compared the number of proteins in the list with matching organ labels to the total proteins in the list, the total proteins labeled with that organ, and the total proteins analyzed, using the hypergeometric distribution to calculate the significance of enrichment.

### Prediction model using proteomic data

To predict IDPs using plasma proteins, we used an elastic-net regression model for each imaging trait. Following Carrasco-Zanini et al.^16^, we randomly sampled half of the data for training and testing, repeating this process across 200 iterations to ensure robust results. Before analysis, IDPs were normalized to allow comparability across different traits, while protein predictors data were also standardized within the elastic-net model. During each iteration, elastic-net regression was performed, and coefficients for each protein were estimated using the optimal lambda determined through cross-validation. Predicted IDP values for the testing data were then calculated based on these estimated coefficients, and prediction performance was assessed using the *R*-squared metric. After the 200 repetitions, absolute values of coefficients across the 200 repetitions were summed up, as an importance measure of each protein for IDPs. GLIPR1 was excluded from the prediction model due to its low data quality as described in Sun, et al. ^4^. To determine the top five predictors for each IDP, we calculated a combined score for each protein by integrating the absolute sum of coefficients and the frequency of selection across iterations. Both metrics were first normalized to a [0, 1] scale by dividing each value by the maximum value in its respective column. A weighted sum of the normalized metrics was then computed for each protein, with equal weights (0.5) assigned to the normalized coefficient sum and frequency. Proteins were ranked in descending order of their combined scores, and the top five with the highest scores were selected as the strongest predictors for the IDP.

### Mendelian randomization and colocalization

We conducted Mendelian randomization (MR) analysis to investigate the genetic causal links between plasma proteins and IDPs. The list of proteins and IDPs used in the MR analysis was largely the same as in previous sections, with minor adjustments due to data availability or data filtering criteria. For IDPs, genome-wide association study (GWAS) summary statistics were obtained from previous studies^85,111,157,160^. Ten global brain volume measures were excluded due to data unavailability, while the remaining 1,041 IDPs were included in the MR analysis (average *n* = 40,682 participants). Detailed information on these GWAS, including data preprocessing and quality control procedures, is available in the respective studies. For plasma proteins, 67 out of the 2,923 plasma proteins were excluded based on the following three criteria: First, fusion proteins which represent combined entities or closely related paralogs rather than distinct proteins, were excluded to avoid ambiguity in biological interpretation. Second, proteins encoded by genes located on the sex chromosomes were removed to account for sex-specific differences in expression. Finally, proteins that could not be mapped to genomic locations in the GRCh37/hg19 genome build were excluded. Detailed description of these exclusions is available in the previous study^32^. Additionally, 36 proteins located within the MHC region were excluded to minimize bias from high linkage disequilibrium (LD) in this genomic region, leaving 2,820 proteins for the MR analysis. To prevent overlap with subjects in the imaging GWAS, we generated protein quantitative trait loci (pQTL) specifically for this MR analysis, ensuring that participants (and their relatives^158^) from the imaging GWAS were excluded from the pQTL analysis (average *n* = 34,566 participants). We removed the effects of the same set of covariates as in the previous study^32^. In addition, only *cis*-pQTLs, defined as genetic variants located within a gene’s region and extending one megabase on both sides, were used in the MR analysis. To generate independent instrumental variables, *cis*-pQTLs were clumped using PLINK 2.0^161^, with a *P*-value threshold of 5× 10^−8^, a window size of 250 kb, and an *r²* threshold of 0.001 for LD. Additionally, variants with an *F*-statistic less than 10 were excluded to avoid weak instrument bias.

The MR methods used in the study were the inverse variance weighted (IVW) and the Wald-ratio (for single instrument variant). The Wald-ratio was conducted with the TwoSampleMR^113,114^ package, while the IVW method was implemented with the MendelianRandomization^112^ package. Following the pipeline described by Zheng, et al. ^115^, we used a range of sensitivity tests to ensure the robustness of our results. Specifically, the Steiger *P*-value evaluated whether the genetic instruments explained more variance in the exposure (plasma proteins) than in the outcome (IDPs), confirming the correct causal direction. Additionally, the heterogeneity *P*-value assessed the consistency of causal estimates across genetic instruments, with significant heterogeneity indicating potential violations of MR assumptions. To account for multiple testing, a Bonferroni correction was applied after filtering out results that failed sensitivity tests, resulting in an adjusted *P*-value threshold of *P* < 2.98× 10^−8^ and 8,116 significant causal protein-imaging trait associations.

To further assess whether the significant MR pairs shared a causal variant, we performed colocalization analysis on the 8,116 pairs with significant causal relationships. We applied the coloc.abf() method from the coloc package^33^, a Bayesian framework that calculates five posterior probabilities: PPH0 (no association), PPH1 (association with the protein only), PPH2 (association with the imaging trait only), PPH3 (independent associations with both traits), and PPH4 (a shared causal variant). Pairs with PPH4 > 80% were considered colocalized, providing strong evidence for a shared genetic basis between the plasma protein and IDP.

## Code availability

We made use of publicly available software and tools. The code used in this study will be deposited in Zenodo upon publication.

## Data availability

The individual-level data used in this study can be obtained from UK Biobank (https://www.ukbiobank.ac.uk/). The GWAS summary statistics for imaging phenotypes across different organs are available from previous studies^85,111,157,160^. The GWAS summary statistics of Olink plasma proteins (removing imaging subjects) produced in this study will be deposited in Zenodo upon publication. Other datasets in this paper include: the STRING database (https://string-db.org/), the Therapeutic Target Database (https://idrblab.net/ttd/), and the GTEx dataset v8 (https://gtexportal.org/).

