## Supplementary material for "The landscape of plasma proteomic links to human organ imaging": supp_information

**This PDF file includes:**

Supplementary Text
Supplementary Figures Figs. S1-S75

**Other Supplementary Materials for this manuscript include the following:**

Tables S1 to S13 (.xlsx) (available in a zip file)

### Supplementary Text

#### Additional results on putative origins of proteomic effects on imaging traits

Similar to the brain, liver, and pancreas, we found that proteins associated with spleen, body fat, and lung were highly expressed in the same organ (**Table S7** and **Figs. 3F-H**). Sixteen proteins associated with spleen volume were highly expressed in the spleen or immune system, reflecting their critical roles in immune function (enrichment $P$ = $1.56\times 10^{-6}$). For example, NCR1 and CD244, primarily expressed in natural killer (NK) cells, regulate cytotoxicity[1,2]. Similarly, FASLG, primarily expressed in antigen-stimulated T lymphocytes and NK cells, is a member of tumor necrosis factor receptor family and induces apoptosis upon binding with its receptor[3,4]. Additionally, CD8A, as part of the CD8 co-receptor expressed on cytotoxic T cells, plays a pivotal role in anticancer immunity and serves as a foundation for modern cancer immunotherapies[5]. Proteins associated with body fat also demonstrated similar organ-specific expression patterns (enrichment $P$ = $2.01\times 10^{-6}$). Four body fat-associated proteins, FABP4, LEP, ADIPOQ, and CD300LG, were highly expressed in adipose tissue and play critical roles in lipid metabolism, insulin sensitivity, and energy homeostasis. ADIPOQ, secreted from adipocytes, promotes adipocyte proliferation, differentiation, lipid accumulation, and insulin responsiveness[6]. FABP4 regulates lipid metabolism and insulin resistance, contributing to conditions such as obesity, diabetes, hypertension, and atherosclerosis[7]. LEP, primarily secreted by adipose tissue, is essential for adipocyte metabolism and energy balance[8]. A similar pattern was observed in the lung as well (enrichment $P$ = $6.95\times 10^{-4}$). Among the 11 proteins associated with lung volume, SCGB3A2 was highly expressed in the lung. SCGB3A2 acts as a growth factor in lung development, promoting both early organogenesis and late-stage maturation[9].

#### Additional results on prediction performance of plasma protein for imaging traits

For brain dMRI traits, plasma proteins showed slightly reduced predictive power compared to brain sMRI traits ($r^2$ < 0.04). GFAP not only demonstrated extensive phenotypic associations with dMRI traits but also served as a key predictor for white matter integrity in tracts such as the cingulum hippocampus, sagittal stratum, external capsule, and anterior corona radiata (**Figs. S41-S44**). In addition, GDF15 and LEP, which were strong predictors for cerebellum volumes, also predicted white matter microstructure of the superior cerebellar peduncle (**Fig. S45**). This alignment is consistent with the close relationship between the superior cerebellar peduncle and the cerebellum, as the peduncle connects the cerebellum to other parts of the brain, sharing the same predictors. Moreover, APCS predicted the white matter integrity of the pontine crossing fibers (**Fig. S46**). APCS, produced exclusively in the liver, has been linked to a higher risk of neurodegenerative diseases such as Alzheimer's disease when it appears in the brain due to compromised blood-brain barrier integrity. APCS is cytotoxic to cerebral neurons, promotes Aβ amyloid formation, highlighting an inter-organ relationship in disease pathology[1,2]. For the eye, the predictive power of plasma proteins was primarily focused on the average thickness of ganglion cell-inner plexiform layer and retinal nerve fiber layer ($r^2$ range = (0.02, 0.03), **Fig. S47**), aligning with findings from phenotypic associations. Top predictors for these IDPs included OMG and CCL2.

**Additional examples of genetic-root causal links between plasma proteins and imaging traits**

Proteins uniquely associated with dMRI included BCAN and OMG, both of which also exhibited phenotypic associations with dMRI. BCAN was causally associated with the reduced structural integrity of multiple white matter tracts, including the cingulum hippocampus, corpus callosum, internal capsule, anterior corona radiata, and superior corona radiata (β range = (-0.37, -0.26), $P < 5.02\times 10^{-9}$, PPH4 range = (99.28%, 100%), **Figs. 6A** and **S60-S63**). BCAN is a critical biomarker for brain aging and is implicated in brain disorders such as dementia and stroke[3,4]. In contrast, increased levels of OMG displayed positive causal associations with enhanced white matter integrity in the anterior and posterior corona radiata (β range = (0.42, 0.45), $P < 2.01\times 10^{-8}$, PPH4 range = (98.92%, 99.18%), **Fig. S64**). In addition to proteins overlapping with phenotypic associations, other proteins such as FOXO1 and PCDH7 were identified. Elevated FOXO1 levels showed causal associations with increased white matter integrity in the inferior cerebellar peduncle (β = 0.20, $P = 3.73\times 10^{-11}$, **Fig. S65**), while increased level of PCDH7 was causally associated with decreased white matter integrity in the fornix cres and stria terminalis (β range = (-0.32, -0.33), $P < 8.57\times 10^{-11}$, PPH4 range = (83.84%,

95.66%), **Fig. S66**). PCDH7 is essential for brain development as a neuronal adhesion molecule[5], while FOXO1 is implicated in mood and anxiety regulation[6].

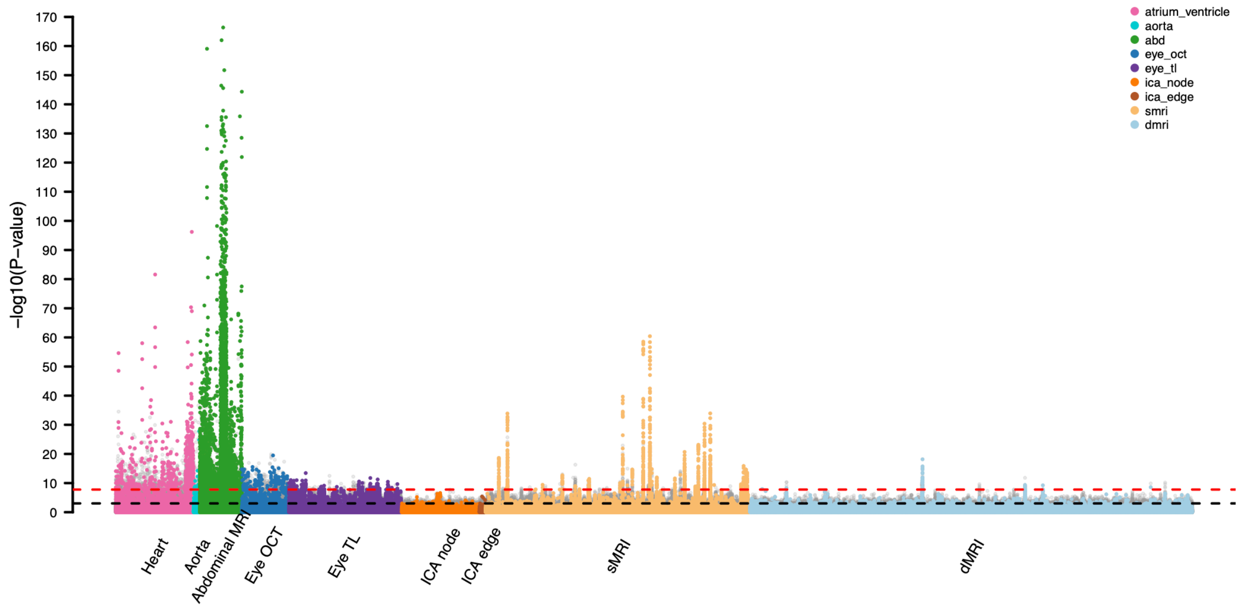


**Fig. S1 Overview of phenotypic associations between plasma proteins and multi-organ imaging-derived phenotypes (IDPs).**

The -log10(*P*-value) between 2,923 plasma proteins and multi-organ IDPs, including 258 brain structural MRI (sMRI) traits, 432 brain diffusion MRI (dMRI) traits, 82 brain functional MRI traits (76 ICA nodes and 6 ICA edges), 82 cardiac (atrium and ventricle) and aortic MRI traits, 41 abdominal (abd) MRI traits, 46 eye optical coherence tomography (OCT) traits, and 110 eye fundus imaging features (eye tl). Associations replicated in an independent hold-out dataset are highlighted. A red dashed line represents the threshold from Bonferroni correction ($P < 1.63\times 10^{-8}$), and a black dashed line represents threshold from 5% false discovery rate correction ($P < 9.33\times 10^{-4}$). See Table S1 for more information on plasma proteins and Table S2 on multi-organ IDPs.

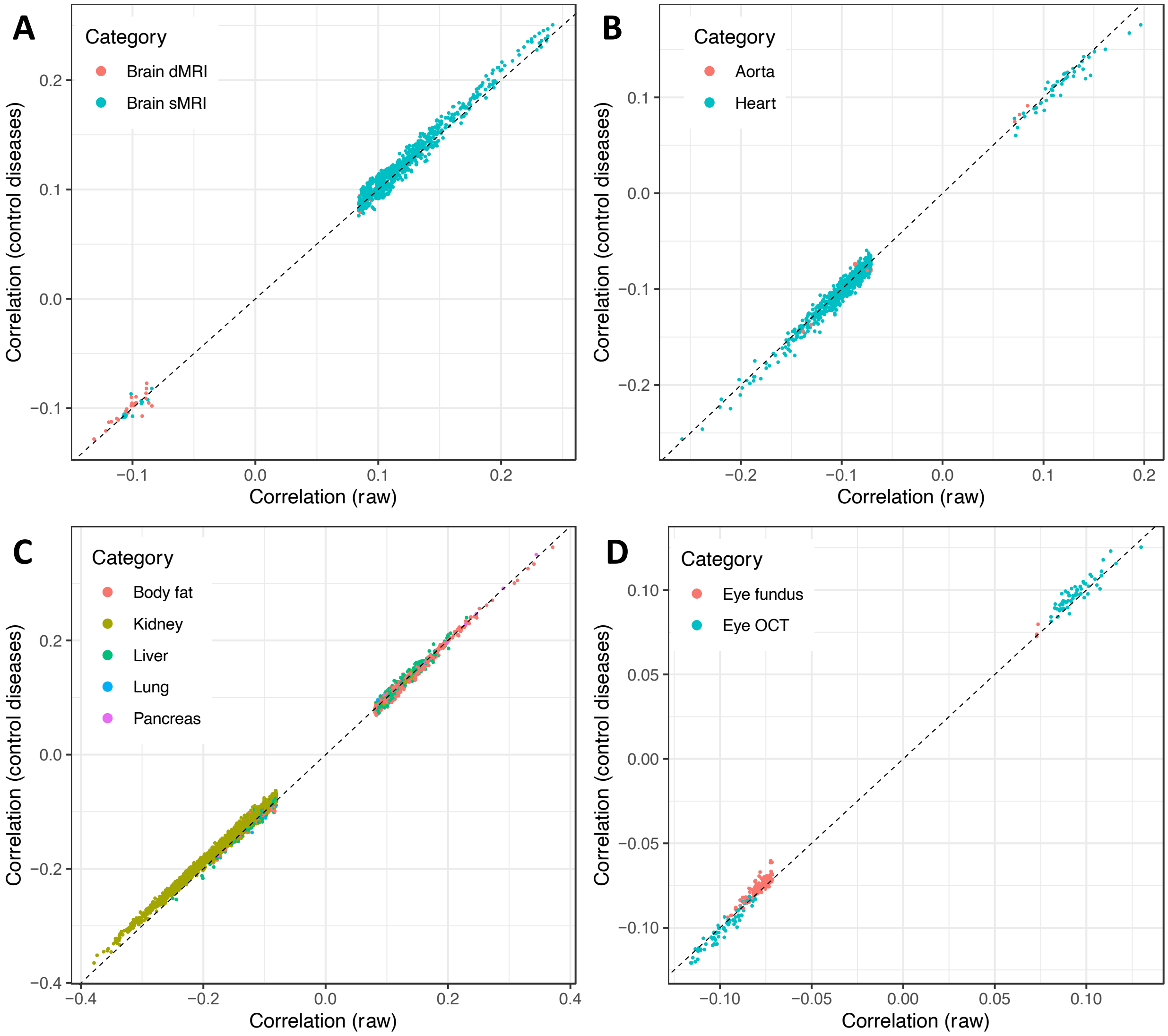


**Fig. S2 Sensitivity check for protein-imaging associations before and after adjusting for disease status.**

The estimated coefficients for protein-imaging associations from models without (*x*-axis) and with (*y*-axis) additional adjustment for relevant disease status. Each point represents a protein-imaging trait pair, with panels representing imaging traits from different organs: **A** brain, **B** heart and aorta, **C** abdomen, and **D** eye. Only pairs that passed Bonferroni correction ($P < 1.63\times 10^{-8}$) and were replicated in an independent hold-out sample were plotted.

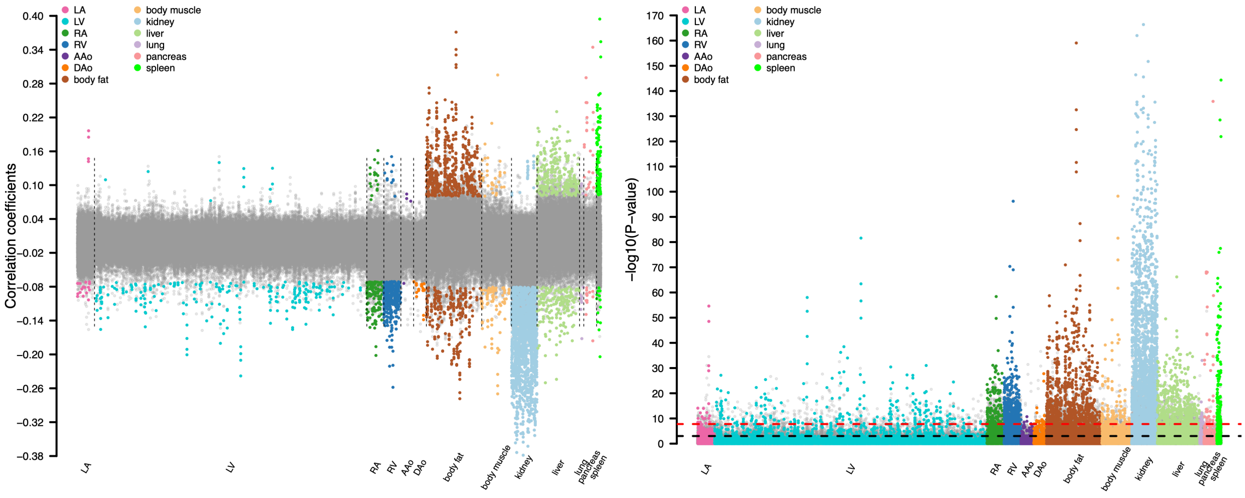


**Fig. S3 Phenotypic associations between plasma proteins and non-brain imaging-derived phenotypes (IDPs).**

The -log10(*P*-value) and correlation coefficients between 2,923 plasma proteins and heart and abdominal IDPs, including 82 cardiac and aortic MRI traits (including six sub-categories: left ventricle [LV], right ventricle [RV], left atrium [LA], right atrium [RA], ascending aorta [AAo], and descending aorta [DAo]), 8 body fat traits, 12 body muscle traits, 6 kidney traits, 10 liver traits, 1 lung trait, 3 pancreas traits, and 1 spleen trait. Associations survived the Bonferroni correction ($P < 1.63\times 10^{-8}$) and were replicated in an independent hold-out dataset were highlighted with colors. A red dashed line represents the threshold from Bonferroni correction ($P < 1.63\times 10^{-8}$), and a black dashed line represents threshold from 5% false discovery rate correction ($P < 9.33\times 10^{-4}$). See Table S1 for more information on plasma proteins and Table S2 on multi-organ IDPs.

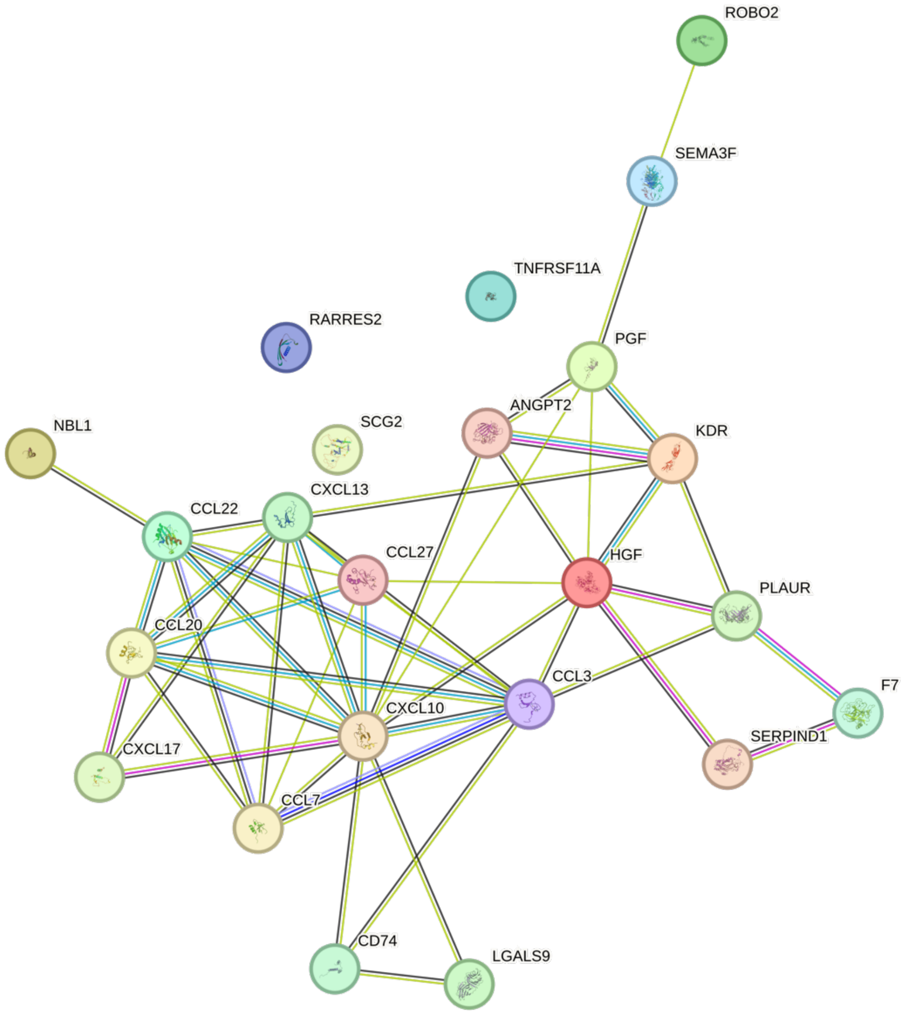


**Fig. S4 Example of protein-protein interaction (PPI) network of specific proteins associated with heart imaging-derived phenotypes (IDPs).**

PPI network of proteins associated with heart IDPs that enriched in chemotaxis and taxis. Colored edges between the protein pairs represent different interaction sources on the STRING database, including curated databases, experimental evidence, gene neighborhood, gene fusions, gene co-occurrence, text mining, co-expression, and protein homology. Only high-confidence interactions (PPI score > 0.7) are shown.

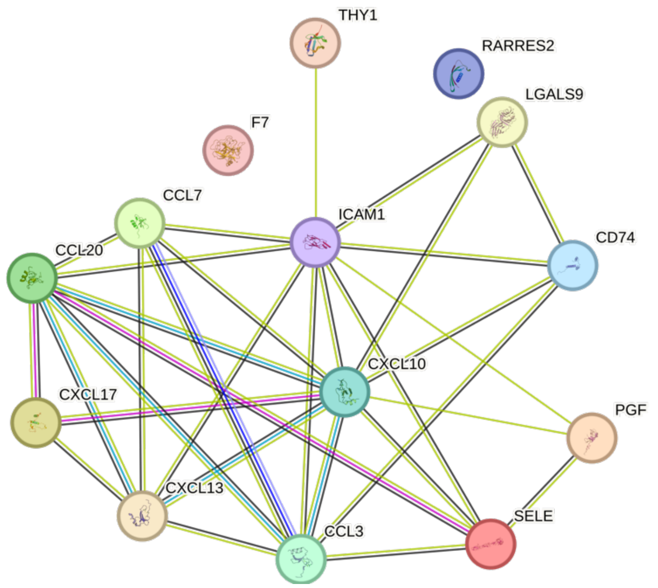


**Fig. S5 Example of protein-protein interaction (PPI) network of specific proteins associated with heart imaging-derived phenotypes (IDPs).**

PPI network of proteins associated with heart IDPs that enriched in the positive regulation of leukocyte migration. Colored edges between the protein pairs represent different interaction sources on the STRING database, including curated databases, experimental evidence, gene neighborhood, gene fusions, gene co-occurrence, text mining, co-expression, and protein homology. Only high-confidence interactions (PPI score > 0.7) are shown.

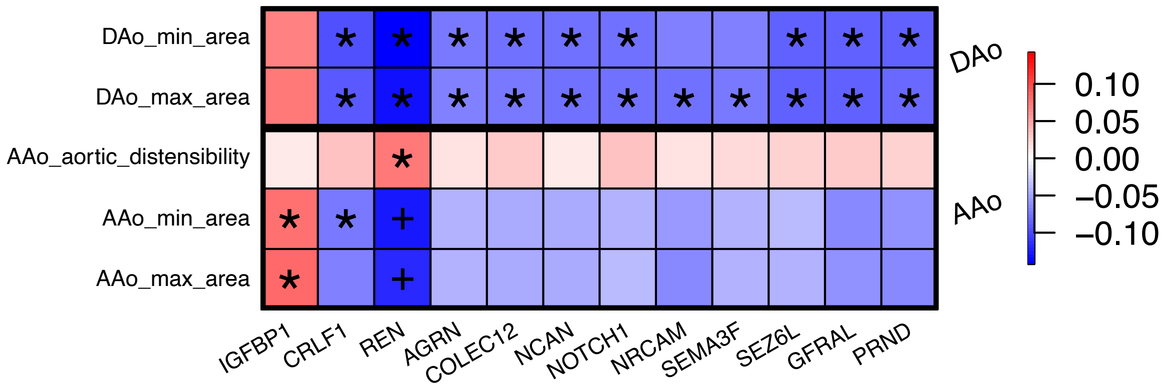


**Fig. S6 Phenotypic associations between plasma proteins and aortic imaging-derived phenotypes (IDPs).**

We illustrate the correlation coefficients between plasma proteins (*x*-axis) and aortic IDPs (*y*-axis). The color represents correlation estimates. The coefficients that passed Bonferroni correction ($P < 1.63\times 10^{-8}$) and were replicated in an independent hold-out dataset were marked with asterisk. The coefficients passed Bonferroni multiple testing but were not replicated in our approach are marked with plus sign.

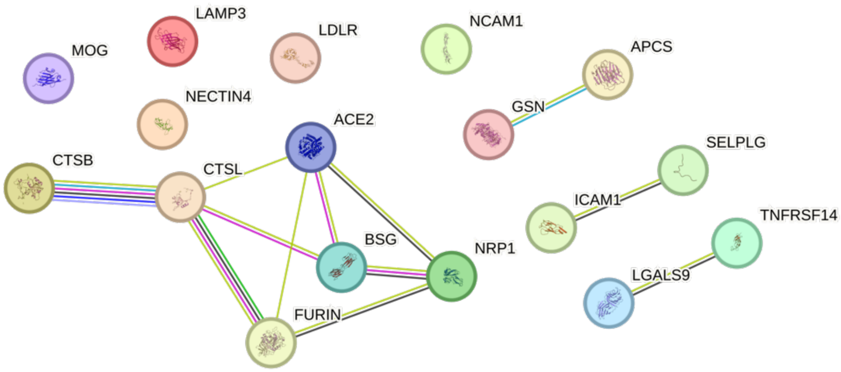


**Fig. S7 Example of protein-protein interaction (PPI) network of specific proteins associated with liver imaging-derived phenotypes (IDPs).**

PPI network of proteins associated with live IDPs that enriched in the viral life cycle pathway. Colored edges between the protein pairs represent different interaction sources on the STRING database, including curated databases, experimental evidence, gene neighborhood, gene fusions, gene co-occurrence, text mining, co-expression, and protein homology. Only high-confidence interactions (PPI score > 0.7) are shown.

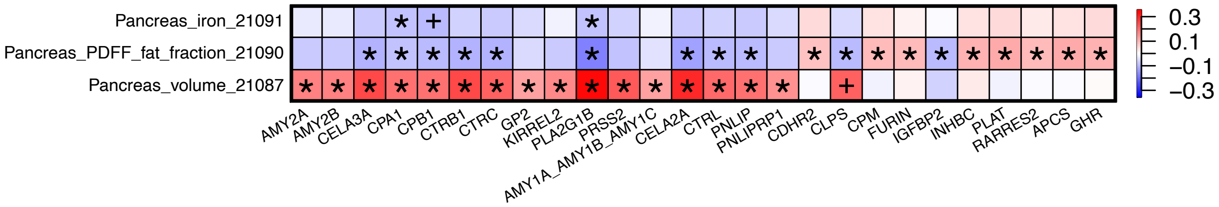


**Fig. S8 Phenotypic associations between plasma proteins and pancreas imaging-derived phenotypes (IDPs).**

We illustrate the correlation coefficients between pancreas IDPs (*y*-axis) and plasma proteins (*x*-axis). The color represents correlation estimates. The coefficients that passed Bonferroni correction ($P$ < $1.63\times 10^{-8}$) and were replicated in an independent hold-out dataset were marked with asterisk. The coefficients passed Bonferroni multiple testing but were not replicated in our approach are marked with plus sign.

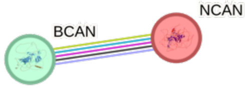


**Fig. S9 Strong protein-protein interaction between NCAN and BCAN.**

Two proteins associated with brain structural MRI, NCAN and BCAN, have a strong protein-protein interaction (PPI score = 0.96) on the STRING database. Colored edges between the protein pairs represent different interaction sources, including curated databases, experimental evidence, gene neighborhood, gene fusions, gene co-occurrence, text mining, co-expression, and protein homology.

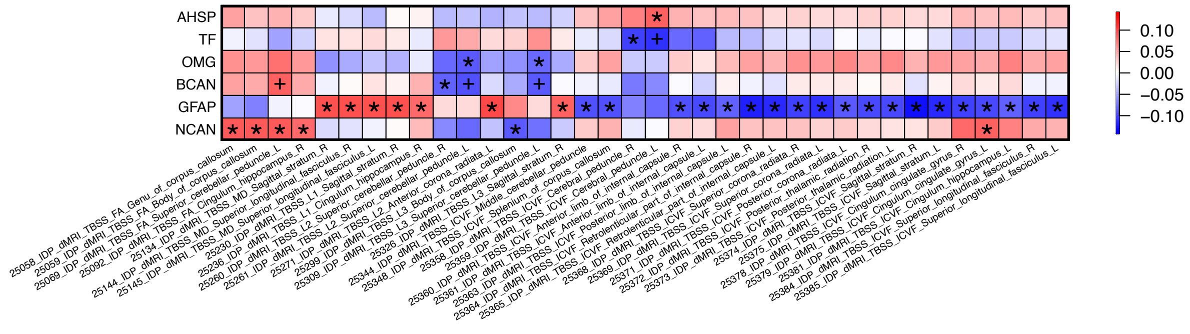


**Fig. S10 Phenotypic associations between plasma proteins and brain diffusion MRI (dMRI) imaging-derived phenotypes (IDPs).**

We illustrate the correlation coefficients between plasma proteins (*y*-axis) and brain dMRI IDPs (*x*-axis). The color represents correlation estimates. The coefficients that passed Bonferroni correction ($P < 1.63\times 10^{-8}$) and were replicated in an independent hold-out dataset were marked with asterisk. The coefficients passed Bonferroni multiple testing but were not replicated in our approach are marked with plus sign.

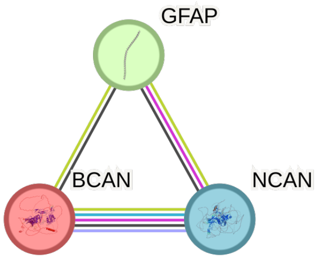


**Fig. S11 Example of protein-protein interaction (PPI) network of specific proteins associated with brain imaging-derived phenotypes (IDPs).**

PPI network of proteins associated with brain IDPs that enriched in positive regulation of neurogenesis and nervous system development, glial cell differentiation, and positive regulation of neuroblast proliferation pathways. Colored edges between the protein pairs represent different interaction sources on the STRING database, including curated databases, experimental evidence, gene neighborhood, gene fusions, gene co-occurrence, text mining, co-expression, and protein homology. Only high-confidence interactions (PPI score > 0.7) are shown.

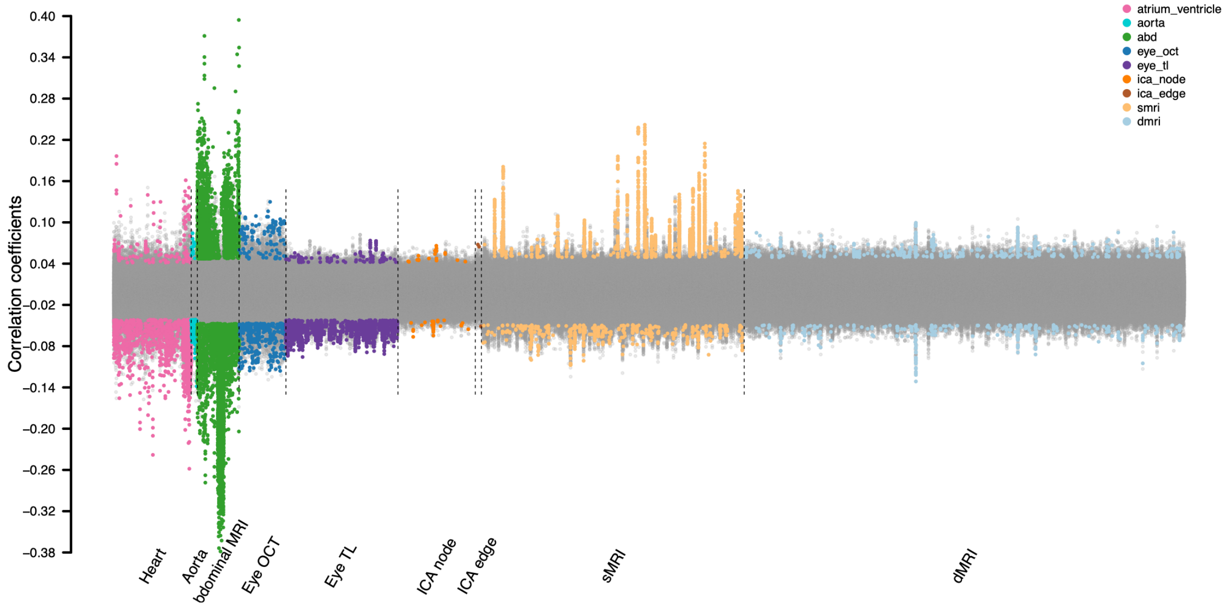


**Fig. S12 Phenotypic associations between plasma proteins and multi-organ imaging-derived phenotypes (IDPs) under false discovery rate (FDR) correction.**

The correlation coefficients between 2,923 plasma proteins and multi-organ IDPs, including 258 brain structural MRI (sMRI) traits, 432 brain diffusion MRI (dMRI) traits, 82 brain functional MRI traits (76 ICA nodes and 6 ICA edges), 82 cardiac (atrium and ventricle) and aortic MRI traits, 41 abdominal (abd) MRI traits, 46 eye optical coherence tomography (OCT) traits, and 110 eye fundus imaging features (eye tl). Associations survived the FDR rate of 5% ($P < 9.33\times 10^{-4}$) and were replicated in an independent hold-out dataset were highlighted with colors. See Table S1 for more information on plasma proteins and Table S2 on multi-organ IDPs.

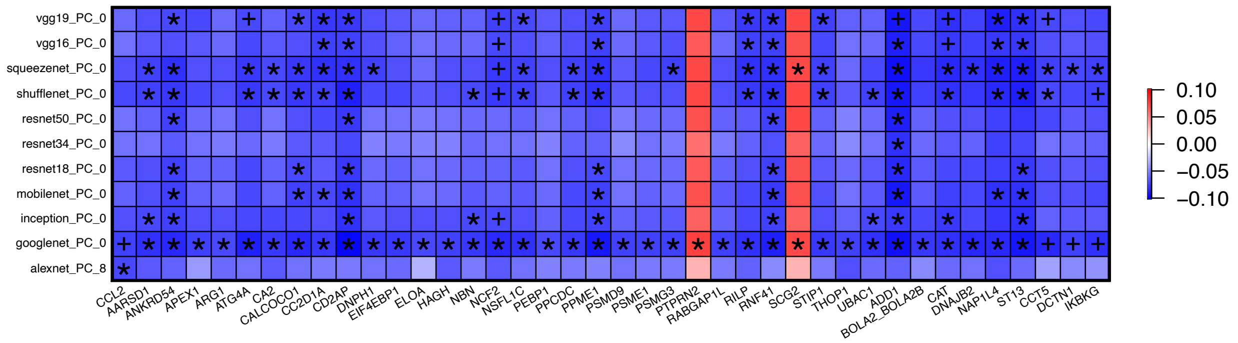


**Fig. S13 Phenotypic associations between plasma proteins and eye fundus imaging-derived phenotypes (IDPs).**

We illustrate the correlation coefficients between plasma proteins (*x*-axis) and eye fundus IDPs (*y*-axis). The color represents correlation estimates. The coefficients that passed Bonferroni correction ($P < 1.63\times 10^{-8}$) and were replicated in an independent hold-out dataset were marked with asterisk. The coefficients passed Bonferroni multiple testing but were not replicated in our approach are marked with plus sign.

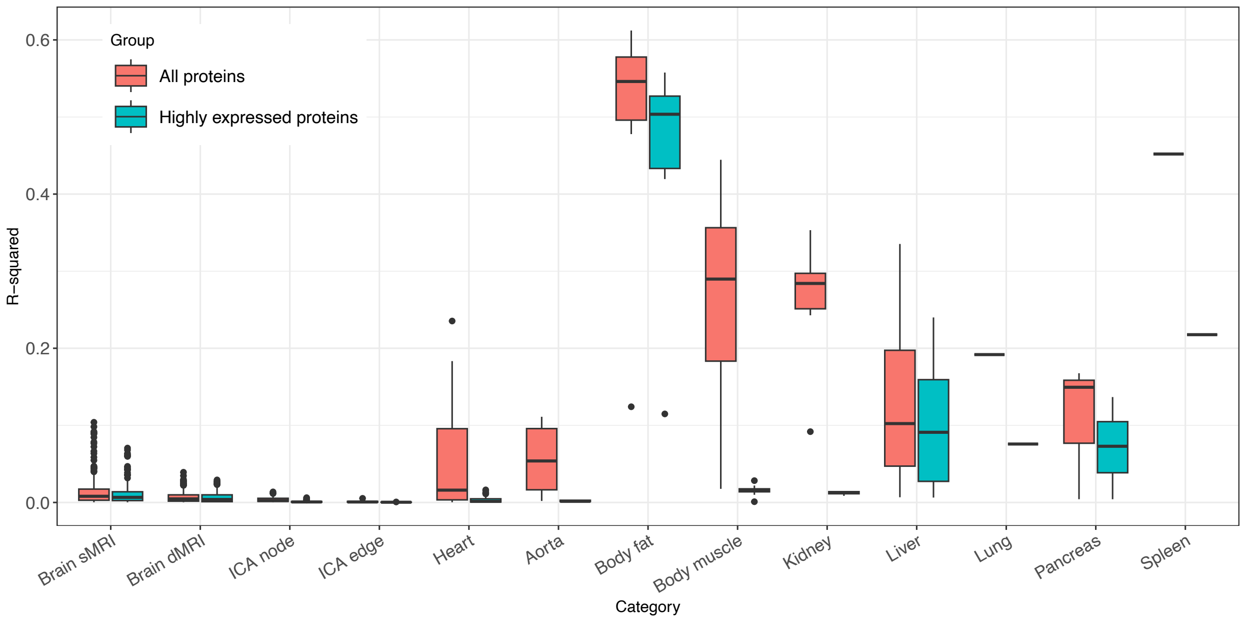


**Fig. S14 Predictive $R^2$ of all plasma proteins and organ-specific highly expressed proteins for each organ.**

The mean prediction $R^2$ (*y*-axis) for each organ (*x*-axis), comparing predictions using all plasma proteins versus only proteins highly expressed in the respective organ. Colors indicate whether the predictions were based on all proteins or only highly expressed proteins.

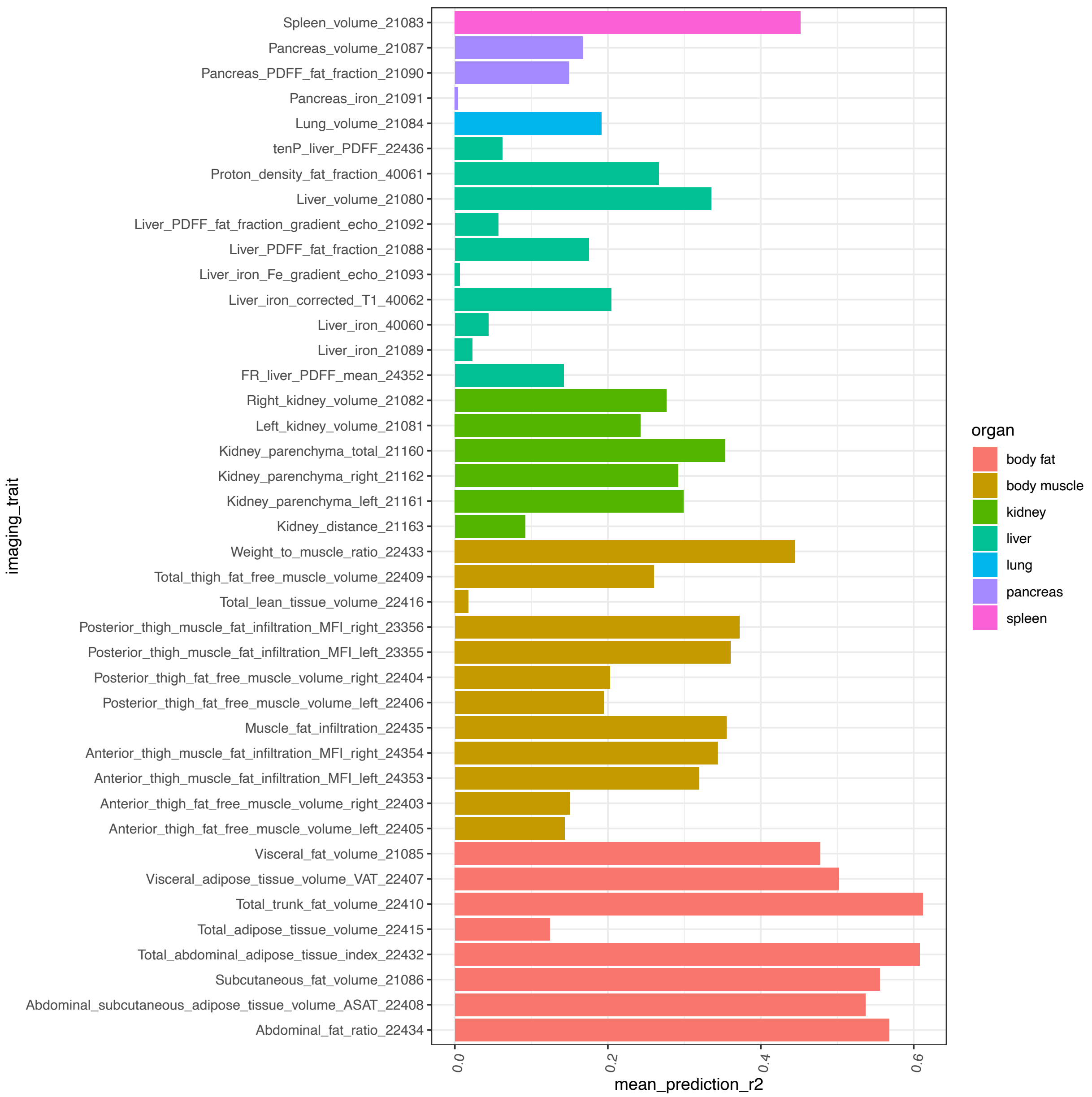


**Fig. S15 Predictive $R^2$ of abdominal imaging-derived phenotypes (IDPs).**

The mean prediction $R^2$ (*x*-axis) for abdominal IDPs (*y*-axis) across 200 resampling iterations. Colors represent the organs associated with each abdominal IDP.

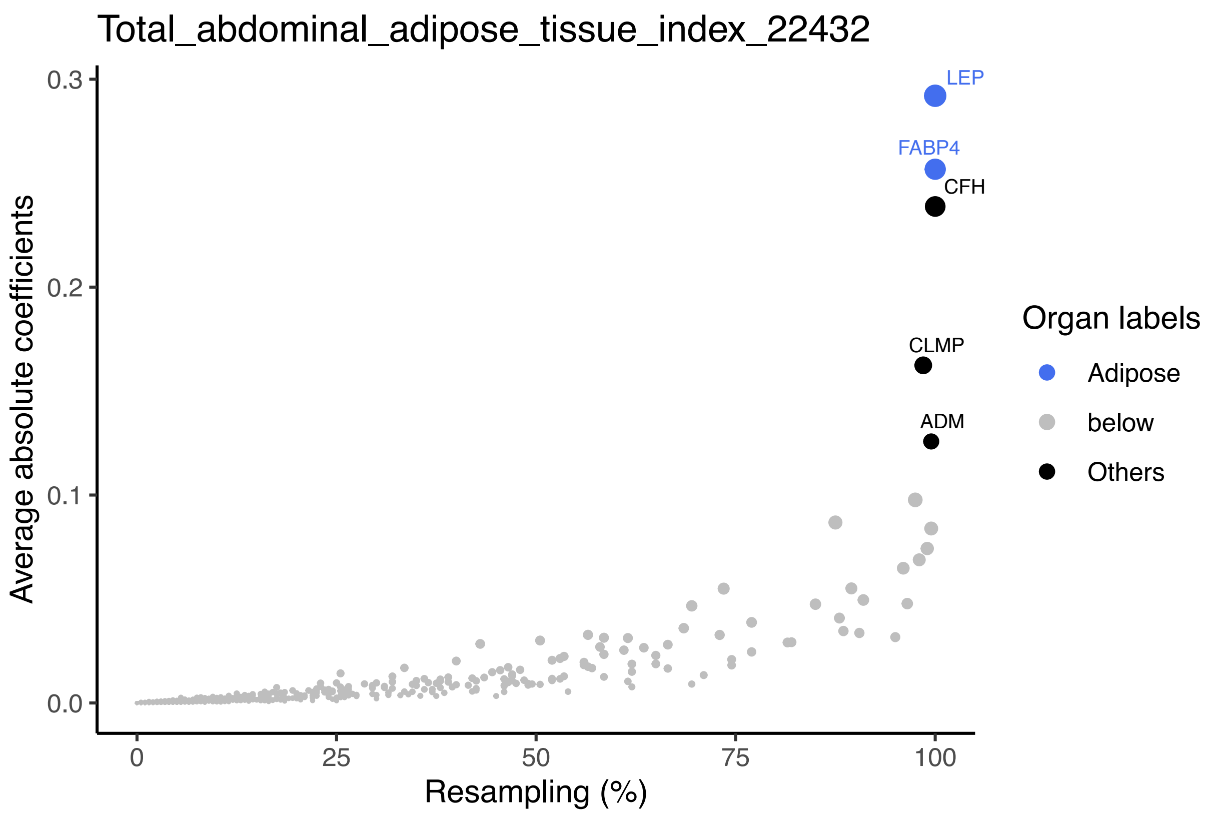


**Fig. S16 Top-ranked plasma protein predictors for abdominal MRI.**

Predictive power of plasma proteins for total abdominal adipose tissue index. The *x*-axis represents the frequency of proteins appearing across 200 resampling iterations, while the *y*-axis represents the average absolute value of coefficients across the 200 resampling iterations. Top proteins are labeled with their names and highlighted in color, with the color indicating the organ where the protein is most highly expressed (at least 4 times higher than any other organs).

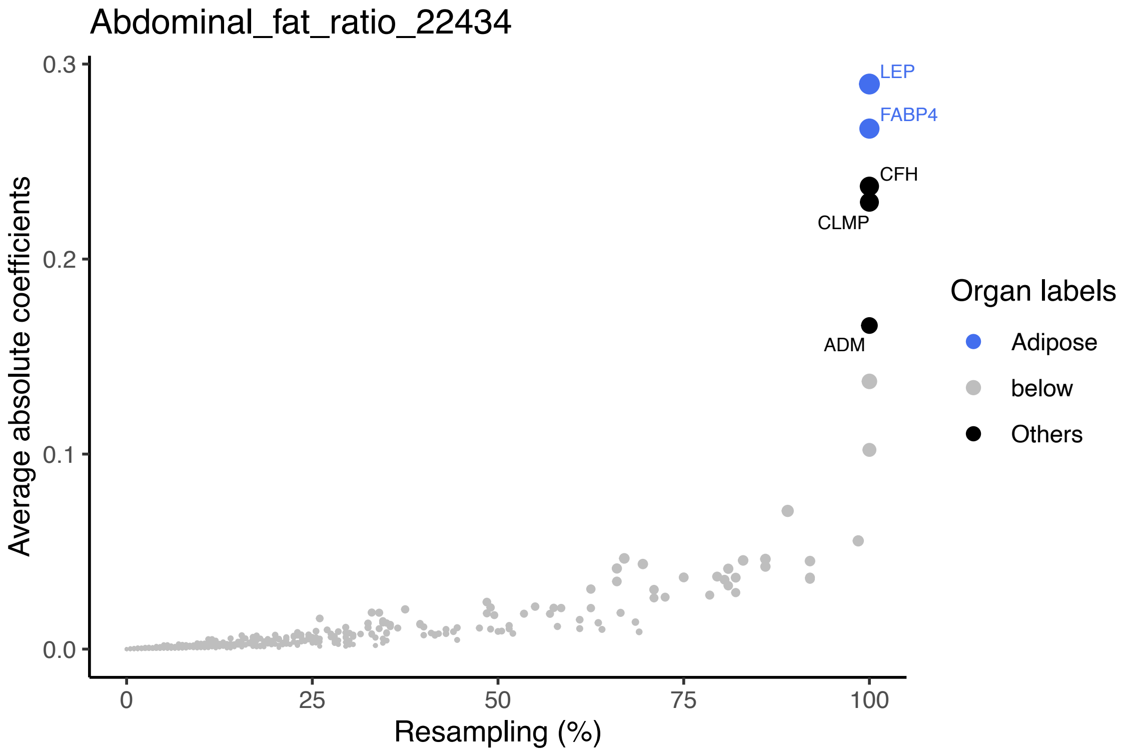


**Fig. S17 Top-ranked plasma protein predictors for abdominal MRI.**

Predictive power of plasma proteins for abdominal fat ratio. The *x*-axis represents the frequency of proteins appearing across 200 resampling iterations, while the *y*-axis represents the average absolute value of coefficients across the 200 resampling iterations. Top proteins are labeled with their names and highlighted in color, with the color indicating the organ where the protein is most highly expressed (at least 4 times higher than any other organs).

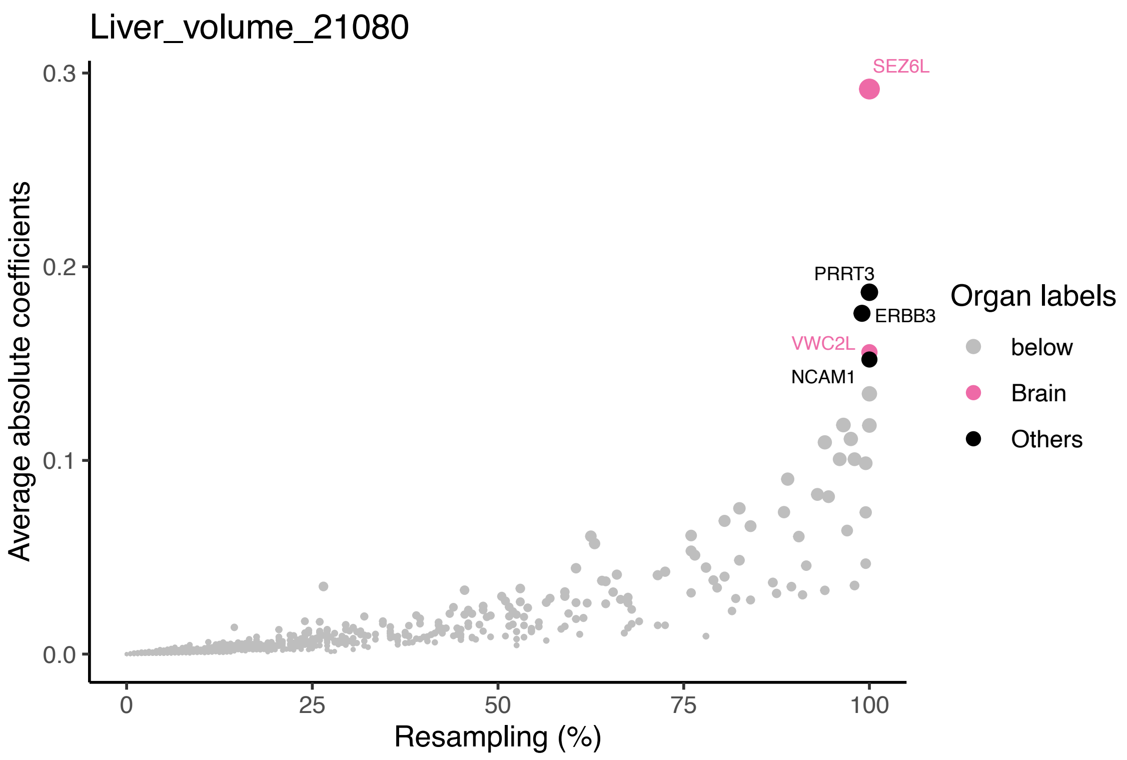


**Fig. S18 Top-ranked plasma protein predictors for abdominal MRI.**

Predictive power of plasma proteins for liver volume. The *x*-axis represents the frequency of proteins appearing across 200 resampling iterations, while the *y*-axis represents the average absolute value of coefficients across the 200 resampling iterations. Top proteins are labeled with their names and highlighted in color, with the color indicating the organ where the protein is most highly expressed (at least 4 times higher than any other organs).

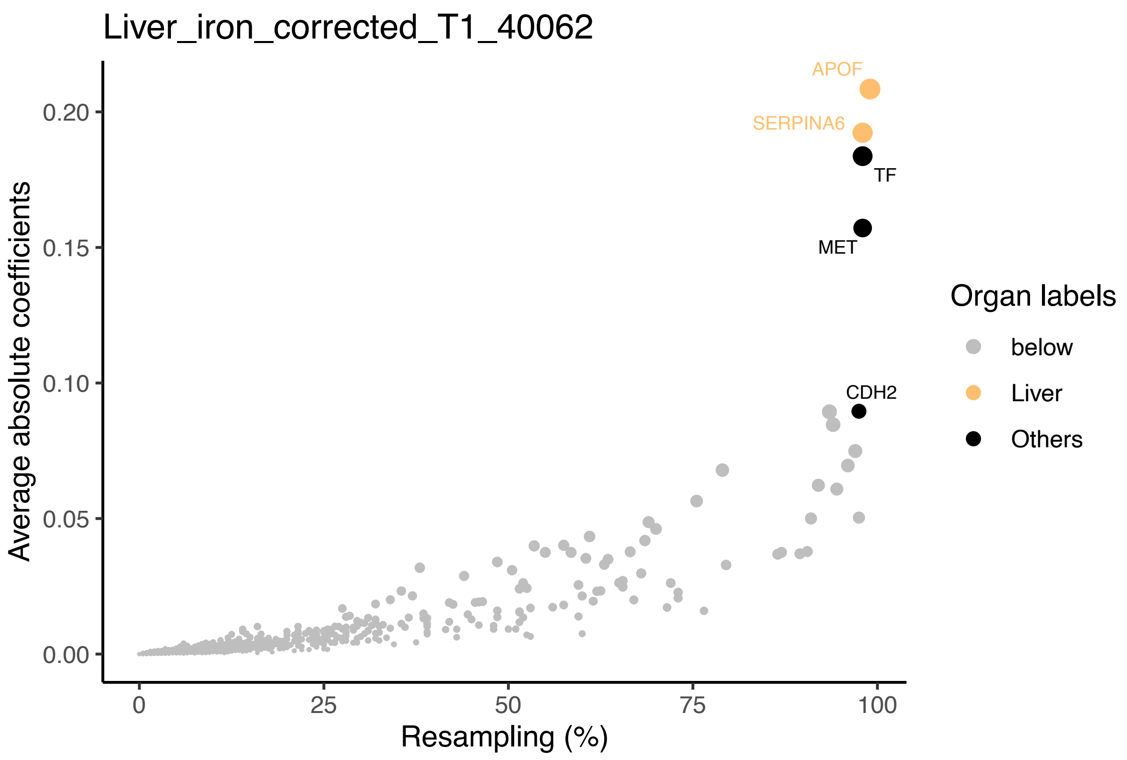


**Fig. S19 Top-ranked plasma protein predictors for abdominal MRI.**

Predictive power of plasma proteins for liver iron corrected T1. The *x*-axis represents the frequency of proteins appearing across 200 resampling iterations, while the *y*-axis represents the average absolute value of coefficients across the 200 resampling iterations. Top proteins are labeled with their names and highlighted in color, with the color indicating the organ where the protein is most highly expressed (at least 4 times higher than any other organs).

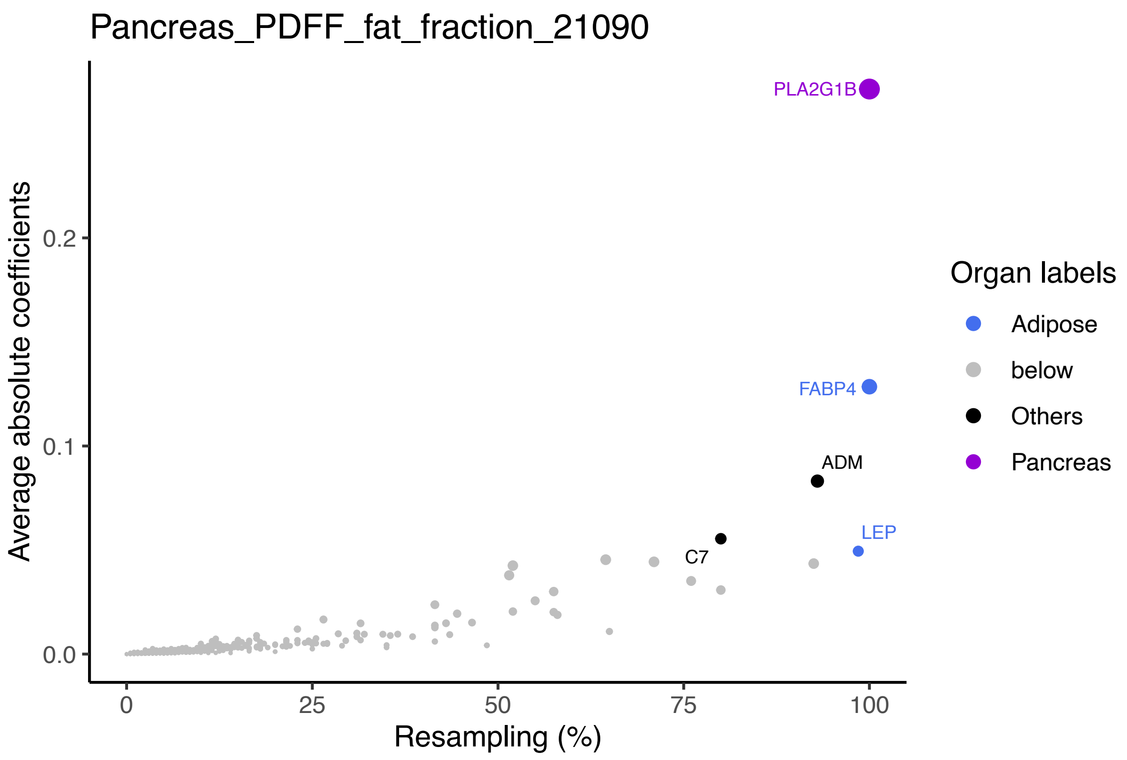


**Fig. S20 Top-ranked plasma protein predictors for abdominal MRI.**

Predictive power of plasma proteins for pancreas fat fraction. The *x*-axis represents the frequency of proteins appearing across 200 resampling iterations, while the *y*-axis represents the average absolute value of coefficients across the 200 resampling iterations. Top proteins are labeled with their names and highlighted in color, with the color indicating the organ where the protein is most highly expressed (at least 4 times higher than any other organs).

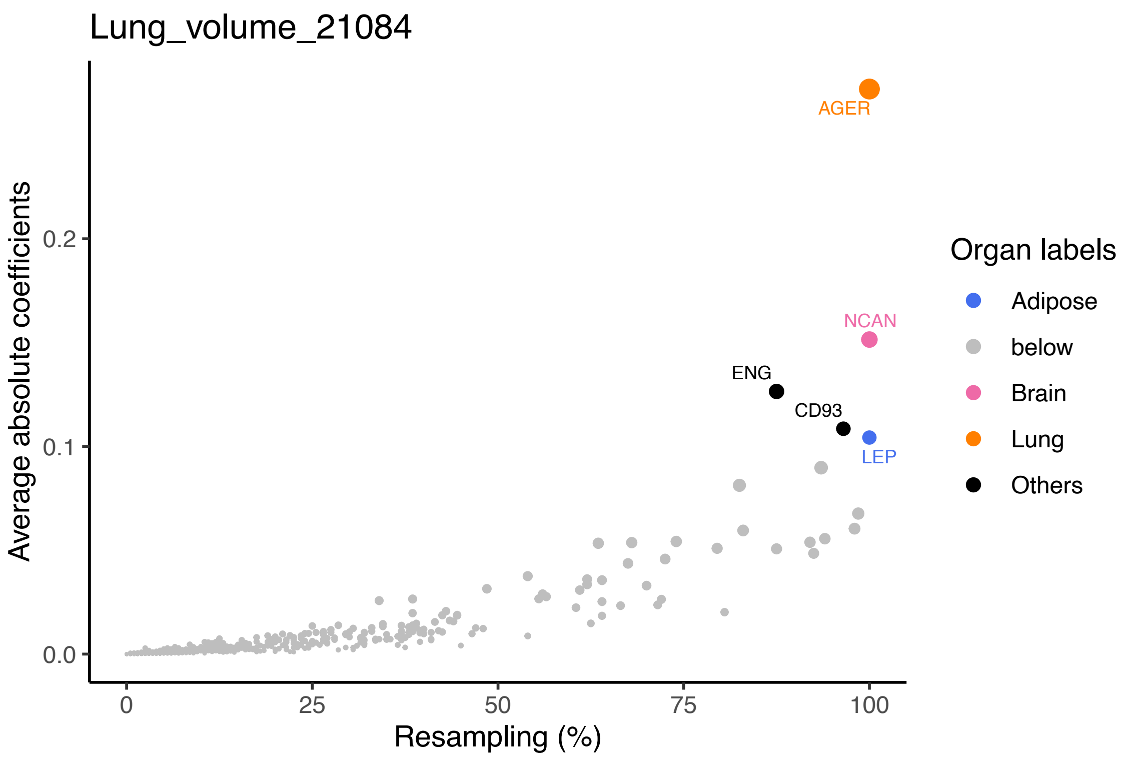


**Fig. S21 Top-ranked plasma protein predictors for abdominal MRI.**

Predictive power of plasma proteins for lung volume. The *x*-axis represents the frequency of proteins appearing across 200 resampling iterations, while the *y*-axis represents the average absolute value of coefficients across the 200 resampling iterations. Top proteins are labeled with their names and highlighted in color, with the color indicating the organ where the protein is most highly expressed (at least 4 times higher than any other organs).

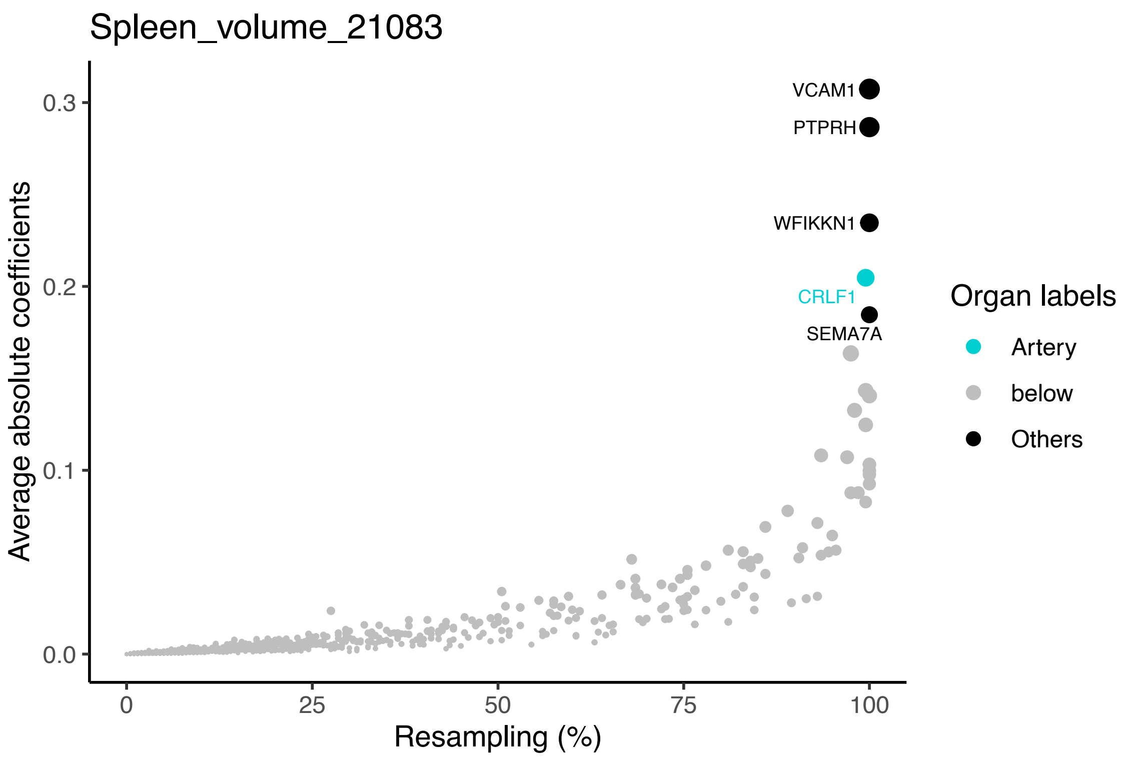


**Fig. S22 Top-ranked plasma protein predictors for abdominal MRI.**

Predictive power of plasma proteins for spleen volume. The *x*-axis represents the frequency of proteins appearing across 200 resampling iterations, while the *y*-axis represents the average absolute value of coefficients across the 200 resampling iterations. Top proteins are labeled with their names and highlighted in color, with the color indicating the organ where the protein is most highly expressed (at least 4 times higher than any other organs).

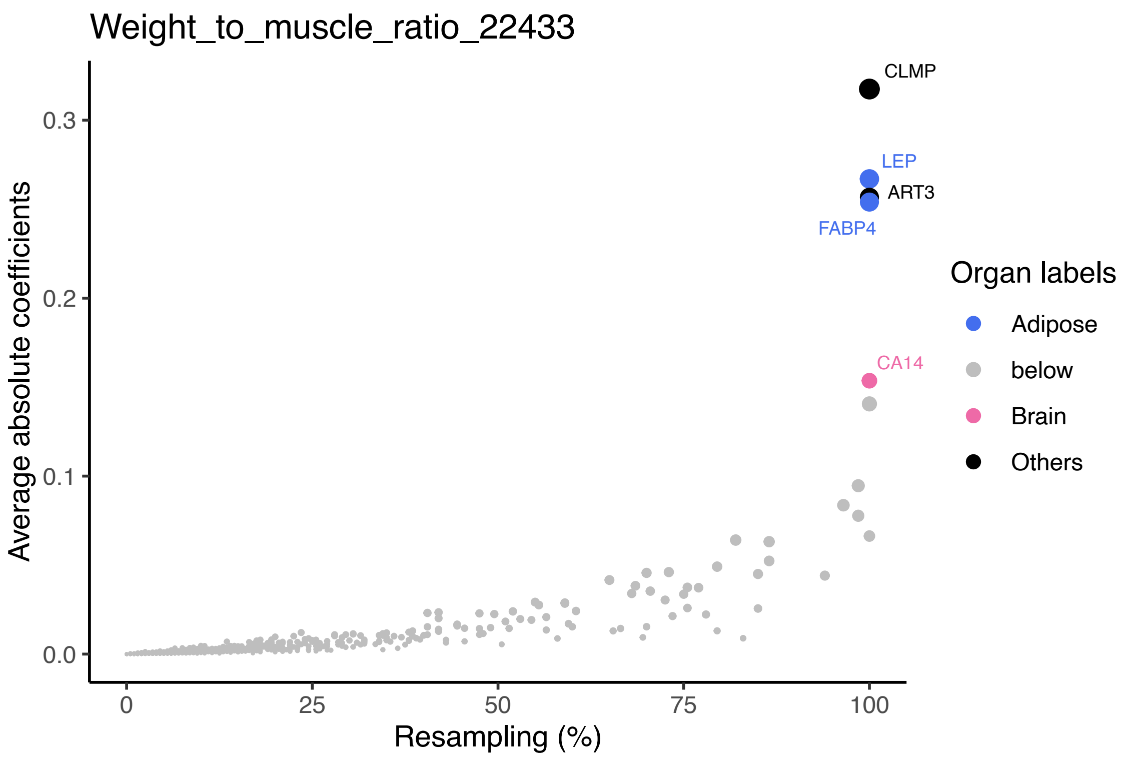


**Fig. S23 Top-ranked plasma protein predictors for abdominal MRI.**

Predictive power of plasma proteins for weight to muscle ratio. The *x*-axis represents the frequency of proteins appearing across 200 resampling iterations, while the *y*-axis represents the average absolute value of coefficients across the 200 resampling iterations. Top proteins are labeled with their names and highlighted in color, with the color indicating the organ where the protein is most highly expressed (at least 4 times higher than any other organs).

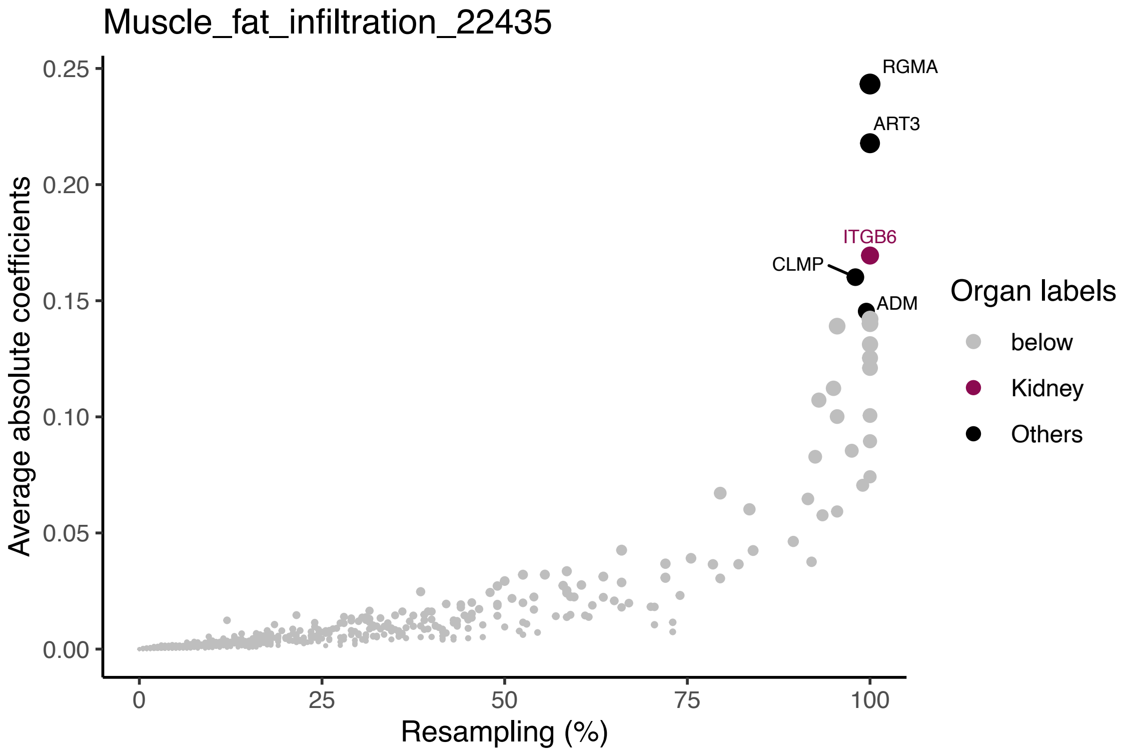


**Fig. S24 Top-ranked plasma protein predictors for abdominal MRI.**

Predictive power of plasma proteins for muscle fat infiltration. The *x*-axis represents the frequency of proteins appearing across 200 resampling iterations, while the *y*-axis represents the average absolute value of coefficients across the 200 resampling iterations. Top proteins are labeled with their names and highlighted in color, with the color indicating the organ where the protein is most highly expressed (at least 4 times higher than any other organs).

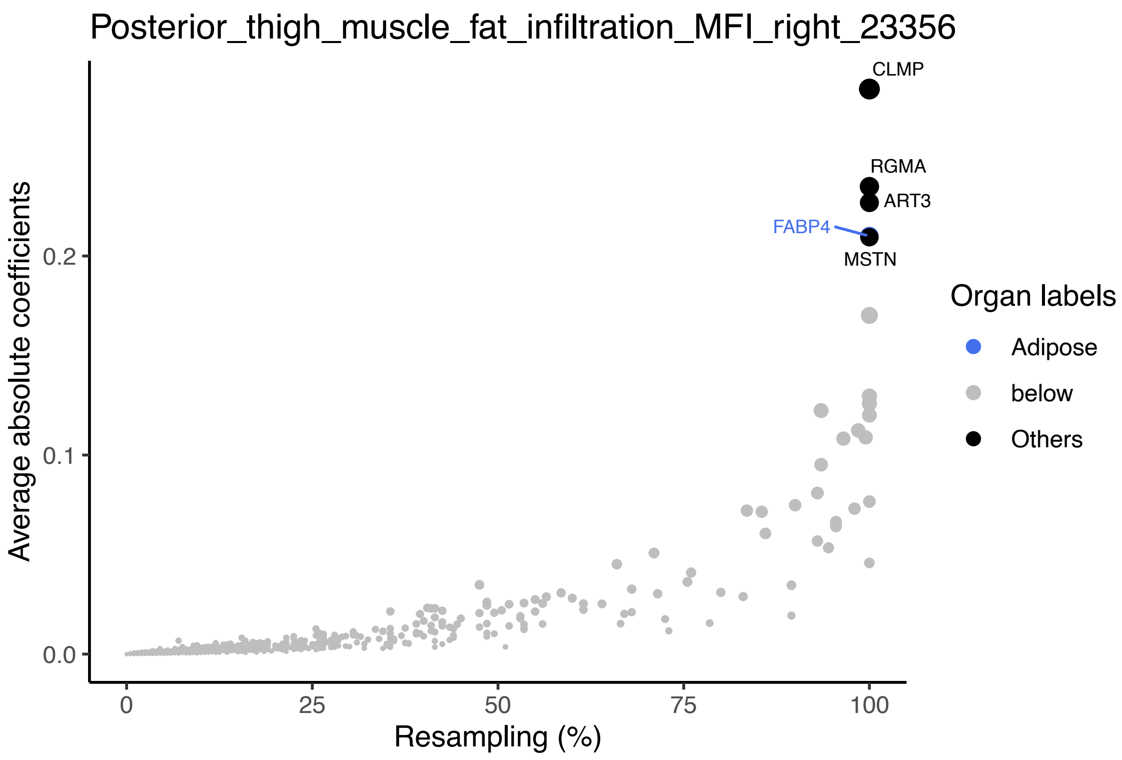


**Fig. S25 Top-ranked plasma protein predictors for abdominal MRI**.

Predictive power of plasma proteins for muscle fat infiltration in the right posterior thigh. The *x*-axis represents the frequency of proteins appearing across 200 resampling iterations, while the *y*-axis represents the average absolute value of coefficients across the 200 resampling iterations. Top proteins are labeled with their names and highlighted in color, with the color indicating the organ where the protein is most highly expressed (at least 4 times higher than any other organs).

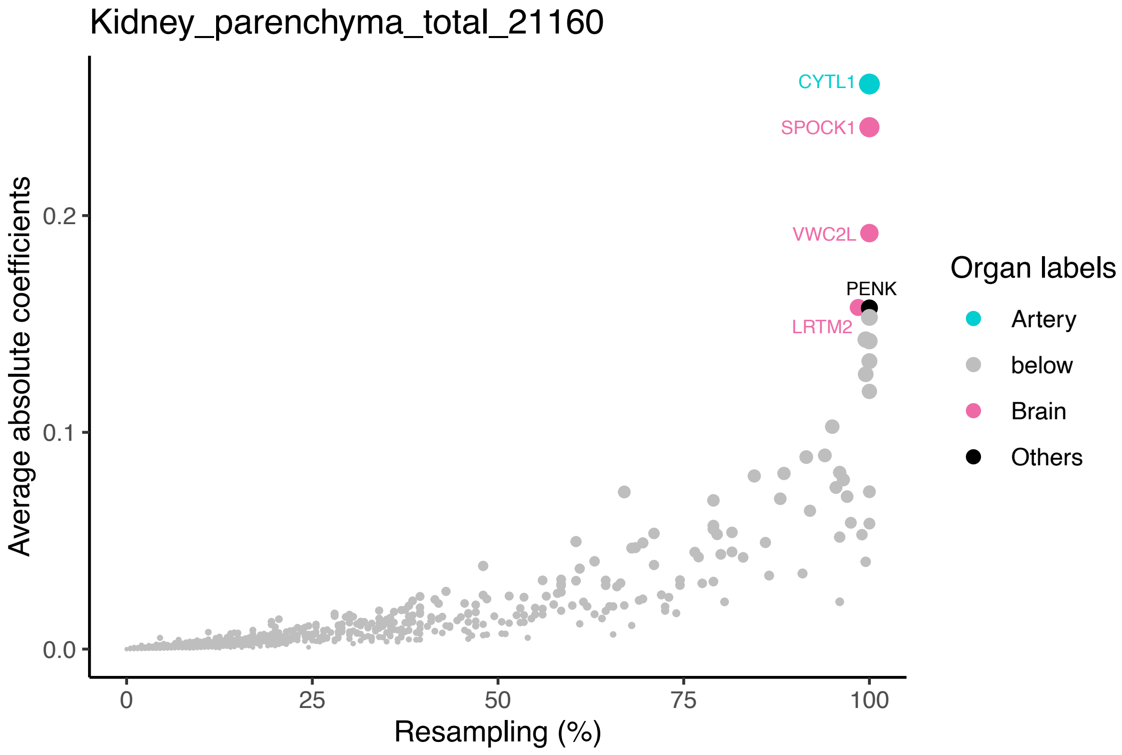


**Fig. S26 Top-ranked plasma protein predictors for abdominal MRI.**

Predictive power of plasma proteins for total parenchymal kidney volume. The *x*-axis represents the frequency of proteins appearing across 200 resampling iterations, while the *y*-axis represents the average absolute value of coefficients across the 200 resampling iterations. Top proteins are labeled with their names and highlighted in color, with the color indicating the organ where the protein is most highly expressed (at least 4 times higher than any other organs).

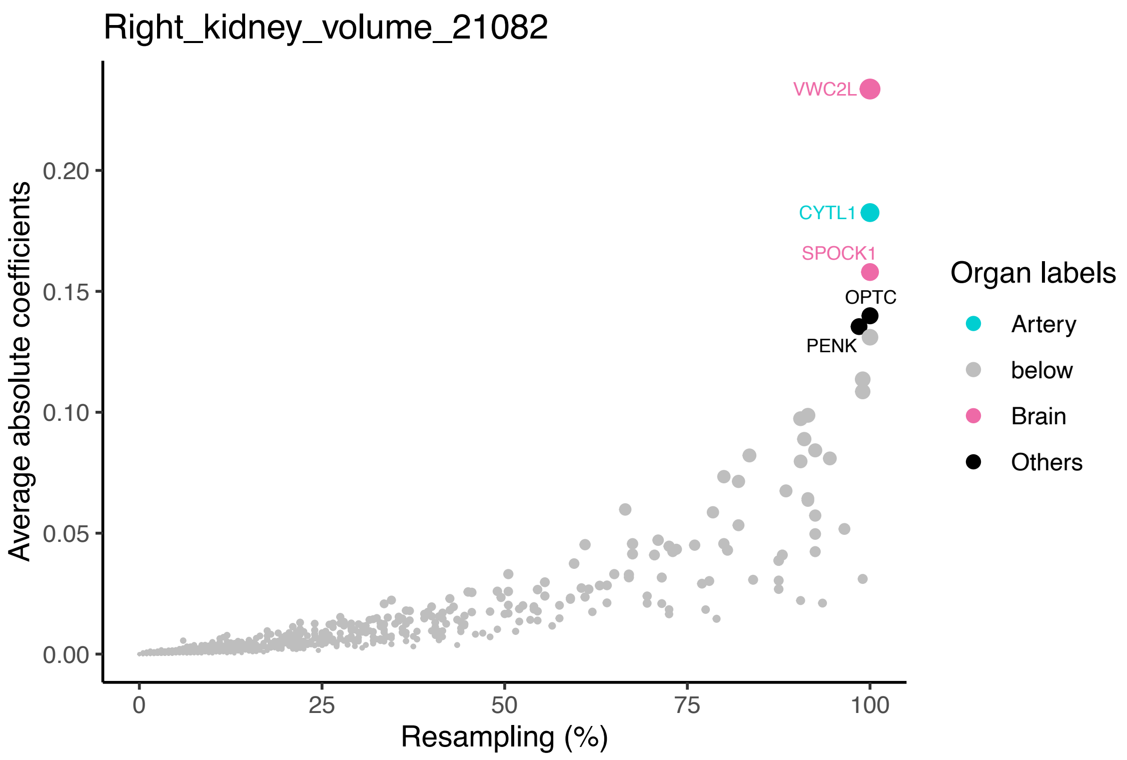


**Fig. S27 Top-ranked plasma protein predictors for abdominal MRI.**

Predictive power of plasma proteins for right kidney volume. The *x*-axis represents the frequency of proteins appearing across 200 resampling iterations, while the *y*-axis represents the average absolute value of coefficients across the 200 resampling iterations. Top proteins are labeled with their names and highlighted in color, with the color indicating the organ where the protein is most highly expressed (at least 4 times higher than any other organs).

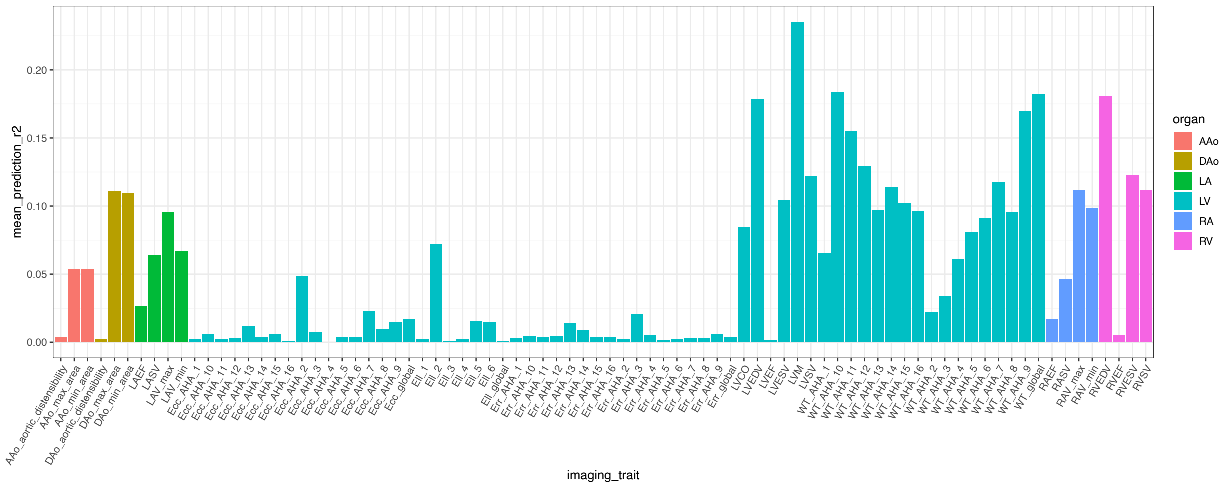


**Fig. S28 Predictive $R^2$ of cardiac and aortic imaging-derived phenotypes (IDPs).**

The mean prediction $R^2$ (*y*-axis) for cardiac and aortic IDPs (*x*-axis) across 200 resampling iterations. Colors represent the categories associated with each cardiac or aortic IDP.

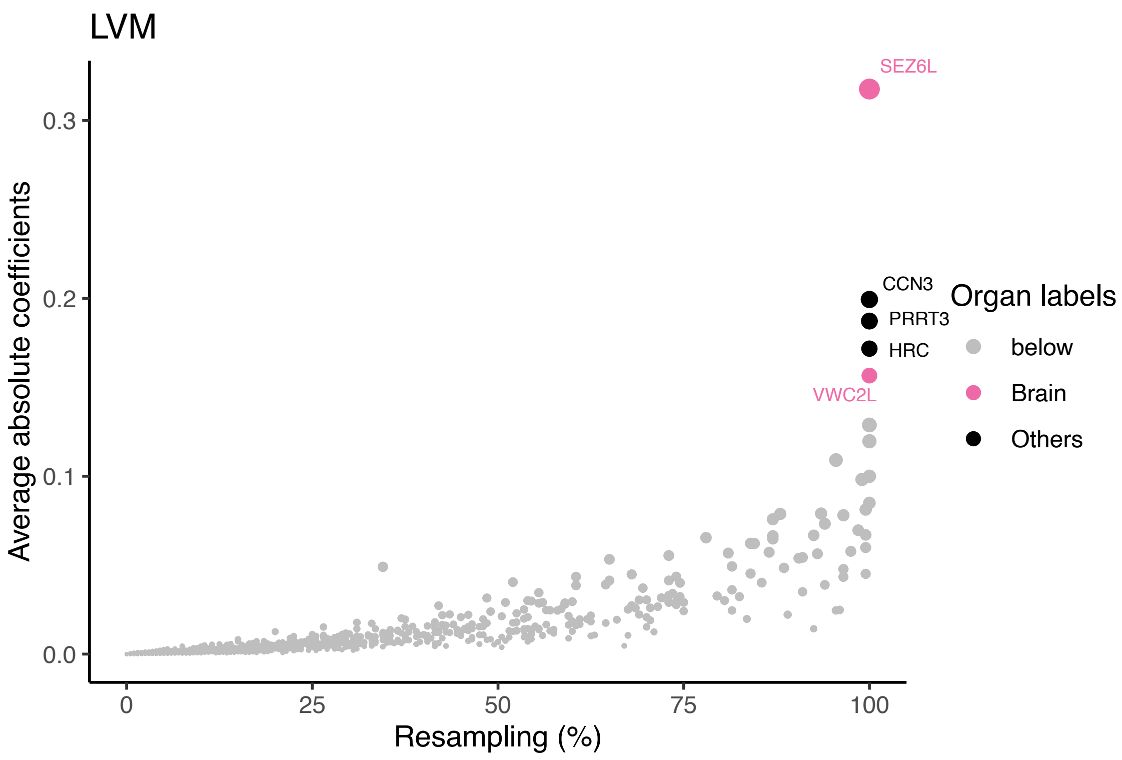


**Fig. S29 Top-ranked plasma protein predictors for cardiac MRI.**

Predictive power of plasma proteins for left ventricular myocardial mass (LVM). The *x*-axis represents the frequency of proteins appearing across 200 resampling iterations, while the *y*-axis represents the average absolute value of coefficients across the 200 resampling iterations. Top proteins are labeled with their names and highlighted in color, with the color indicating the organ where the protein is most highly expressed (at least 4 times higher than any other organs).

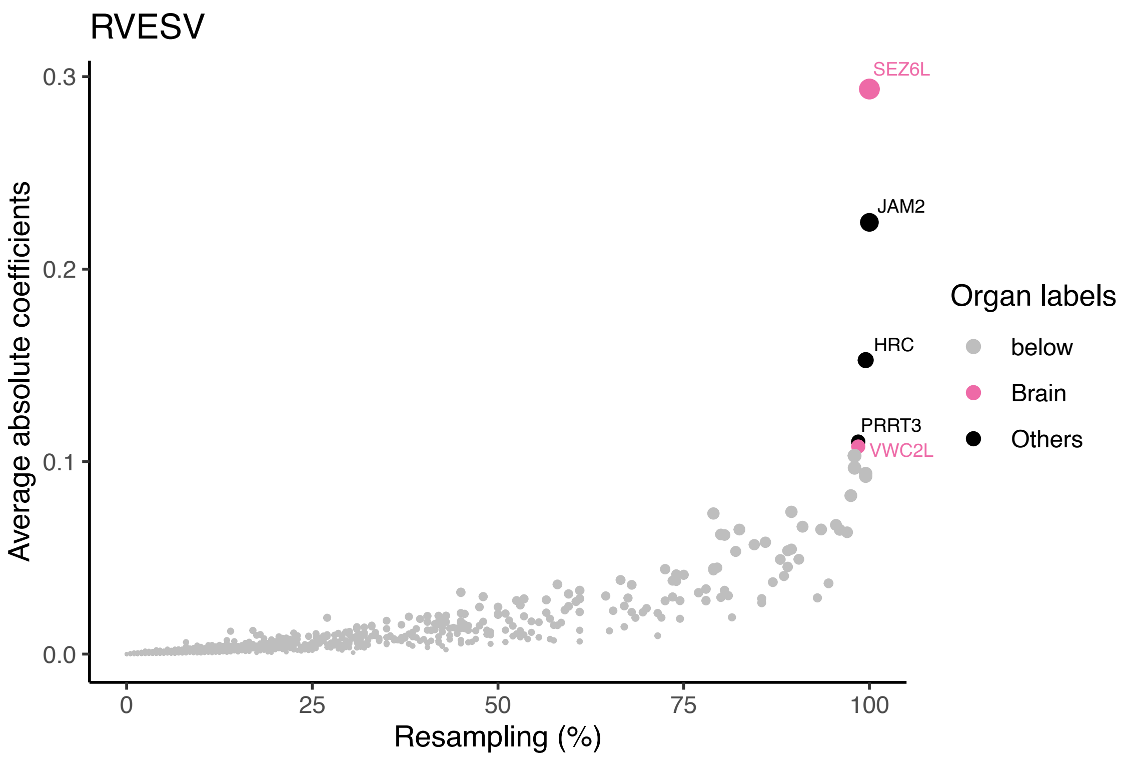


**Fig. S30 Top-ranked plasma protein predictors for cardiac MRI.**

Predictive power of plasma proteins for right ventricular end-systolic volume (RVESV). The *x*-axis represents the frequency of proteins appearing across 200 resampling iterations, while the *y*-axis represents the average absolute value of coefficients across the 200 resampling iterations. Top proteins are labeled with their names and highlighted in color, with the color indicating the organ where the protein is most highly expressed (at least 4 times higher than any other organs).

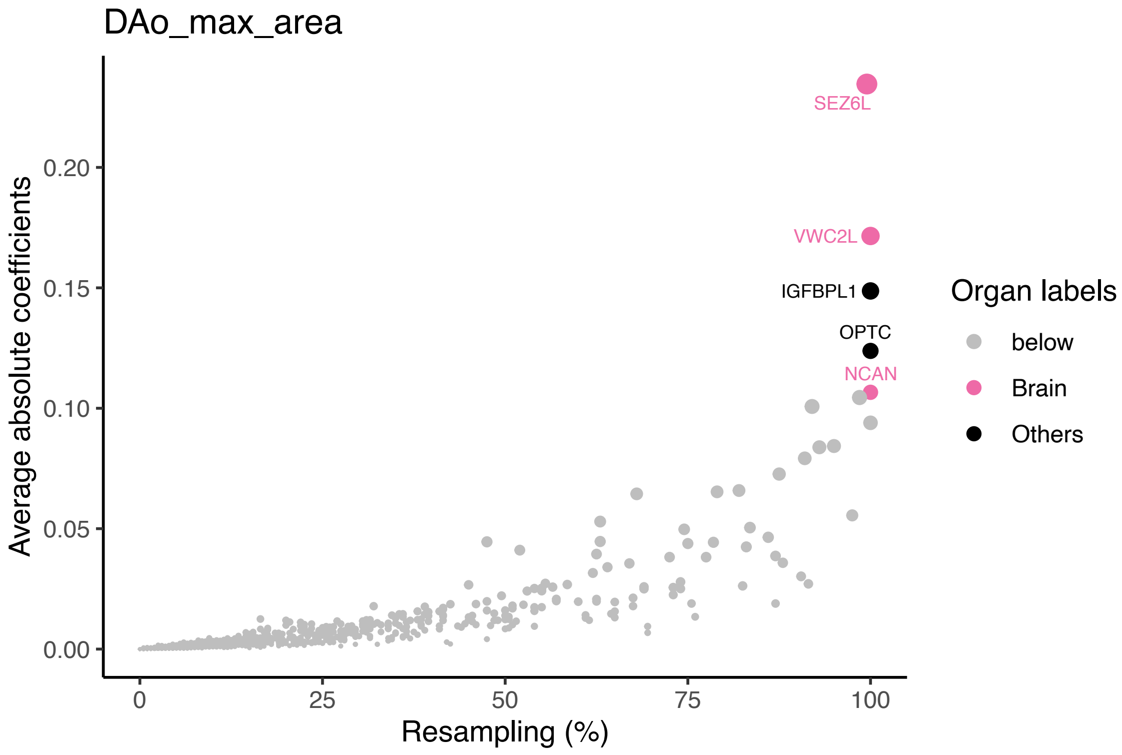


**Fig. S31 Top-ranked plasma protein predictors for aortic MRI.**

Predictive power of plasma proteins for descending aorta maximum area (Dao max area). The *x*-axis represents the frequency of proteins appearing across 200 resampling iterations, while the *y*-axis represents the average absolute value of coefficients across the 200 resampling iterations. Top proteins are labeled with their names and highlighted in color, with the color indicating the organ where the protein is most highly expressed (at least 4 times higher than any other organs).

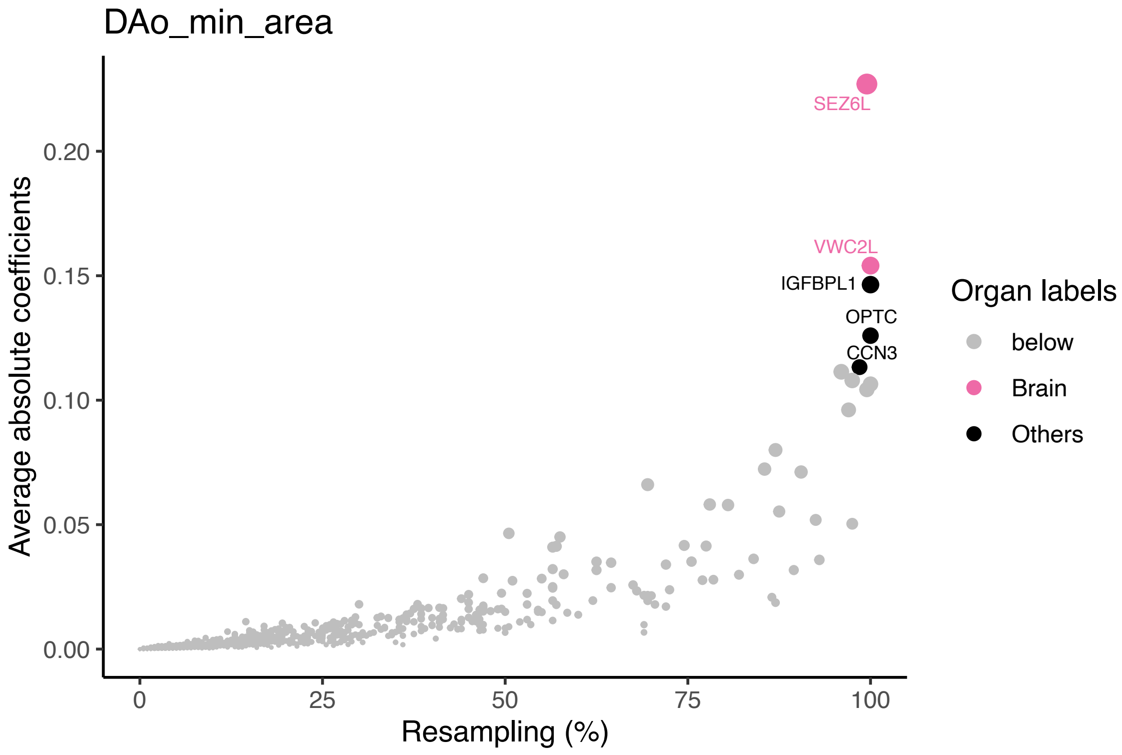


**Fig. S32 Top-ranked plasma protein predictors for aortic MRI.**

Predictive power of plasma proteins for descending aorta minimum area (Dao min area). The *x*-axis represents the frequency of proteins appearing across 200 resampling iterations, while the *y*-axis represents the average absolute value of coefficients across the 200 resampling iterations. Top proteins are labeled with their names and highlighted in color, with the color indicating the organ where the protein is most highly expressed (at least 4 times higher than any other organs).

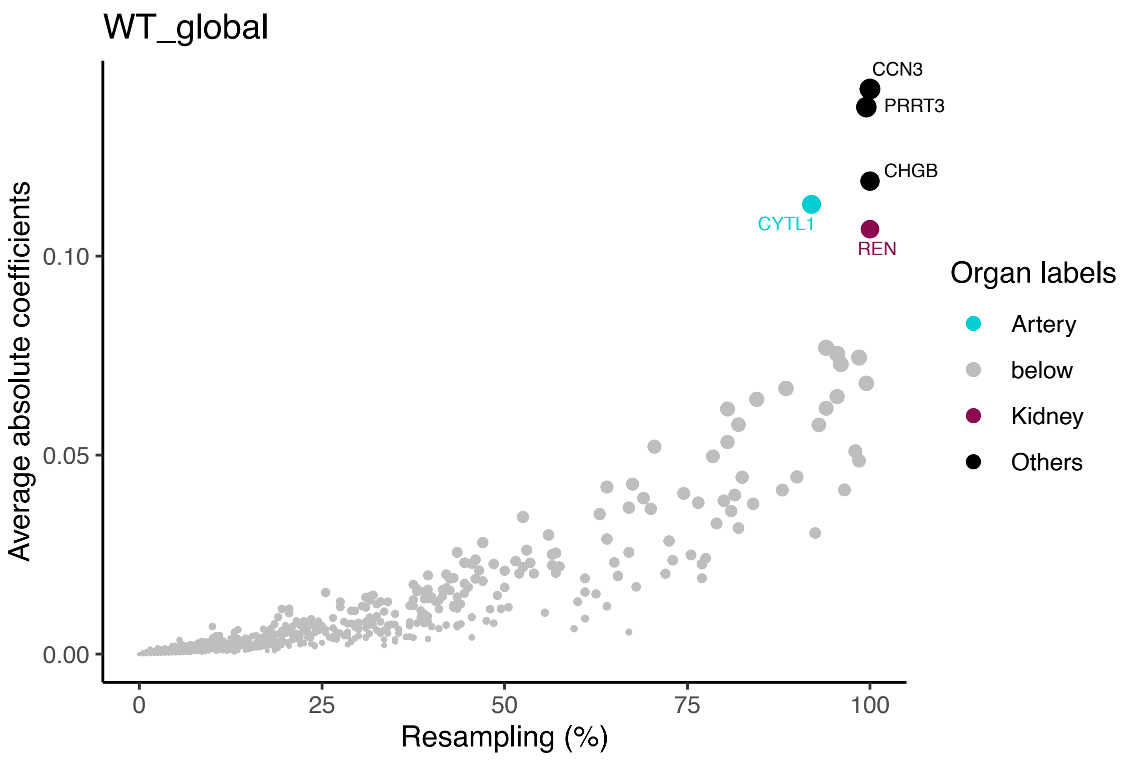


**Fig. S33 Top-ranked plasma protein predictors for cardiac MRI.**

Predictive power of plasma proteins for global myocardial-wall thickness at end-diastole (WT global). The *x*-axis represents the frequency of proteins appearing across 200 resampling iterations, while the *y*-axis represents the average absolute value of coefficients across the 200 resampling iterations. Top proteins are labeled with their names and highlighted in color, with the color indicating the organ where the protein is most highly expressed (at least 4 times higher than any other organs).

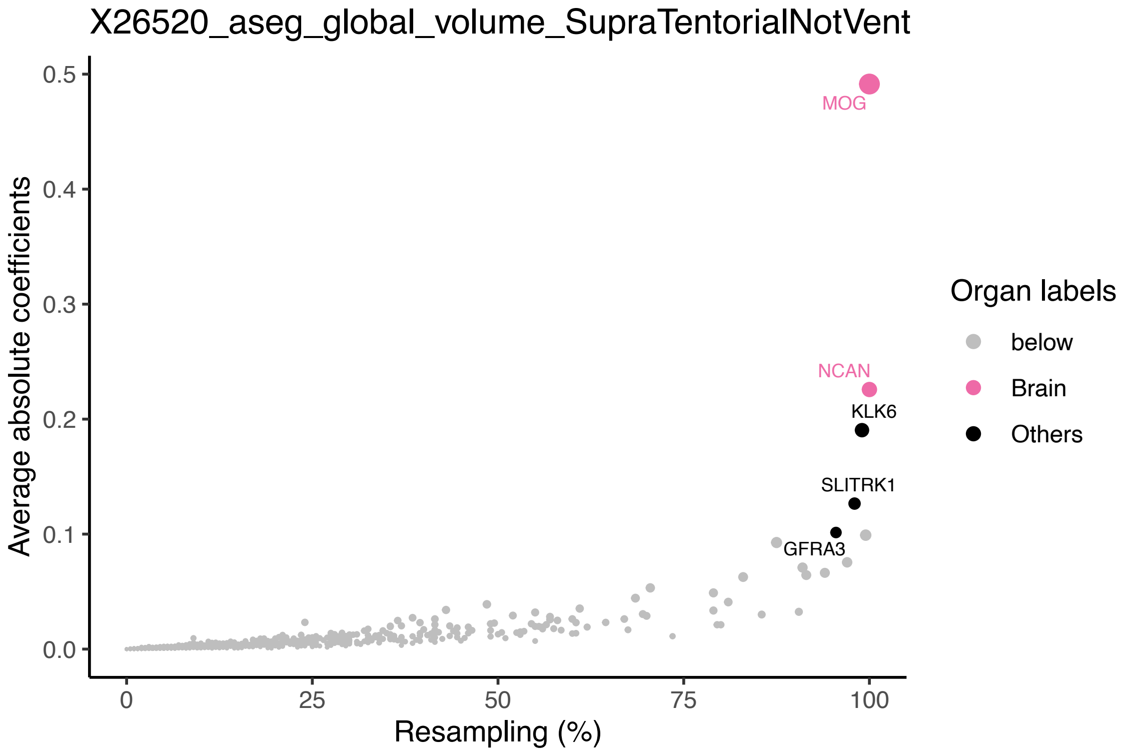


**Fig. S34 Top-ranked plasma protein predictors for brain structural MRI.**

Predictive power of plasma proteins for supratentorial brain volume, excluding ventricles. The *x*-axis represents the frequency of proteins appearing across 200 resampling iterations, while the *y*-axis represents the average absolute value of coefficients across the 200 resampling iterations. Top proteins are labeled with their names and highlighted in color, with the color indicating the organ where the protein is most highly expressed (at least 4 times higher than any other organs).

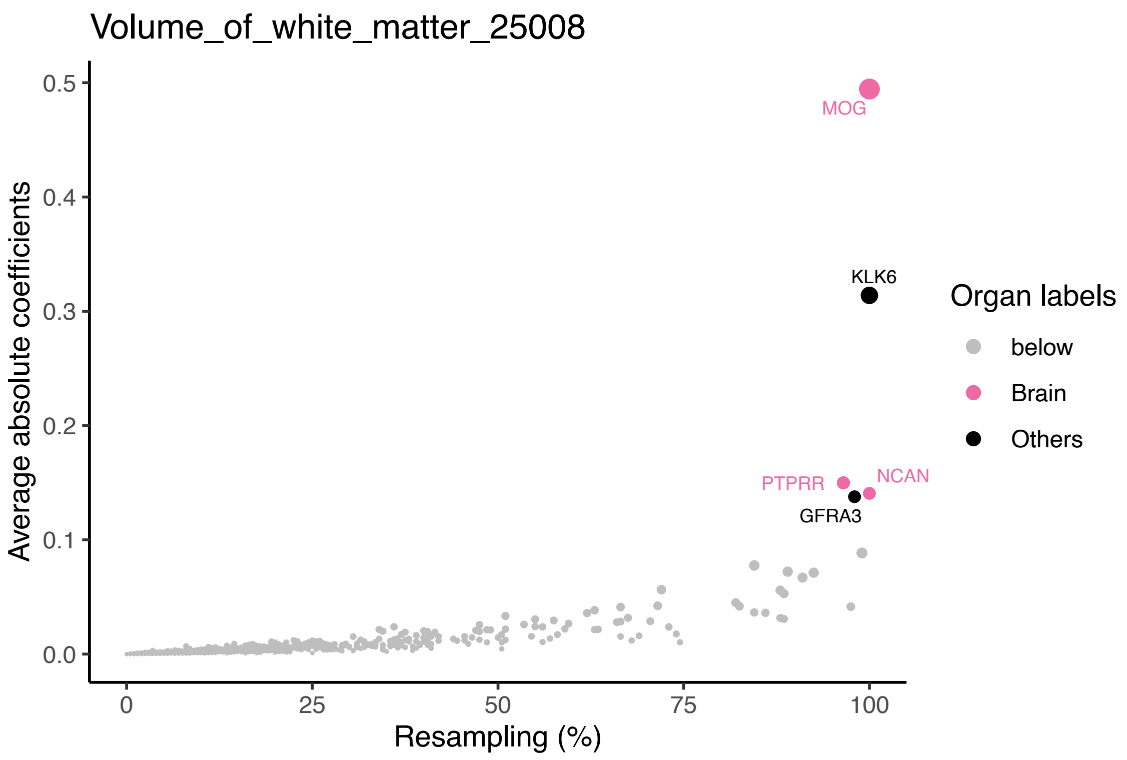


**Fig. S35 Top-ranked plasma protein predictors for brain structural MRI.**

Predictive power of plasma proteins for brain white matter volume. The *x*-axis represents the frequency of proteins appearing across 200 resampling iterations, while the *y*-axis represents the average absolute value of coefficients across the 200 resampling iterations. Top proteins are labeled with their names and highlighted in color, with the color indicating the organ where the protein is most highly expressed (at least 4 times higher than any other organs).

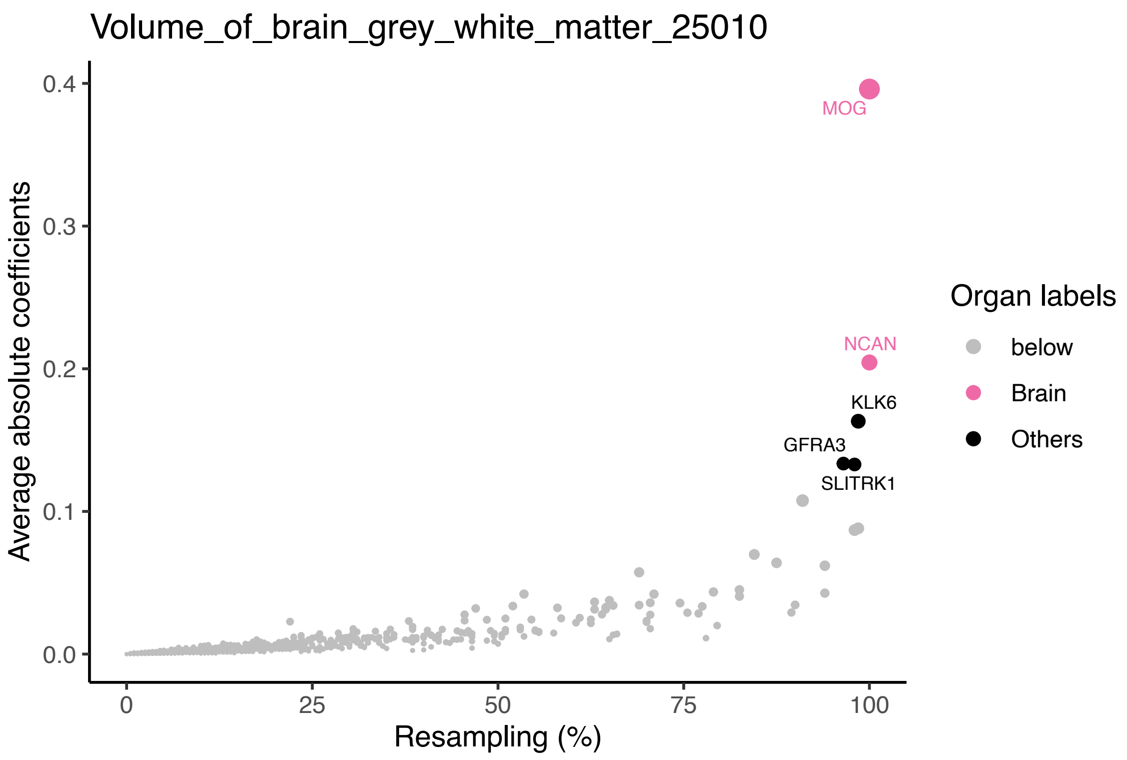


**Fig. S36 Top-ranked plasma protein predictors for brain structural MRI.**

Predictive power of plasma proteins for combined white and grey matter volume of the brain. The *x*-axis represents the frequency of proteins appearing across 200 resampling iterations, while the *y*-axis represents the average absolute value of coefficients across the 200 resampling iterations. Top proteins are labeled with their names and highlighted in color, with the color indicating the organ where the protein is most highly expressed (at least 4 times higher than any other organs).

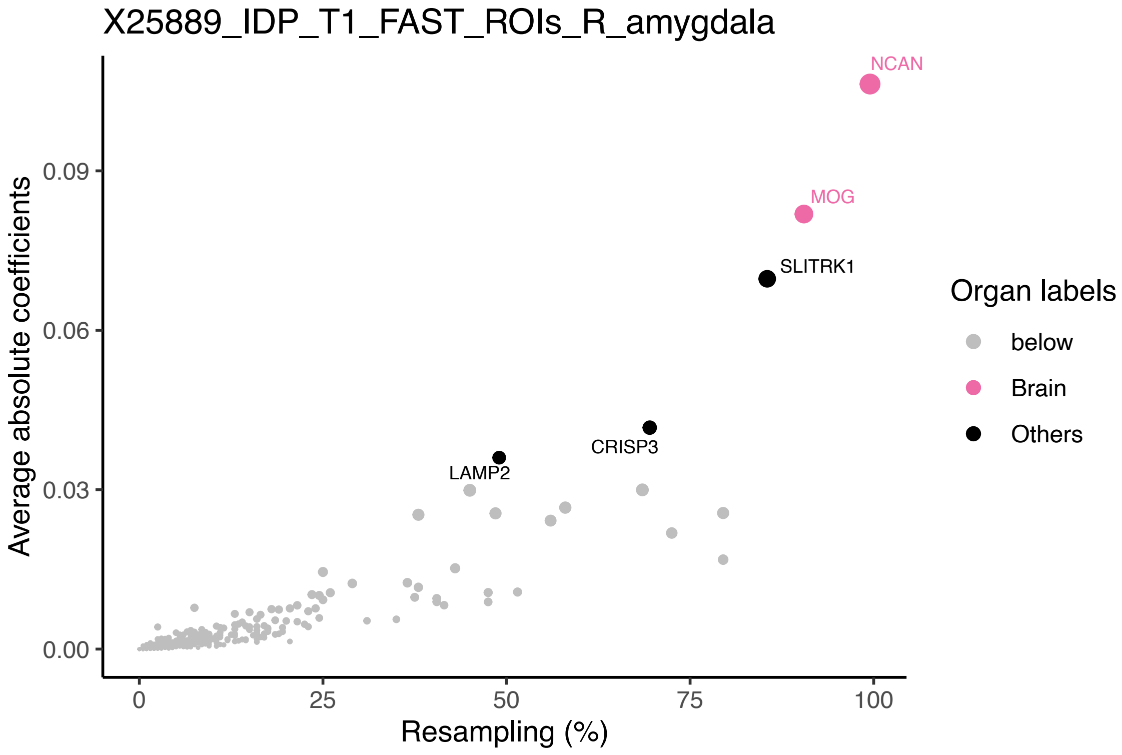


**Fig. S37 Top-ranked plasma protein predictors for brain structural MRI.**

Predictive power of plasma proteins for volume of grey matter in right amygdala. The *x*-axis represents the frequency of proteins appearing across 200 resampling iterations, while the *y*-axis represents the average absolute value of coefficients across the 200 resampling iterations. Top proteins are labeled with their names and highlighted in color, with the color indicating the organ where the protein is most highly expressed (at least 4 times higher than any other organs).

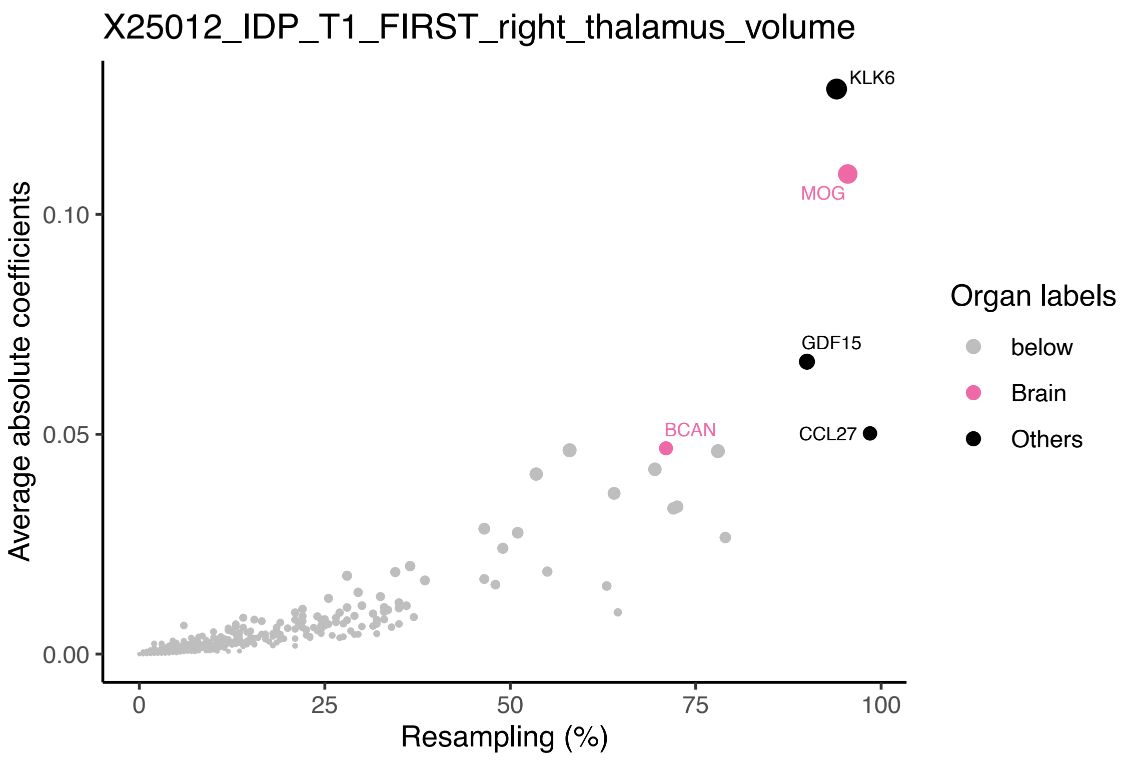


**Fig. S38 Top-ranked plasma protein predictors for brain structural MRI.**

Predictive power of plasma proteins for the volume of right thalamus. The *x*-axis represents the frequency of proteins appearing across 200 resampling iterations, while the *y*-axis represents the average absolute value of coefficients across the 200 resampling iterations. Top proteins are labeled with their names and highlighted in color, with the color indicating the organ where the protein is most highly expressed (at least 4 times higher than any other organs).

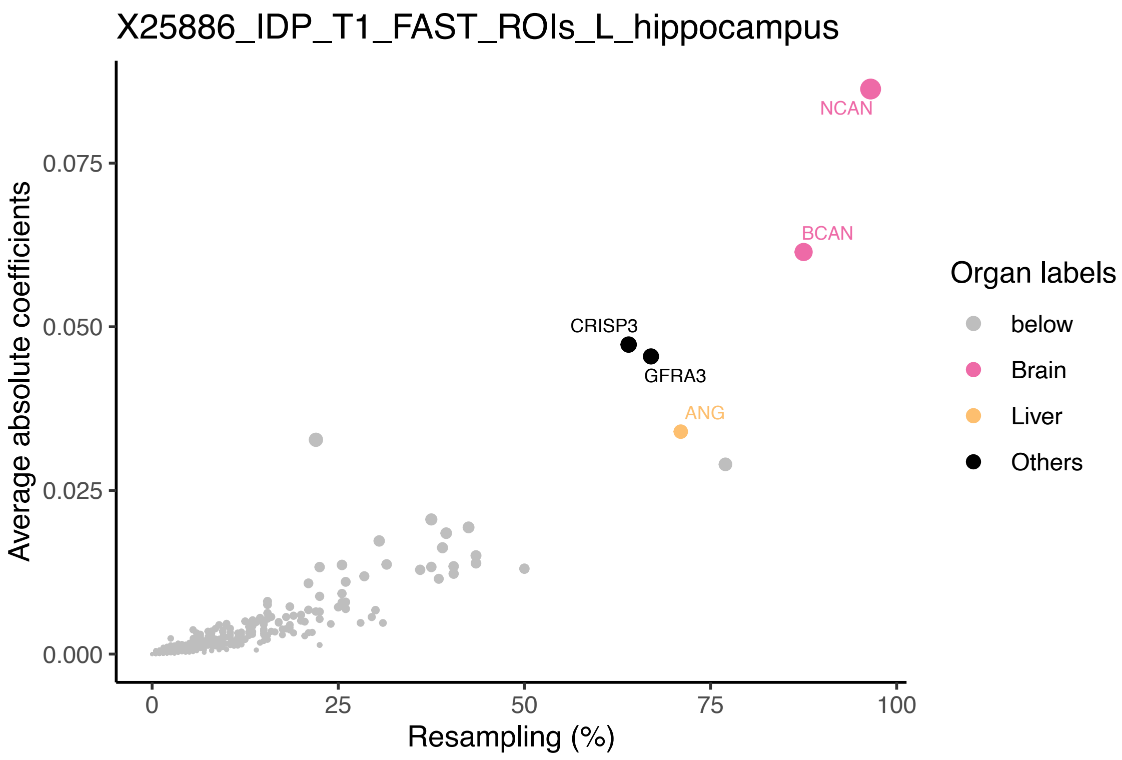


**Fig. S39 Top-ranked plasma protein predictors for brain structural MRI.**

Predictive power of plasma proteins for the volume of grey matter in left hippocampus. The *x*-axis represents the frequency of proteins appearing across 200 resampling iterations, while the *y*-axis represents the average absolute value of coefficients across the 200 resampling iterations. Top proteins are labeled with their names and highlighted in color, with the color indicating the organ where the protein is most highly expressed (at least 4 times higher than any other organs).

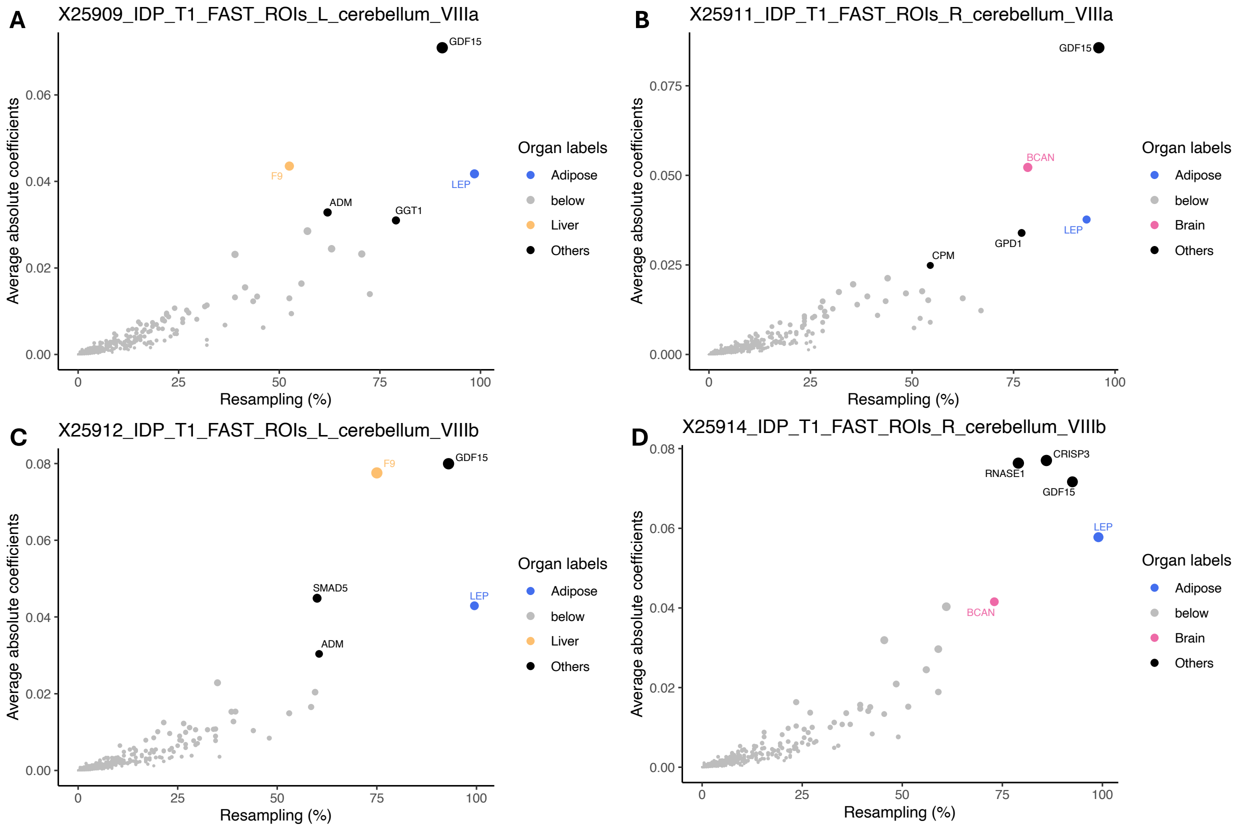


**Fig. S40 Top-ranked plasma protein predictors for brain structural MRI.**

Predictive power of plasma proteins for volumes of grey matter in **A** left VIIIa cerebellum, **B** right VIIIa cerebellum, **C** left VIIIb cerebellum, **D** right VIIIb cerebellum. The *x*-axis represents the frequency of proteins appearing across 200 resampling iterations, while the *y*-axis represents the average absolute value of coefficients across the 200 resampling iterations. Top proteins are labeled with their names and highlighted in color, with the color indicating the organ where the protein is most highly expressed (at least 4 times higher than any other organs).

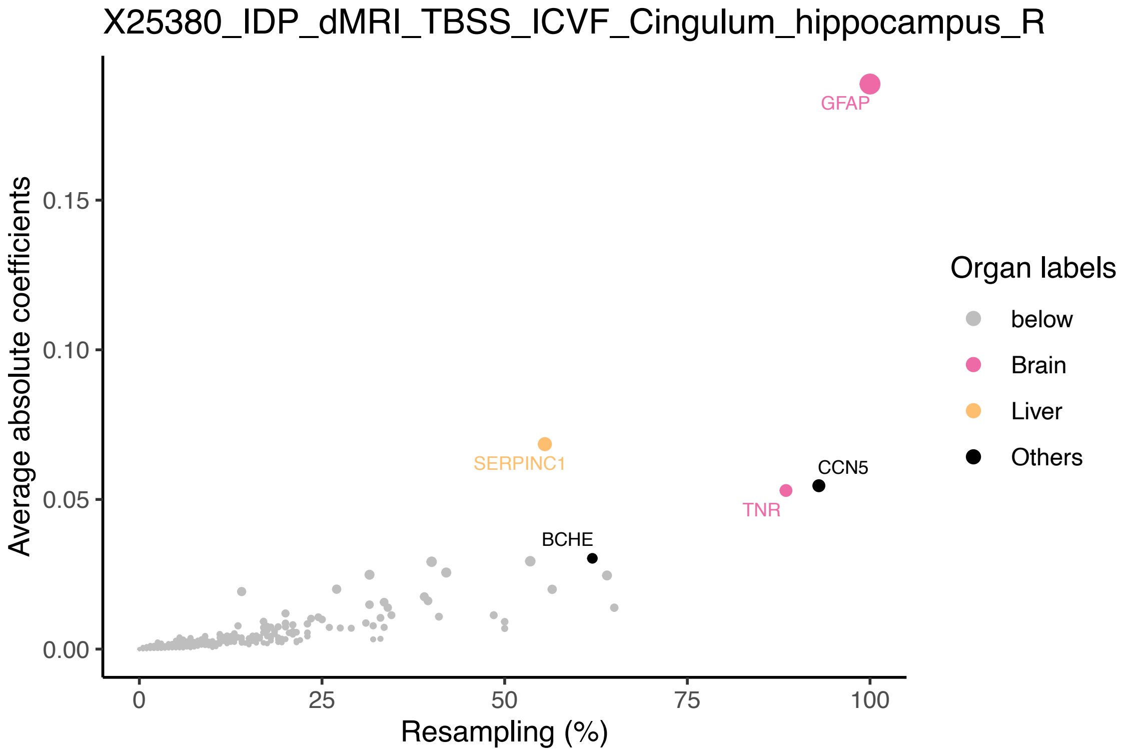


**Fig. S41 Top-ranked plasma protein predictors for brain diffusion MRI.**

Predictive power of plasma proteins for mean intracellular volume fraction (ICVF) in the right cingulum hippocampus. The *x*-axis represents the frequency of proteins appearing across 200 resampling iterations, while the *y*-axis represents the average absolute value of coefficients across the 200 resampling iterations. Top proteins are labeled with their names and highlighted in color, with the color indicating the organ where the protein is most highly expressed (at least 4 times higher than any other organs).

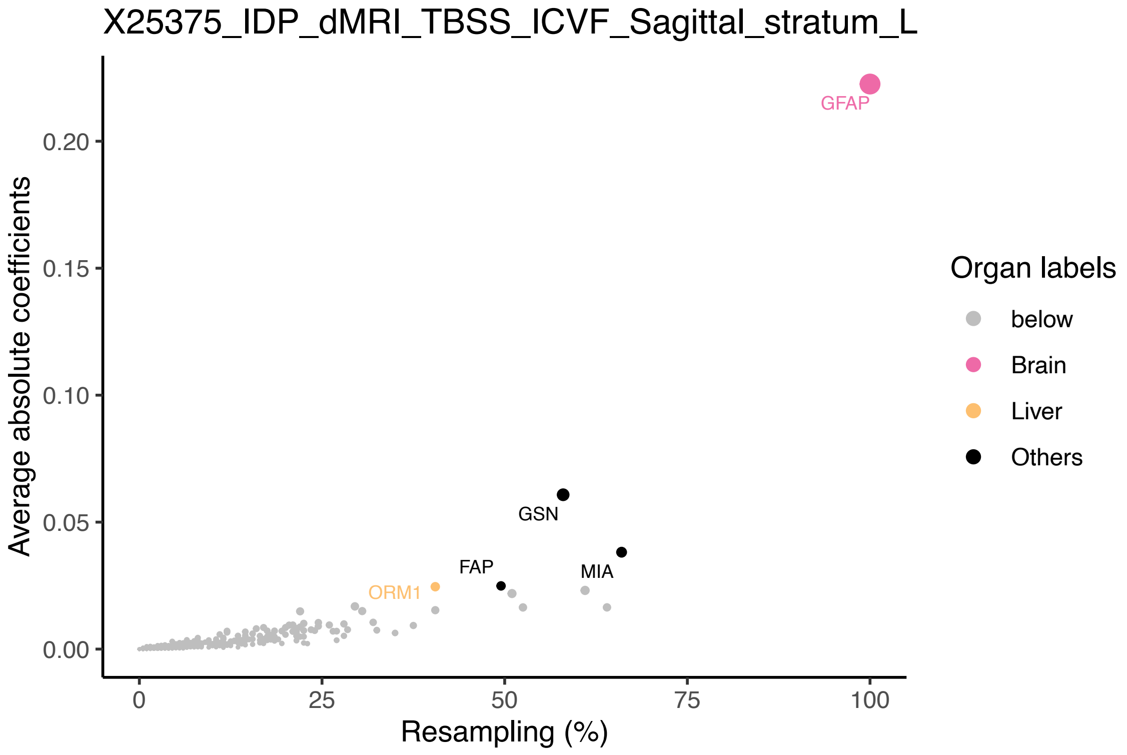


**Fig. S42 Top-ranked plasma protein predictors for brain diffusion MRI.**

Predictive power of plasma proteins for mean intracellular volume fraction (ICVF) in the left sagittal stratum. The *x*-axis represents the frequency of proteins appearing across 200 resampling iterations, while the *y*-axis represents the average absolute value of coefficients across the 200 resampling iterations. Top proteins are labeled with their names and highlighted in color, with the color indicating the organ where the protein is most highly expressed (at least 4 times higher than any other organs).

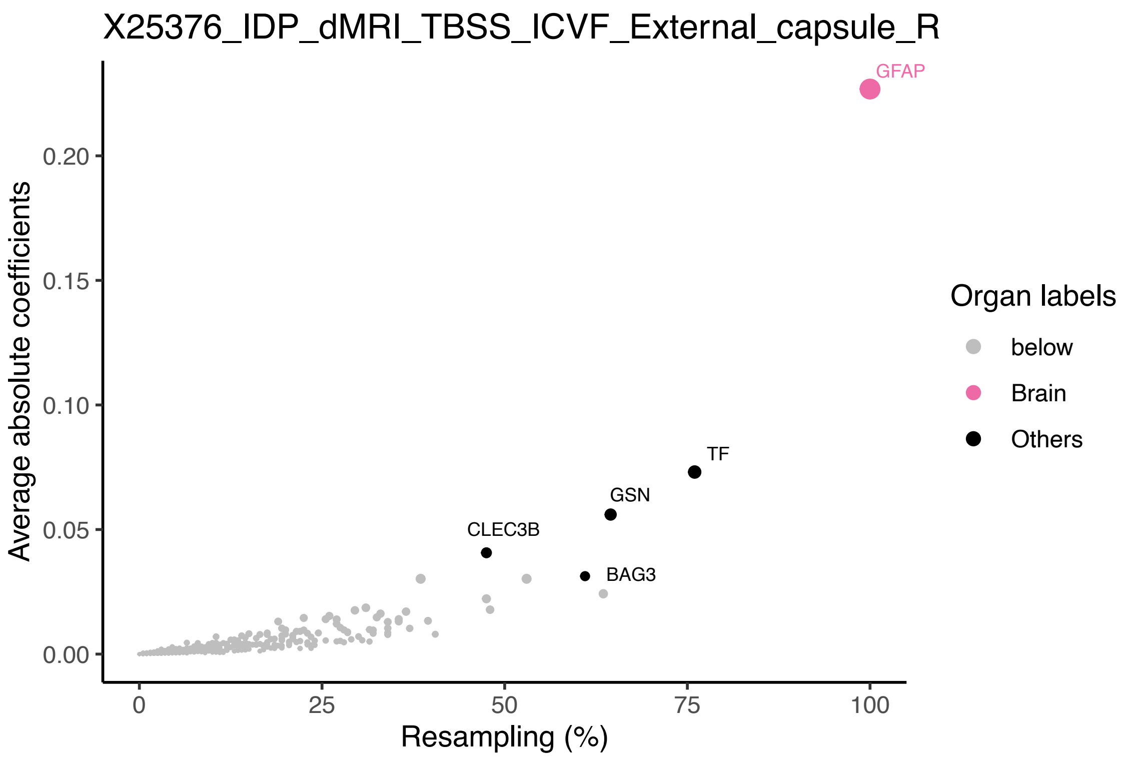


**Fig. S43 Top-ranked plasma protein predictors for brain diffusion MRI.**

Predictive power of plasma proteins for mean intracellular volume fraction (ICVF) in the right external capsule. The *x*-axis represents the frequency of proteins appearing across 200 resampling iterations, while the *y*-axis represents the average absolute value of coefficients across the 200 resampling iterations. Top proteins are labeled with their names and highlighted in color, with the color indicating the organ where the protein is most highly expressed (at least 4 times higher than any other organs).

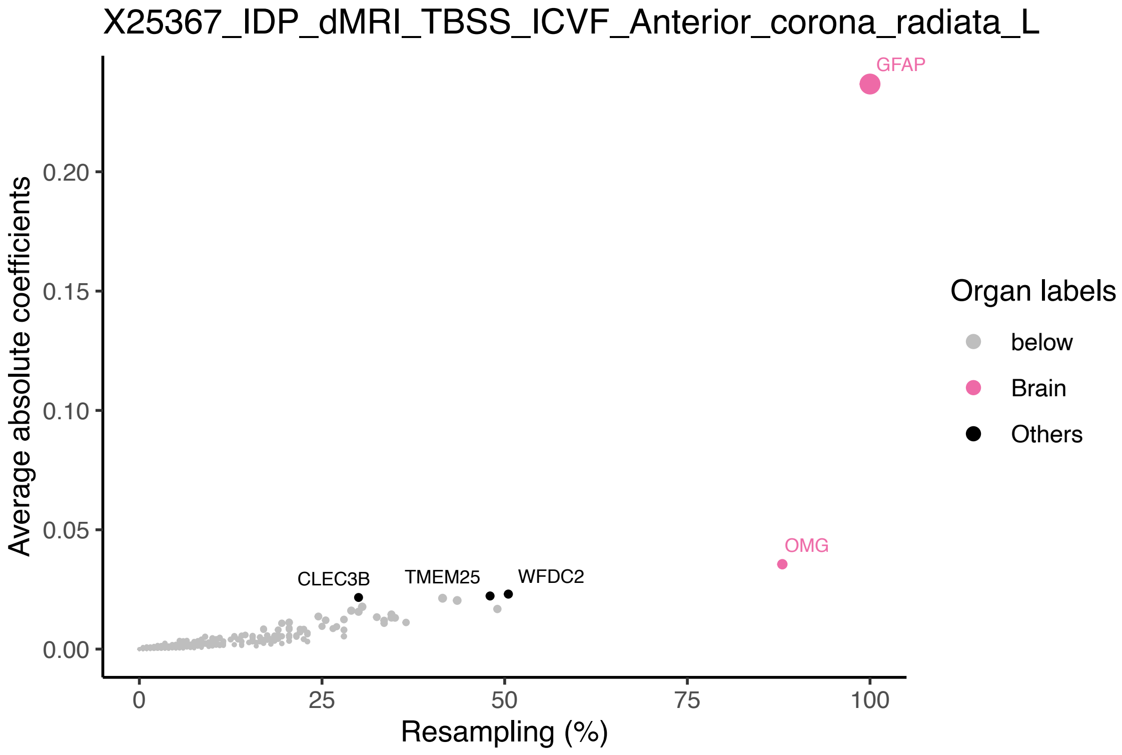


**Fig. S44 Top-ranked plasma protein predictors for brain diffusion MRI.**

Predictive power of plasma proteins for mean intracellular volume fraction (ICVF) in the left anterior corona radiata. The *x*-axis represents the frequency of proteins appearing across 200 resampling iterations, while the *y*-axis represents the average absolute value of coefficients across the 200 resampling iterations. Top proteins are labeled with their names and highlighted in color, with the color indicating the organ where the protein is most highly expressed (at least 4 times higher than any other organs).

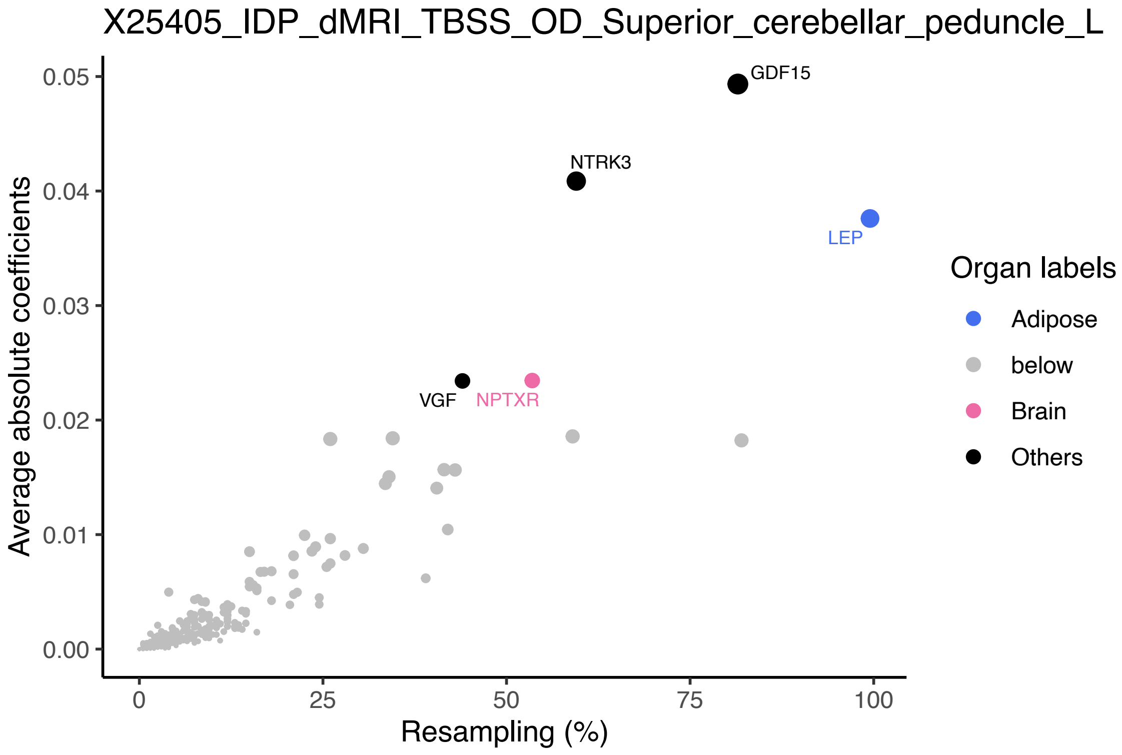


**Fig. S45 Top-ranked plasma protein predictors for brain diffusion MRI.**

Predictive power of plasma proteins for mean orientation dispersion index (OD) in the left superior cerebellar peduncle. The *x*-axis represents the frequency of proteins appearing across 200 resampling iterations, while the *y*-axis represents the average absolute value of coefficients across the 200 resampling iterations. Top proteins are labeled with their names and highlighted in color, with the color indicating the organ where the protein is most highly expressed (at least 4 times higher than any other organs).

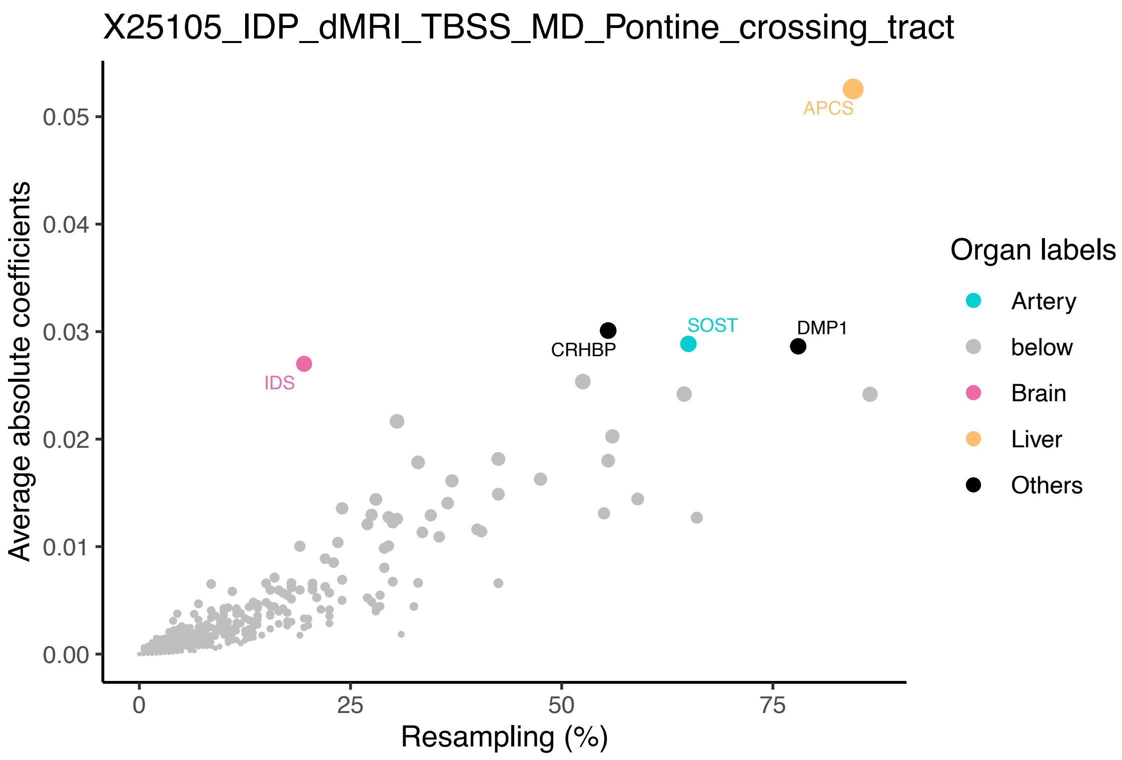


**Fig. S46 Top-ranked plasma protein predictors for brain diffusion MRI.**

Predictive power of plasma proteins for mean diffusivity (MD) in the pontine crossing tract. The *x*-axis represents the frequency of proteins appearing across 200 resampling iterations, while the *y*-axis represents the average absolute value of coefficients across the 200 resampling iterations. Top proteins are labeled with their names and highlighted in color, with the color indicating the organ where the protein is most highly expressed (at least 4 times higher than any other organs).

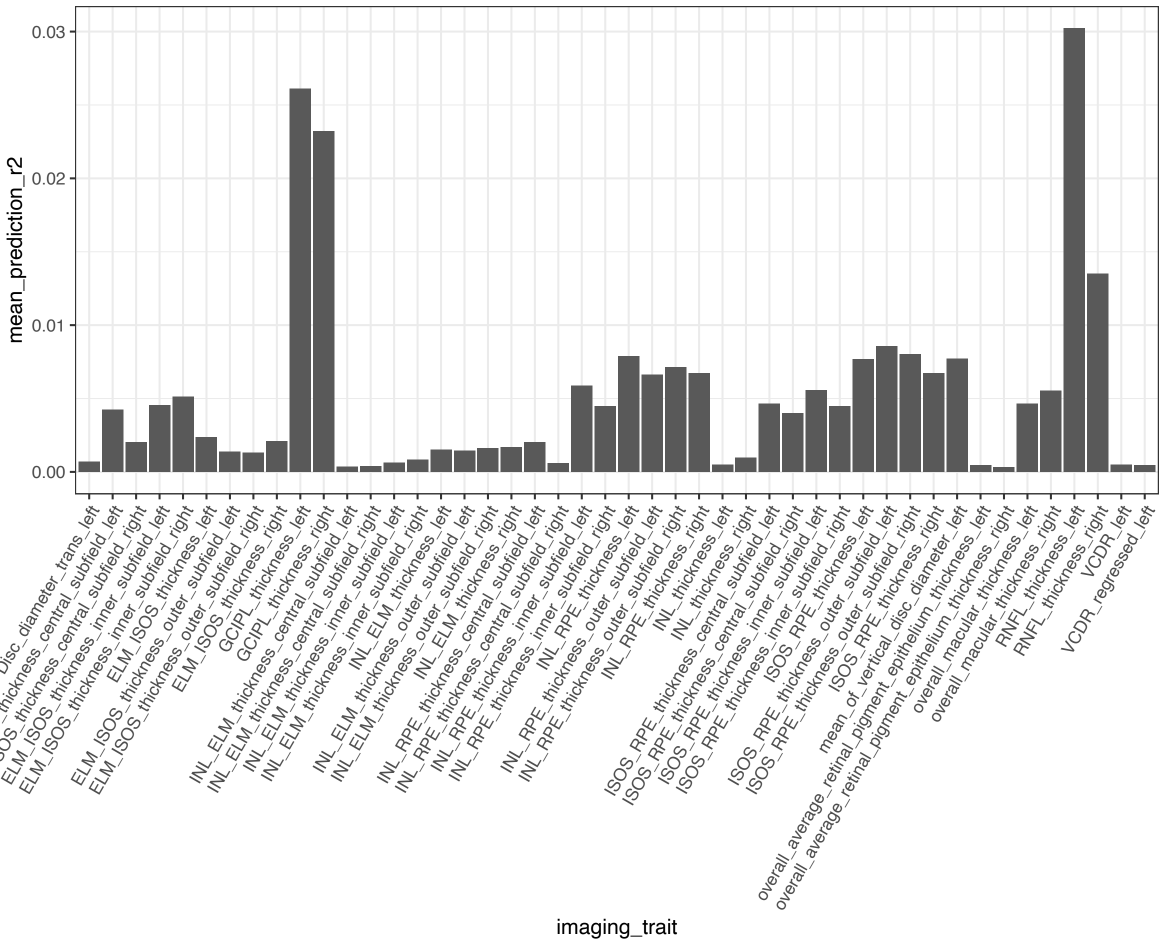


**Fig. S47 Predictive $R^2$ of eye imaging-derived phenotypes.**

The mean prediction $R^2$ (*y*-axis) for eye optical coherence tomography measures (*x*-axis) across 200 resampling iterations.

5

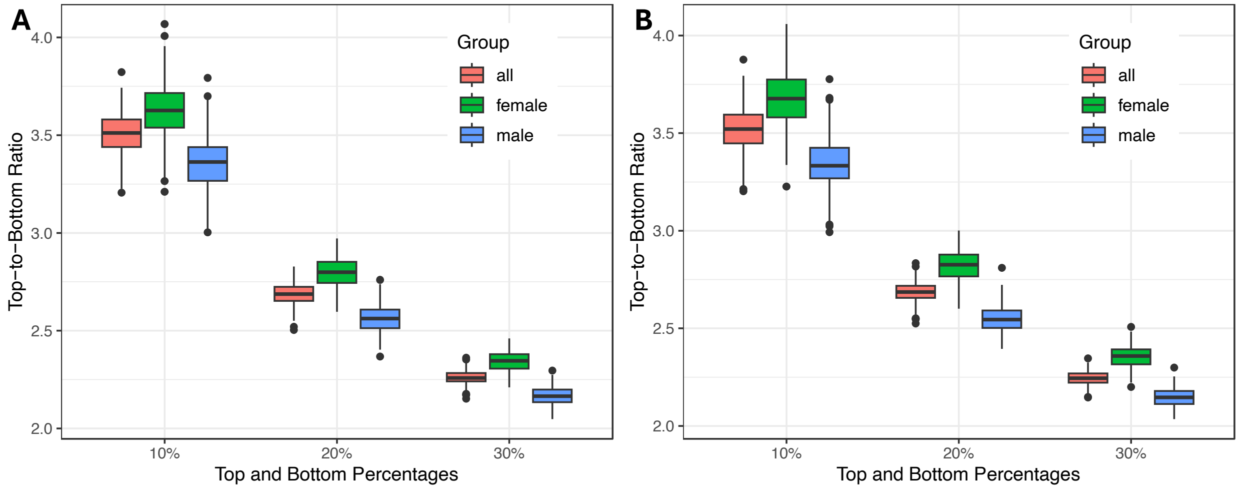


**Fig. S48 Stratification capability of plasma protein prediction models for body fat traits.**

The top-to-bottom ratios of **A** total trunk fat volume and **B** total abdominal adipose tissue index across percentile groups representing the top and bottom 10%, 20%, and 30% of predicted imaging traits based on plasma proteins. The *x*-axis represents the percentile groups, and the *y*-axis represents the top-to-bottom ratios. Sex groups, including all individuals, females, and males, are distinguished by color.

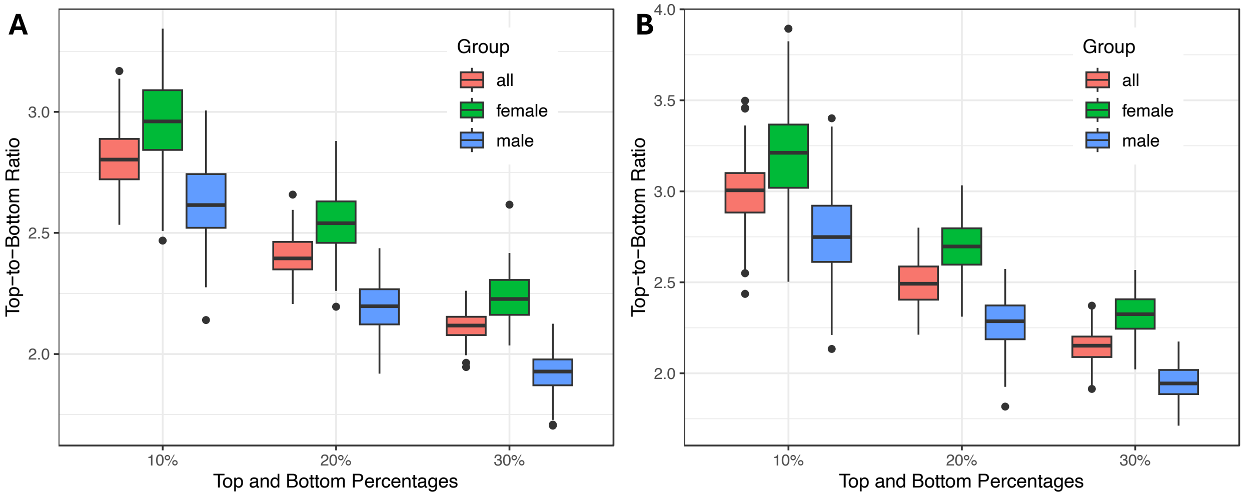


**Fig. S49 Stratification capability of plasma protein prediction models for organ-specific fat traits.**

The top-to-bottom ratios of **A** liver fat fraction and **B** pancreas fat fraction across percentile groups representing the top and bottom 10%, 20%, and 30% of predicted imaging traits based on plasma proteins. The x-axis represents the percentile groups, and the y-axis represents the top-to-bottom ratios. Sex groups, including all individuals, females, and males, are distinguished by color.

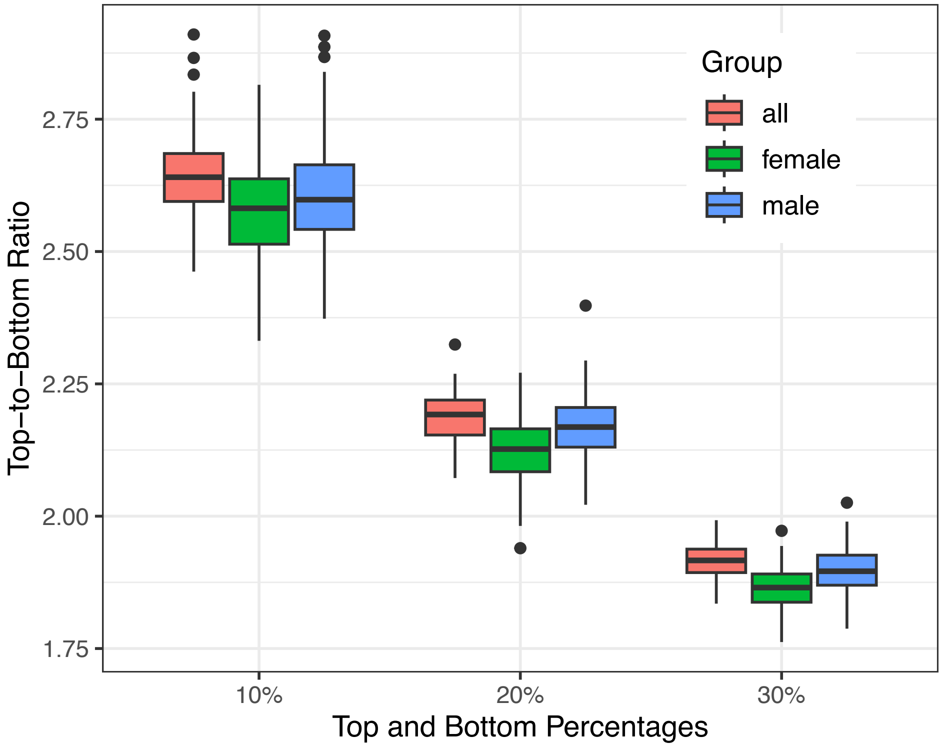


**Fig. S50 Stratification capability of plasma protein prediction models for organ-specific volume traits.**

The top-to-bottom ratios of spleen volume across percentile groups representing the top and bottom 10%, 20%, and 30% of predicted imaging traits based on plasma proteins. The *x*-axis represents the percentile groups, and the *y*-axis represents the top-to-bottom ratios. Sex groups, including all individuals, females, and males, are distinguished by color.

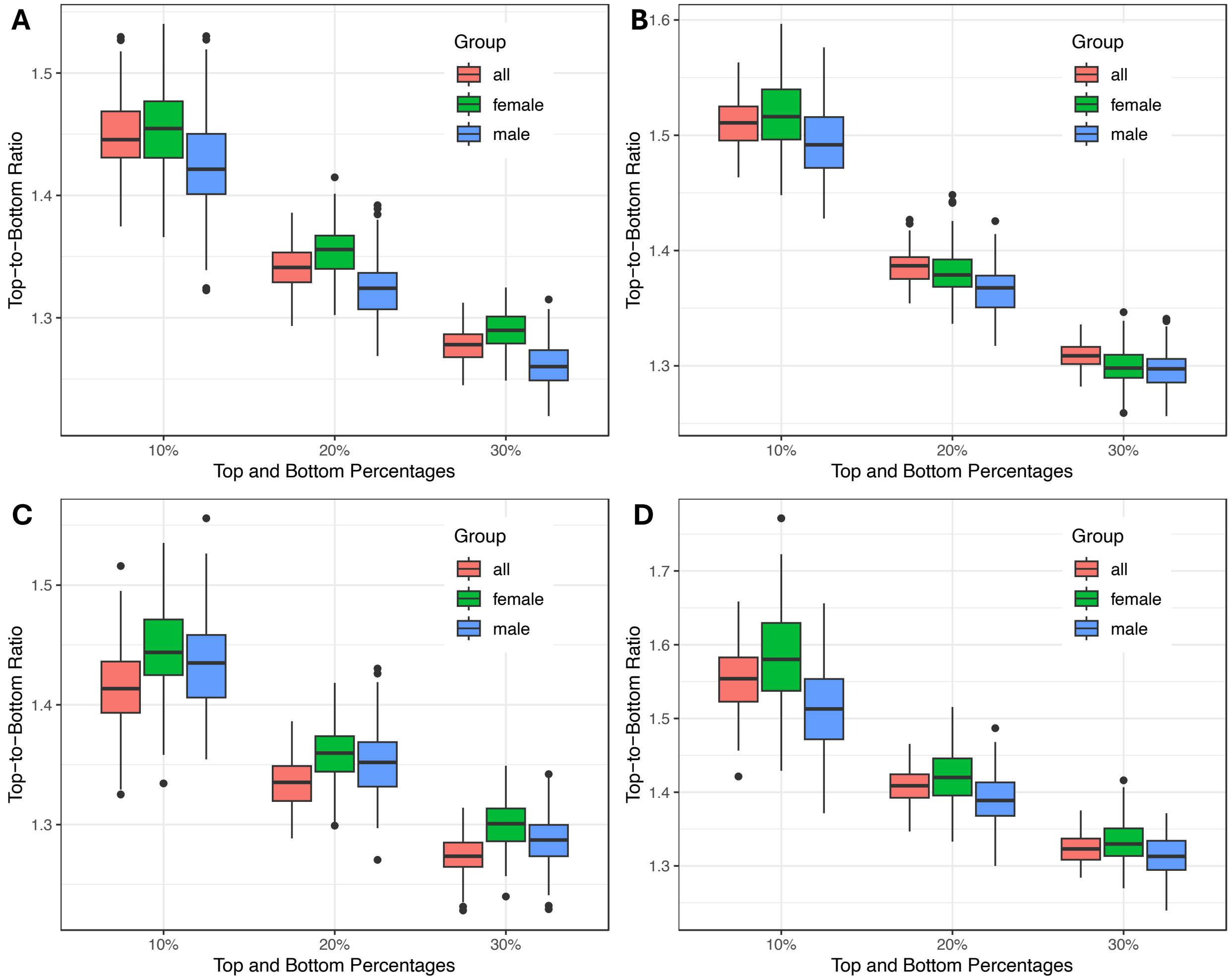


**Fig. S51 Stratification capability of plasma protein prediction models for organ-specific volume traits.**

The top-to-bottom ratios of **A** left kidney volume, **B** liver volume, **C** lung volume, and **D** pancreas volume across percentile groups representing the top and bottom 10%, 20%, and 30% of predicted imaging traits based on plasma proteins. The *x*-axis represents the percentile groups, and the *y*-axis represents the top-to-bottom ratios. Sex groups, including all individuals, females, and males, are distinguished by color.

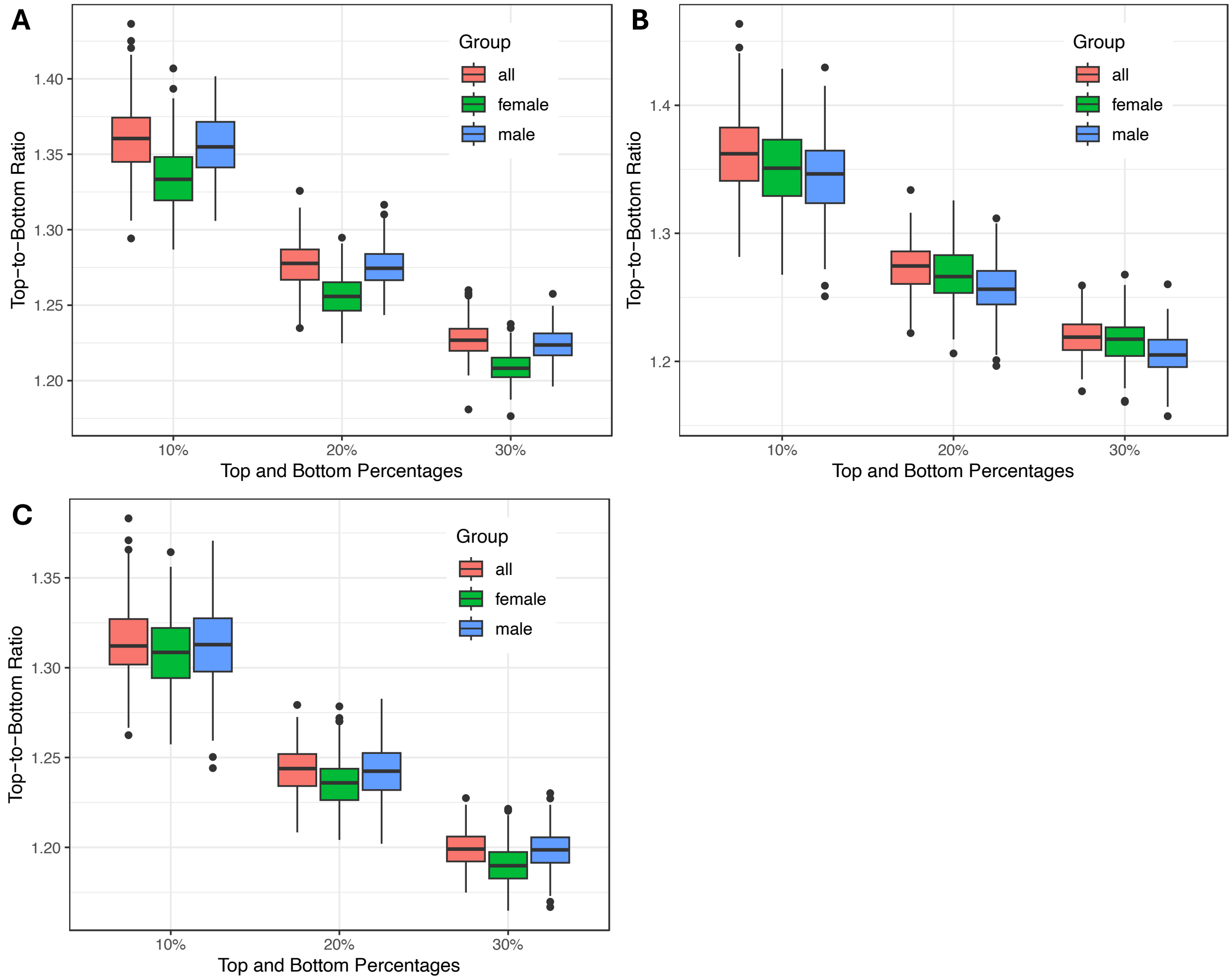


**Fig. S52 Stratification capability of plasma protein prediction models for cardiac MRI.**

The top-to-bottom ratios of **A** left ventricular myocardial mass, **B** right ventricular end-systolic volume, and **C** left ventricular end-diastolic volume across percentile groups representing the top and bottom 10%, 20%, and 30% of predicted imaging traits based on plasma proteins. The *x*-axis represents the percentile groups, and the *y*-axis represents the top-to-bottom ratios. Sex groups, including all individuals, females, and males, are distinguished by color.

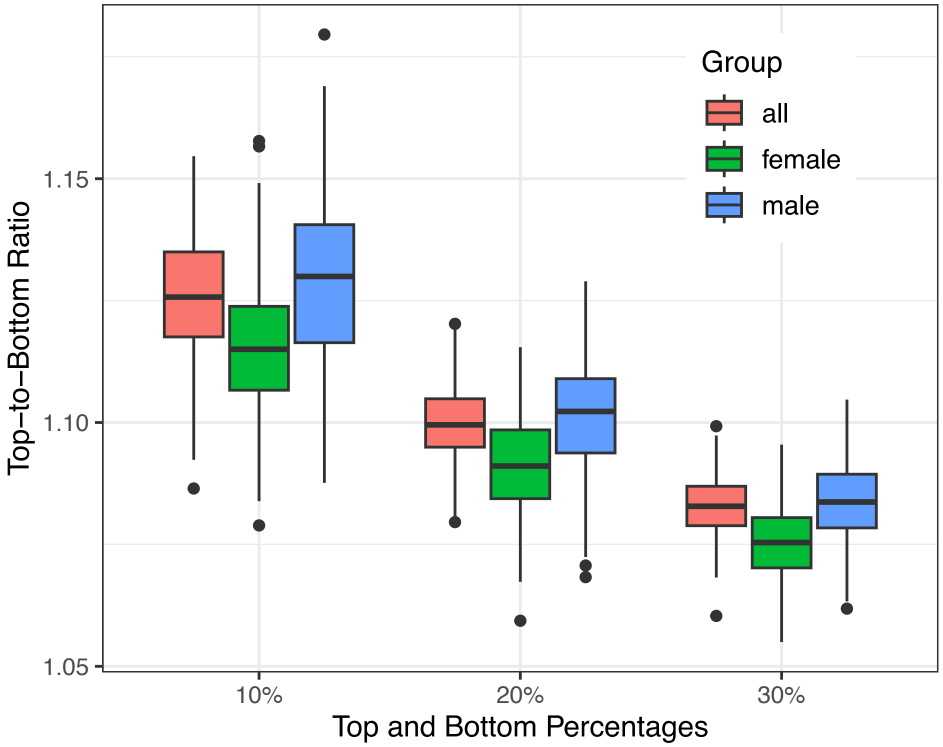


**Fig. S53 Stratification capability of plasma protein prediction models for brain structural MRI.**

The top-to-bottom ratios of the volume of cerebral white matter in the left hemisphere across percentile groups representing the top and bottom 10%, 20%, and 30% of predicted imaging traits based on plasma proteins. The *x*-axis represents the percentile groups, and the *y*-axis represents the top-to-bottom ratios. Sex groups, including all individuals, females, and males, are distinguished by color.

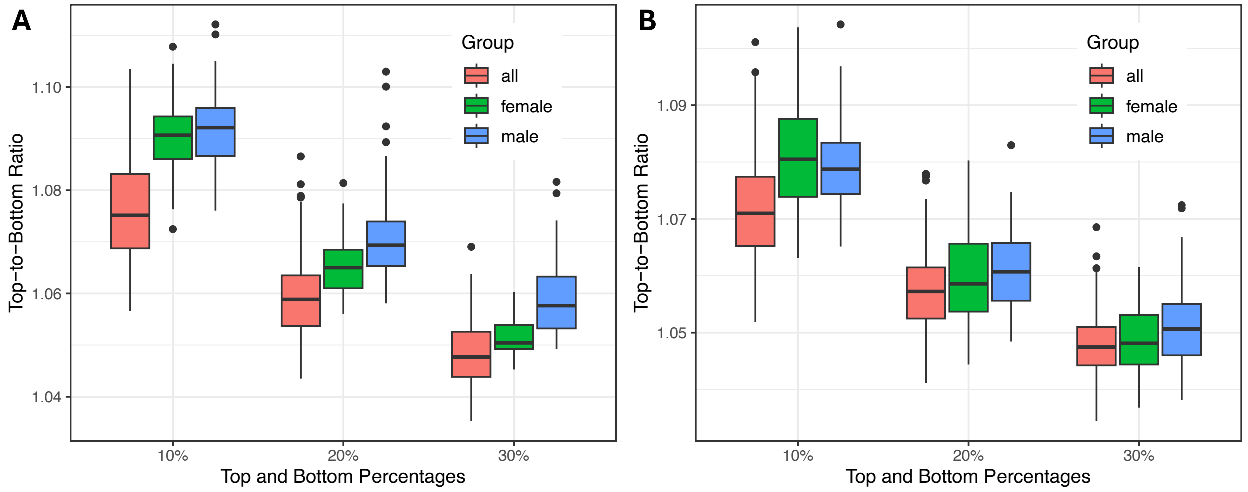


**Fig. S54 Stratification capability of plasma protein prediction models for brain structural MRI.**

The top-to-bottom ratios of **A** the volume of grey matter in the right amygdala and **B** the volume of grey matter in the left insular cortex across percentile groups representing the top and bottom 10%, 20%, and 30% of predicted imaging traits based on plasma proteins. The *x*-axis represents the percentile groups, and the *y*-axis represents the top-to-bottom ratios. Sex groups, including all individuals, females, and males, are distinguished by color.

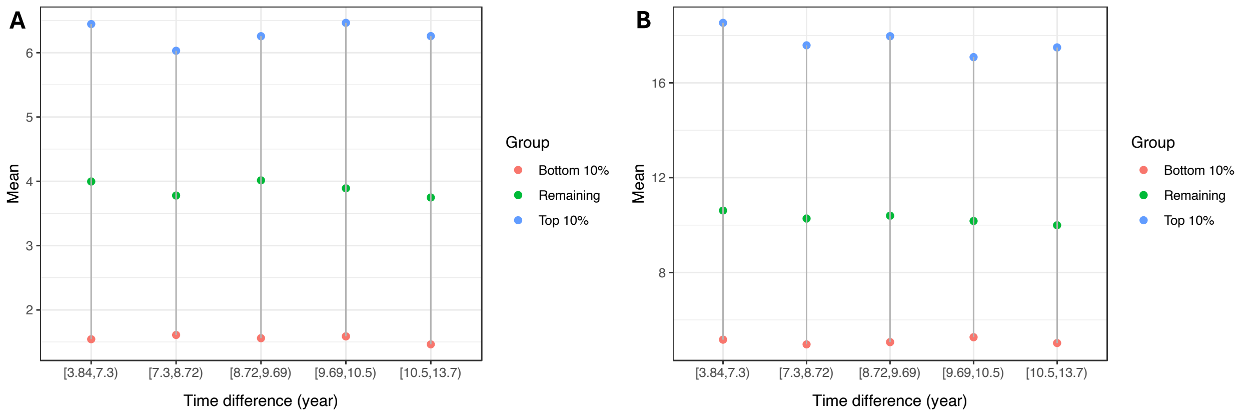


**Fig. S55 Stratification capability of plasma protein prediction models across time difference groups.**

The time difference between blood sample collection and imaging scans (*x*-axis) and the mean values of **A** visceral fat volume and **B** total trunk fat volume (*y*-axis) for the top 10%, bottom 10%, and the remaining individuals based on predicted values from plasma proteins. Colors represent the groups: top 10%, bottom 10%, or the remaining individuals.

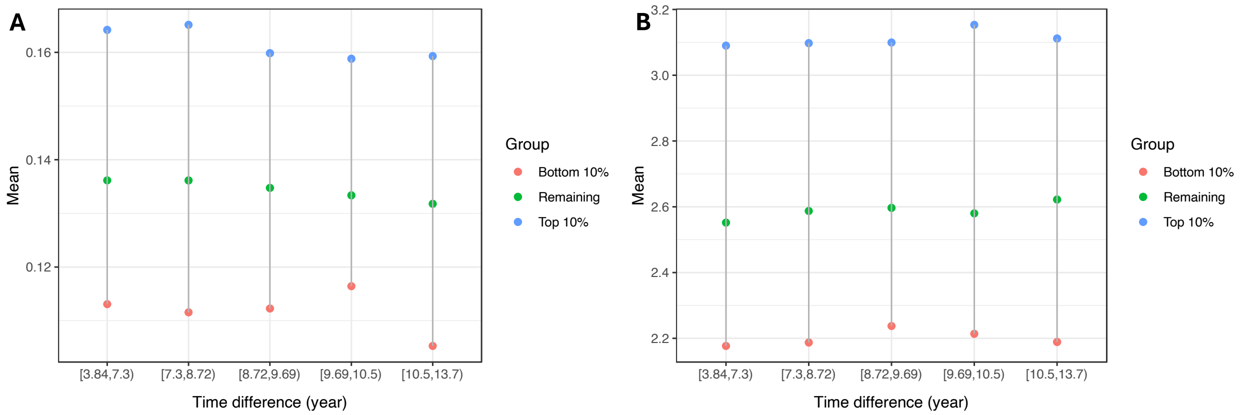


**Fig. S56 Stratification capability of plasma protein prediction models across time difference groups.**

The time difference between blood sample collection and imaging scans (*x*-axis) and the mean values of **A** left kidney volume and **B** lung volume (*y*-axis) for the top 10%, bottom 10%, and the remaining individuals based on predicted values from plasma proteins. Colors represent the groups: top 10%, bottom 10%, or the remaining individuals.

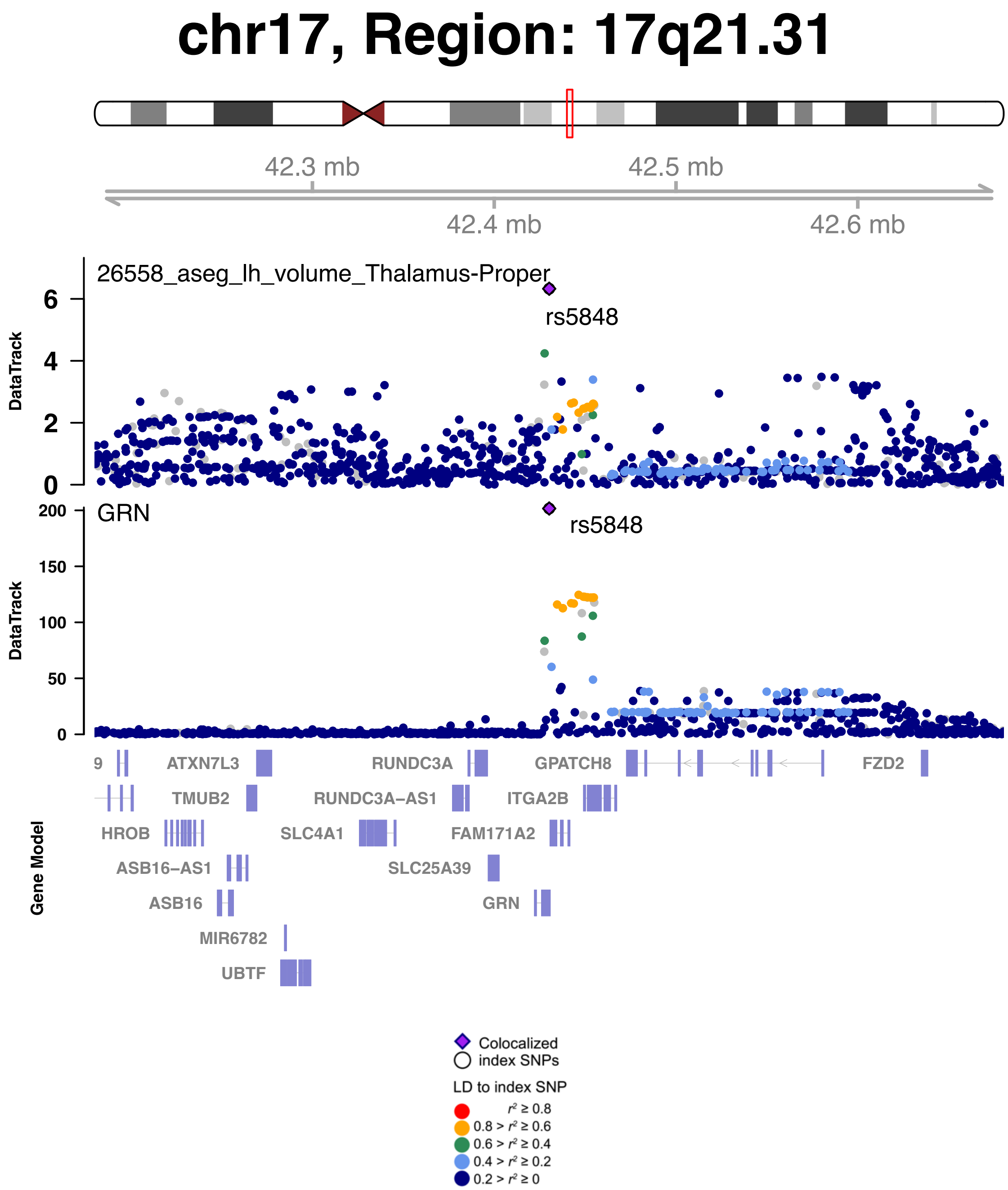


**Fig. S57 Genetic causal pairs between plasma proteins and brain structural MRI identified through Mendelian randomization.**

GRN was causally associated with volume of the thalamus in the left hemisphere. The posterior probability of Bayesian colocalization analysis for the shared causal variant hypothesis (PPH4) is 99.21%.

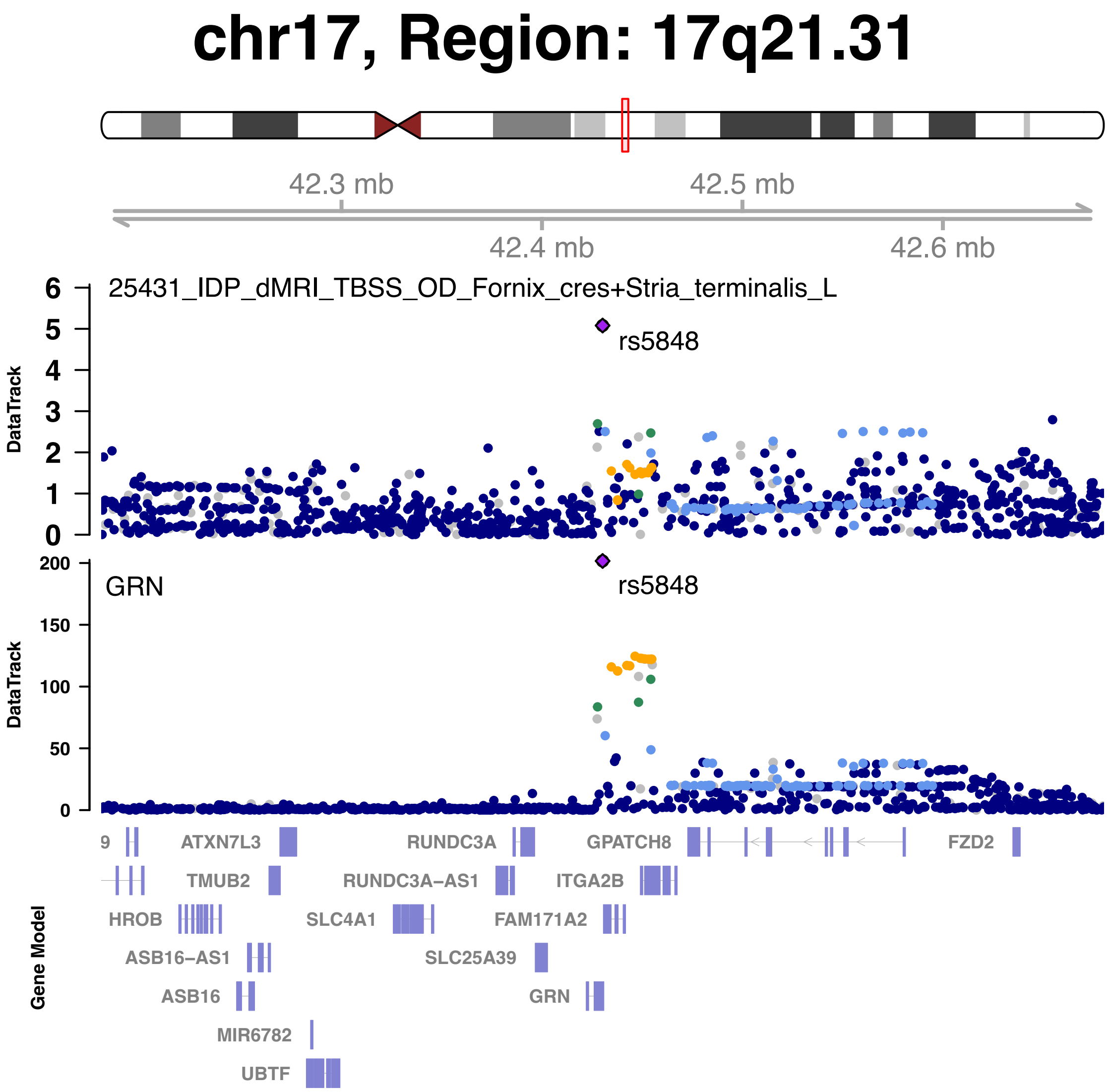


**Fig. S58 Genetic causal pairs between plasma proteins and brain diffusion MRI identified through Mendelian randomization.**

GRN was causally associated with mean orientation dispersion index (OD) in the left fornix cres+stria terminalis. The posterior probability of Bayesian colocalization analysis for the shared causal variant hypothesis (PPH4) is 97.53%.

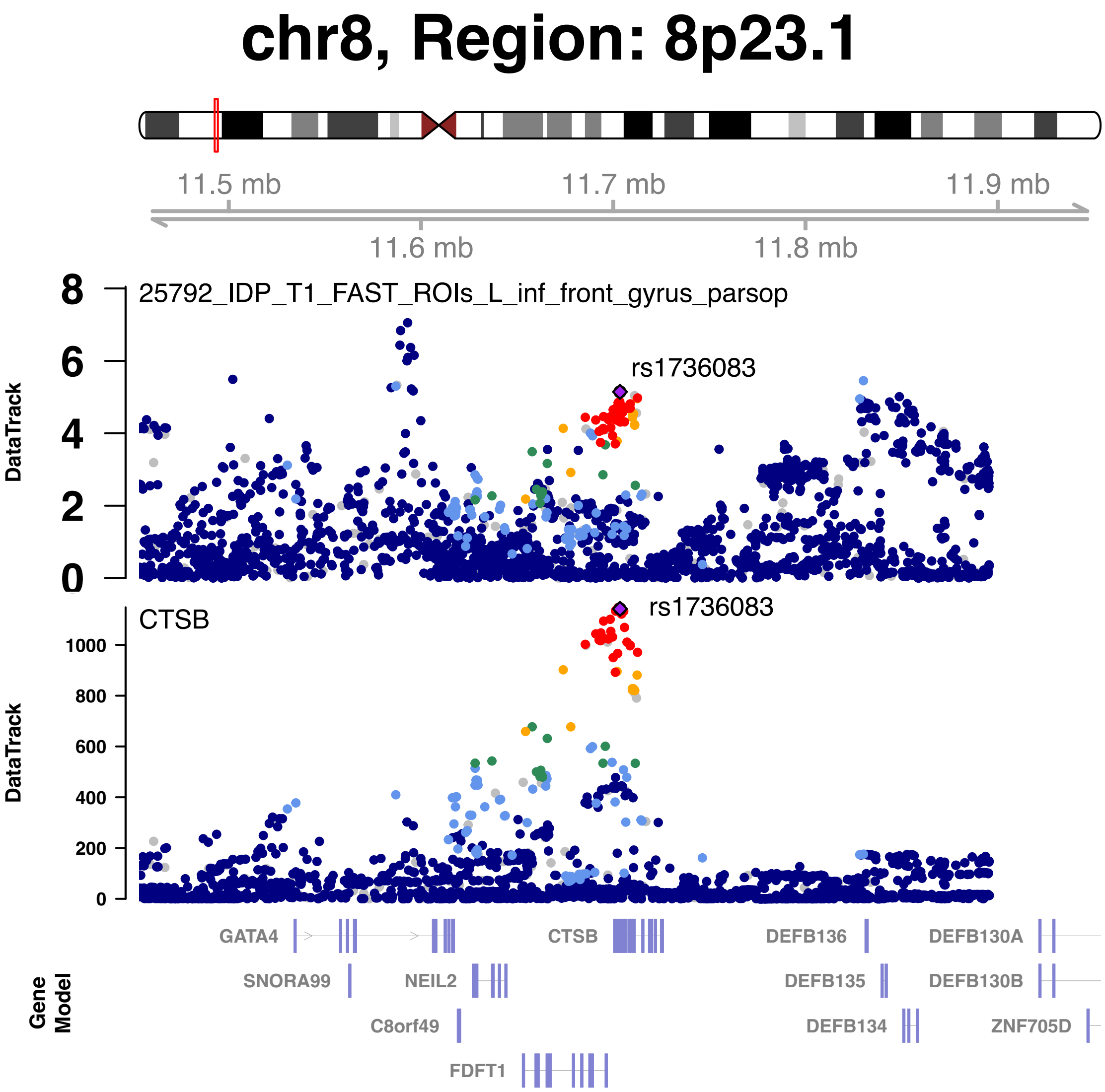


**Fig. S59 Genetic causal pairs between plasma proteins and brain structural MRI identified through Mendelian randomization.**

CTSB was causally associated with the volume of grey matter in the left inferior frontal gyrus, pars opercularis. The posterior probability of Bayesian colocalization analysis for the shared causal variant hypothesis (PPH4) is 80.00%.

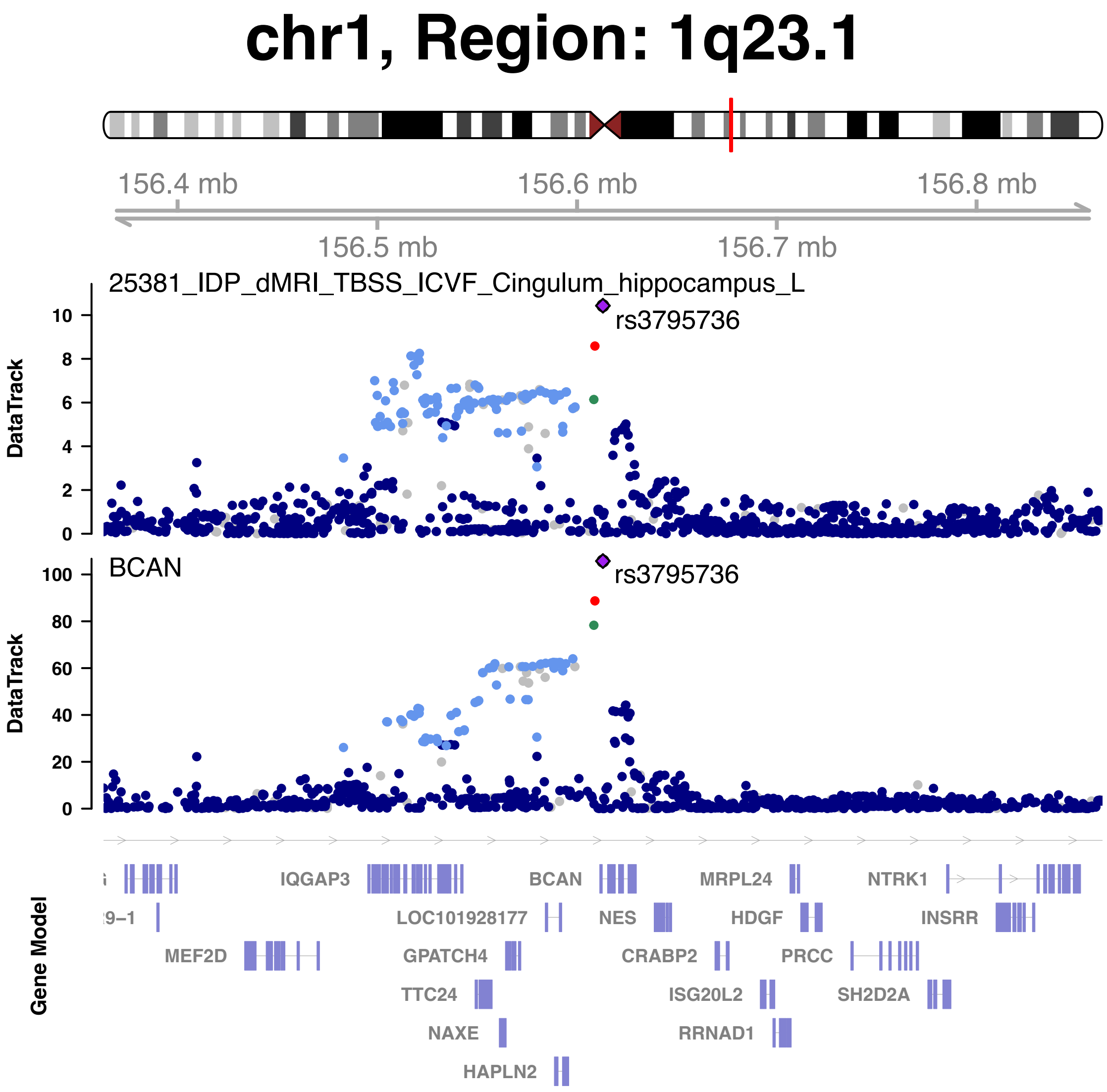


**Fig. S60 Genetic causal pairs between plasma proteins and brain diffusion MRI identified through Mendelian randomization.**

BCAN was causally associated with mean intracellular volume fraction (ICVF) in the left cingulum hippocampus. The posterior probability of Bayesian colocalization analysis for the shared causal variant hypothesis (PPH4) is 99.99%.

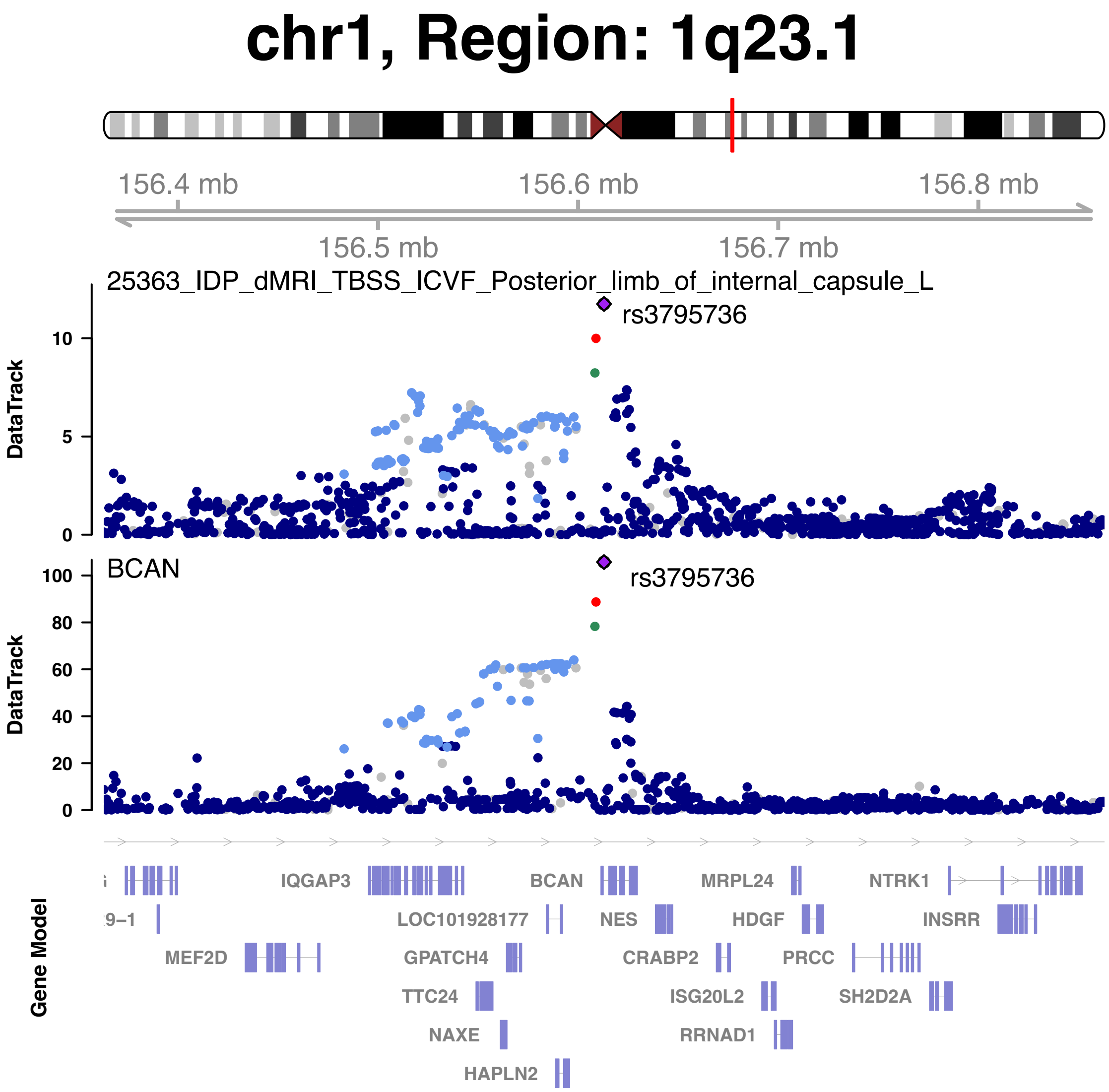


**Fig. S61 Genetic causal pairs between plasma proteins and brain diffusion MRI identified through Mendelian randomization.**

BCAN was causally associated with mean intracellular volume fraction (ICVF) in the posterior limb of the left internal capsule. The posterior probability of Bayesian colocalization analysis for the shared causal variant hypothesis (PPH4) is 100%.

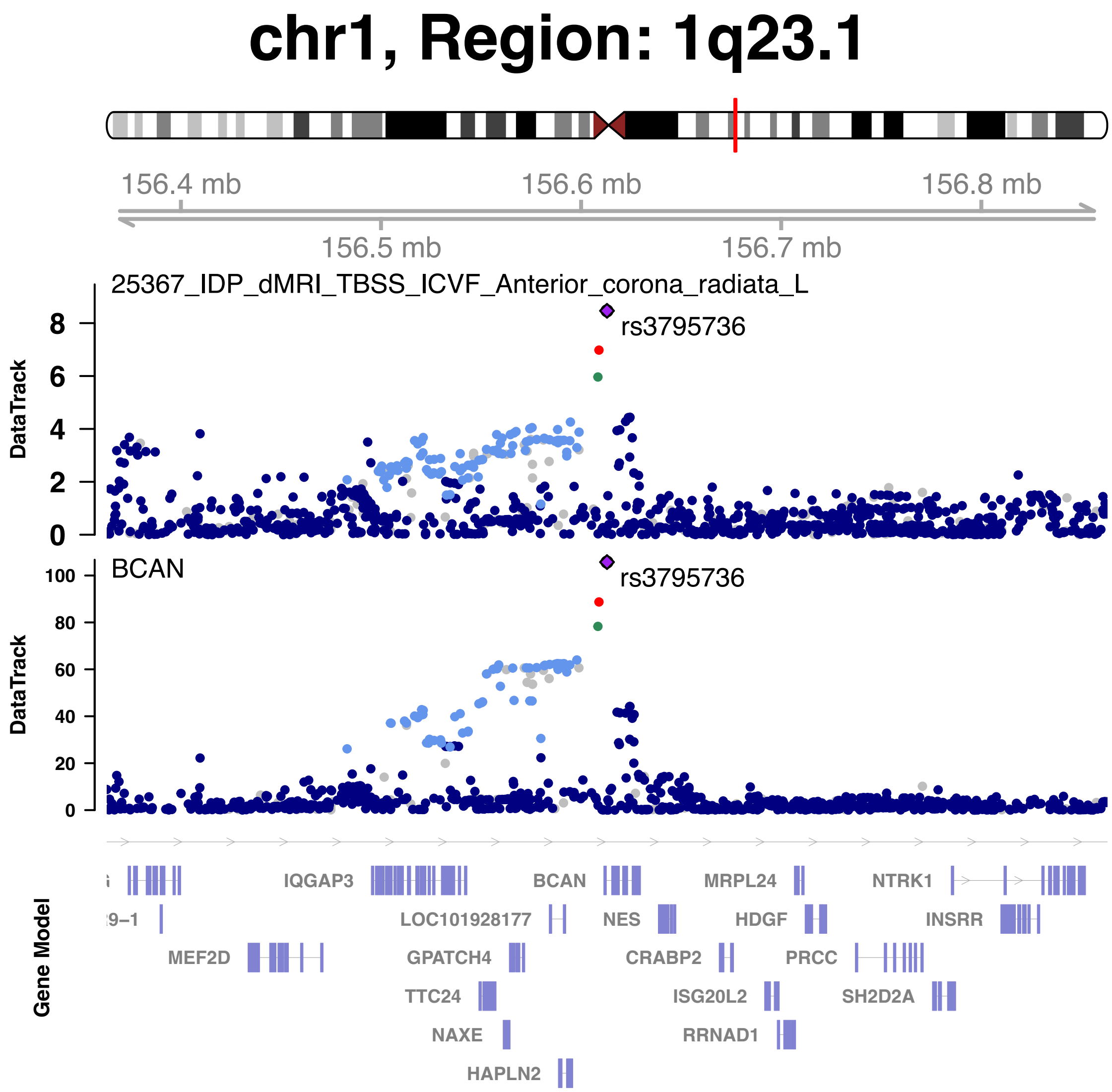


**Fig. S62 Genetic causal pairs between plasma proteins and brain diffusion MRI identified through Mendelian randomization.**

BCAN was causally associated with mean intracellular volume fraction (ICVF) in the left anterior corona radiata. The posterior probability of Bayesian colocalization analysis for the shared causal variant hypothesis (PPH4) is 99.99%.

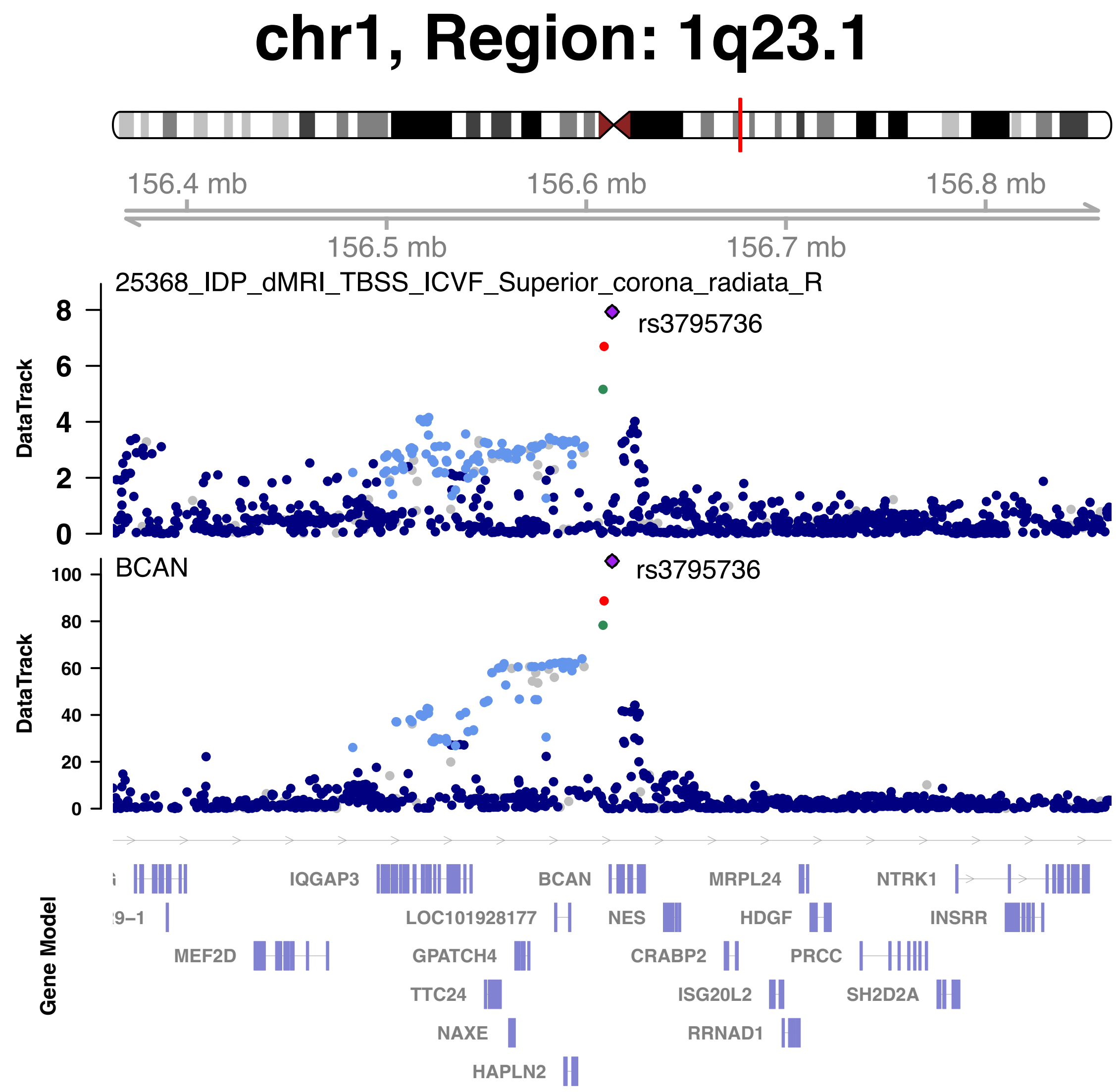


**Fig. S63 Genetic causal pairs between plasma proteins and brain diffusion MRI identified through Mendelian randomization.**

BCAN was causally associated with mean intracellular volume fraction (ICVF) in the right superior corona radiata. The posterior probability of Bayesian colocalization analysis for the shared causal variant hypothesis (PPH4) is 99.99%.

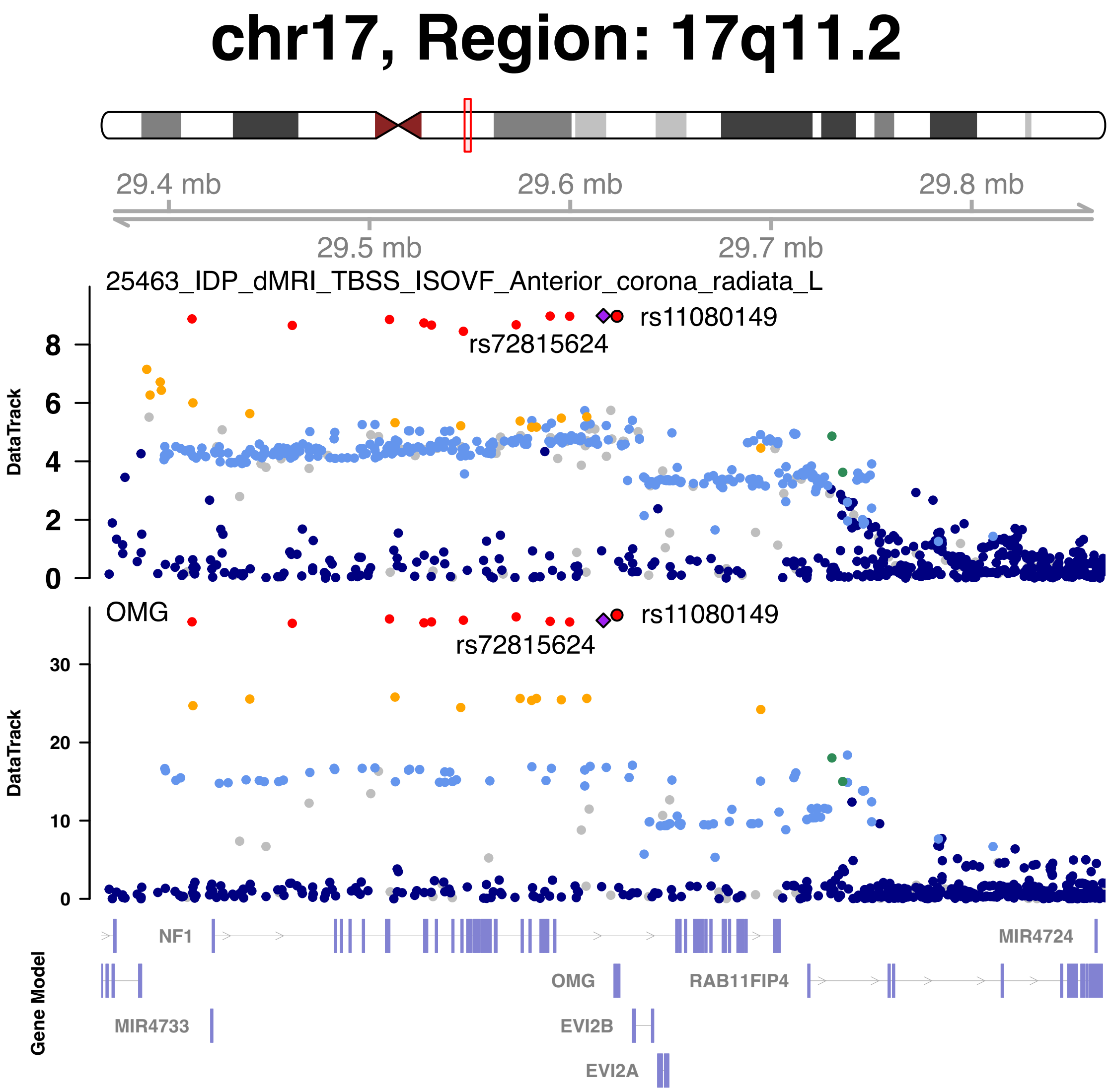


**Fig. S64 Genetic causal pairs between plasma proteins and brain diffusion MRI identified through Mendelian randomization.**

OMG was causally associated with mean isotropic or free water volume fraction (ISOVF) in the left anterior corona radiata. The posterior probability of Bayesian colocalization analysis for the shared causal variant hypothesis (PPH4) is 99.18%.

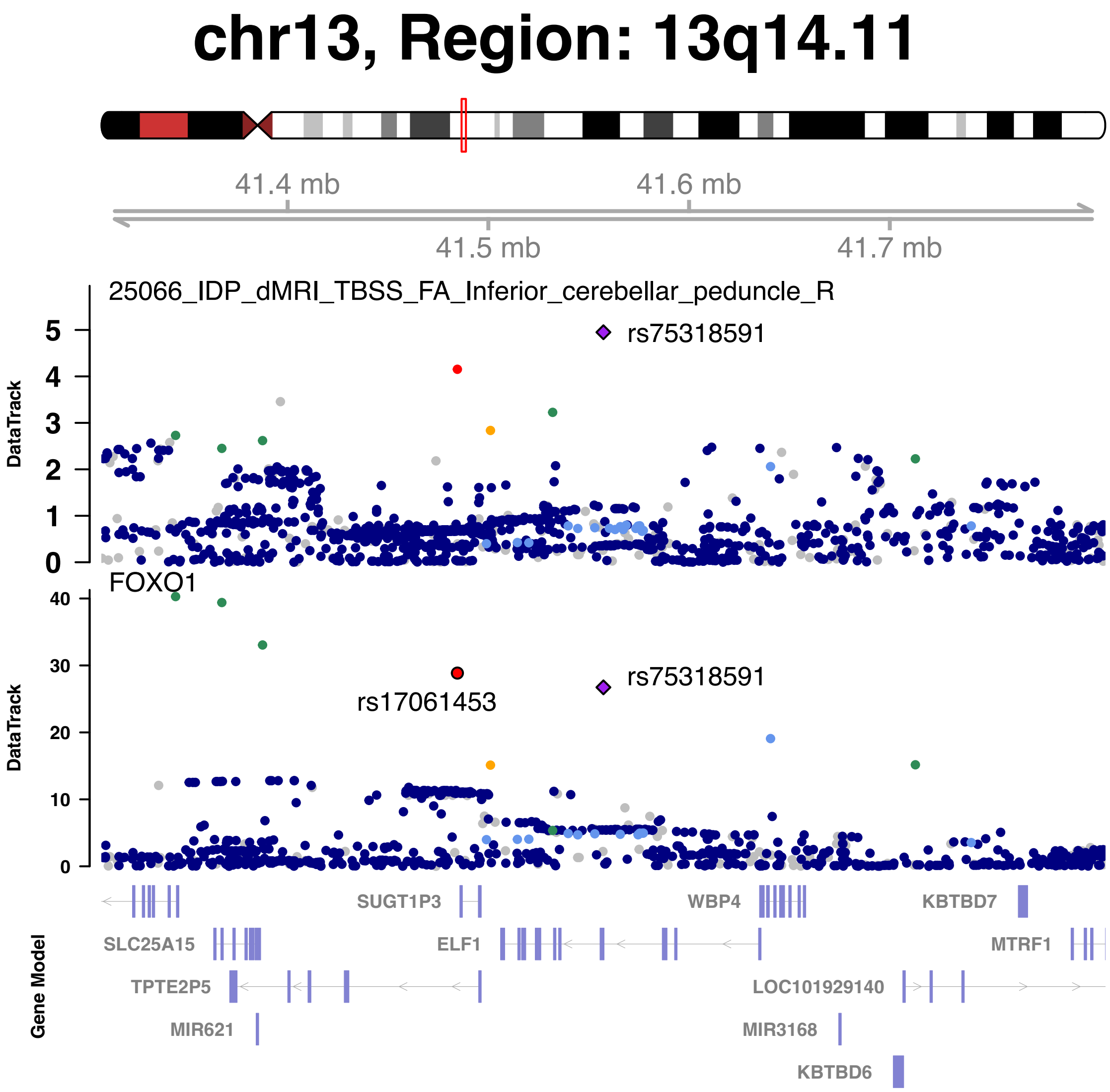


**Fig. S65 Genetic causal pairs between plasma proteins and brain diffusion MRI identified through Mendelian randomization.**

FOXO1 was causally associated with mean fractional anisotropy (FA) in the right inferior cerebellar peduncle. The posterior probability of Bayesian colocalization analysis for the shared causal variant hypothesis (PPH4) is 61.94%.

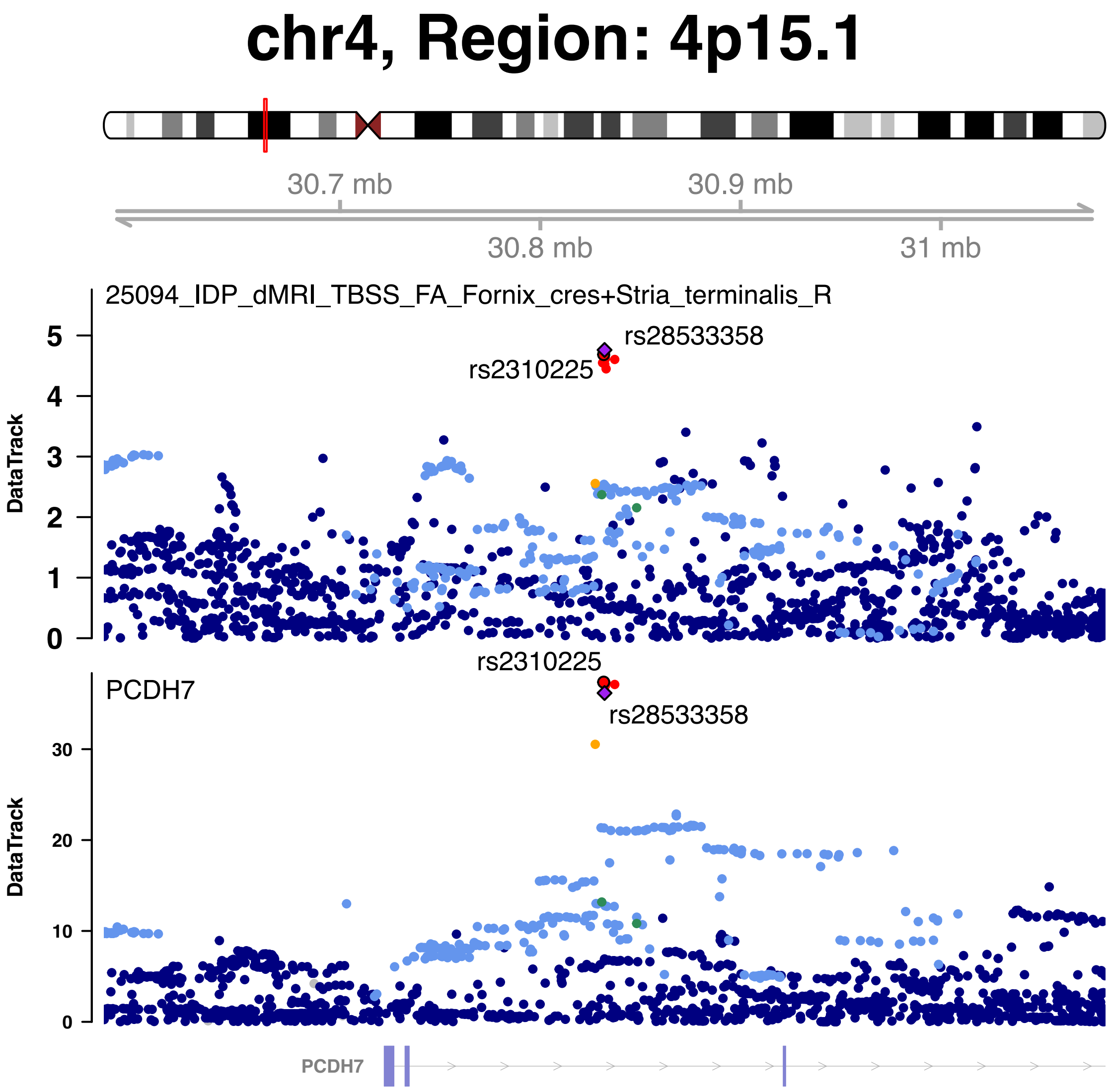


**Fig. S66 Genetic causal pairs between plasma proteins and brain diffusion MRI identified through Mendelian randomization.**

PCDH7 was causally associated with mean fractional anisotropy (FA) in the right fornix cres and stria terminalis. The posterior probability of Bayesian colocalization analysis for the shared causal variant hypothesis (PPH4) is 95.66%.

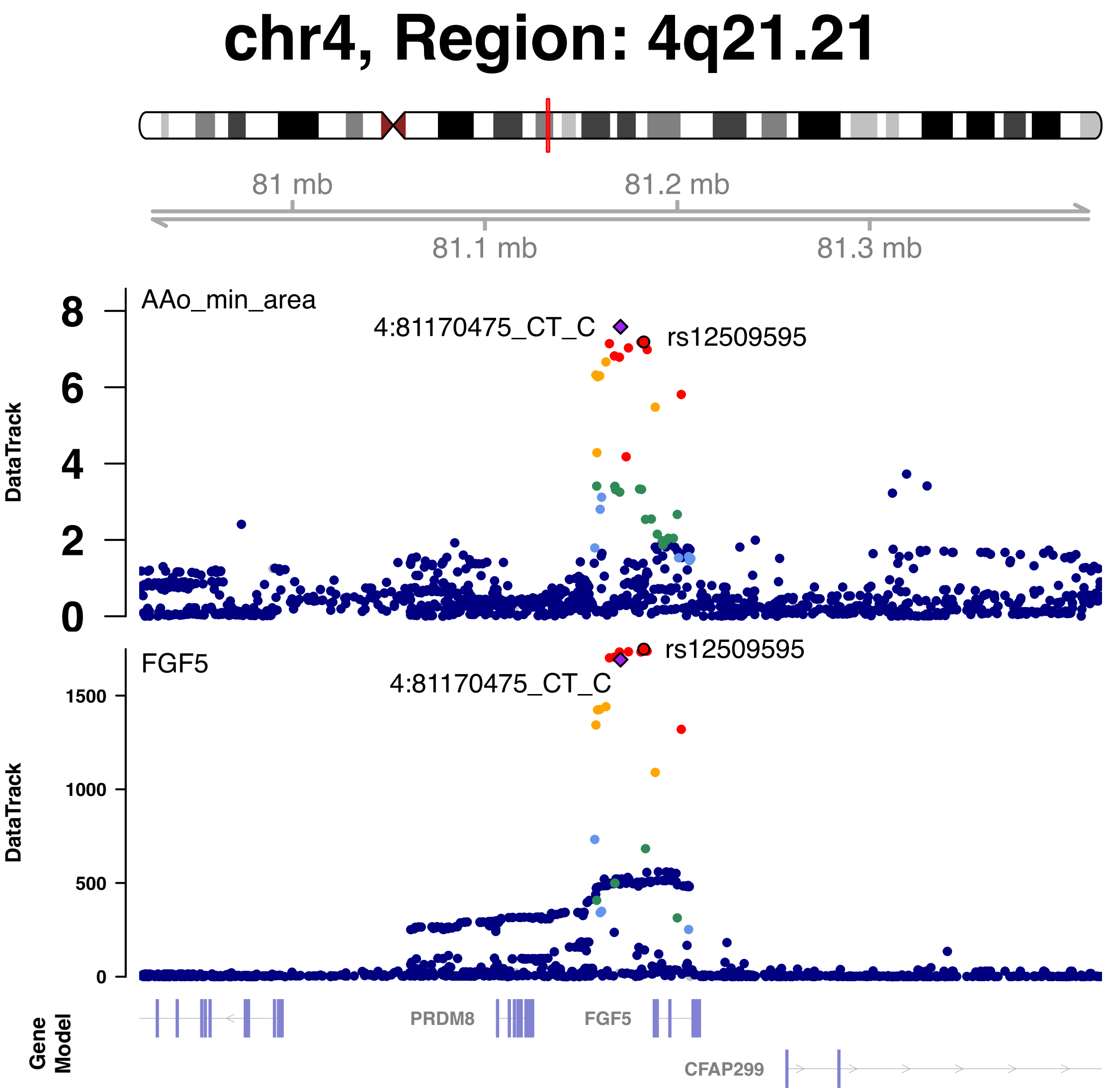


**Fig. S67 Genetic causal pairs between plasma proteins and aortic MRI identified through Mendelian randomization.**

FGF5 was causally associated with ascending aorta minimum area (AAo min area). The posterior probability of Bayesian colocalization analysis for the shared causal variant hypothesis (PPH4) is 99.26%.

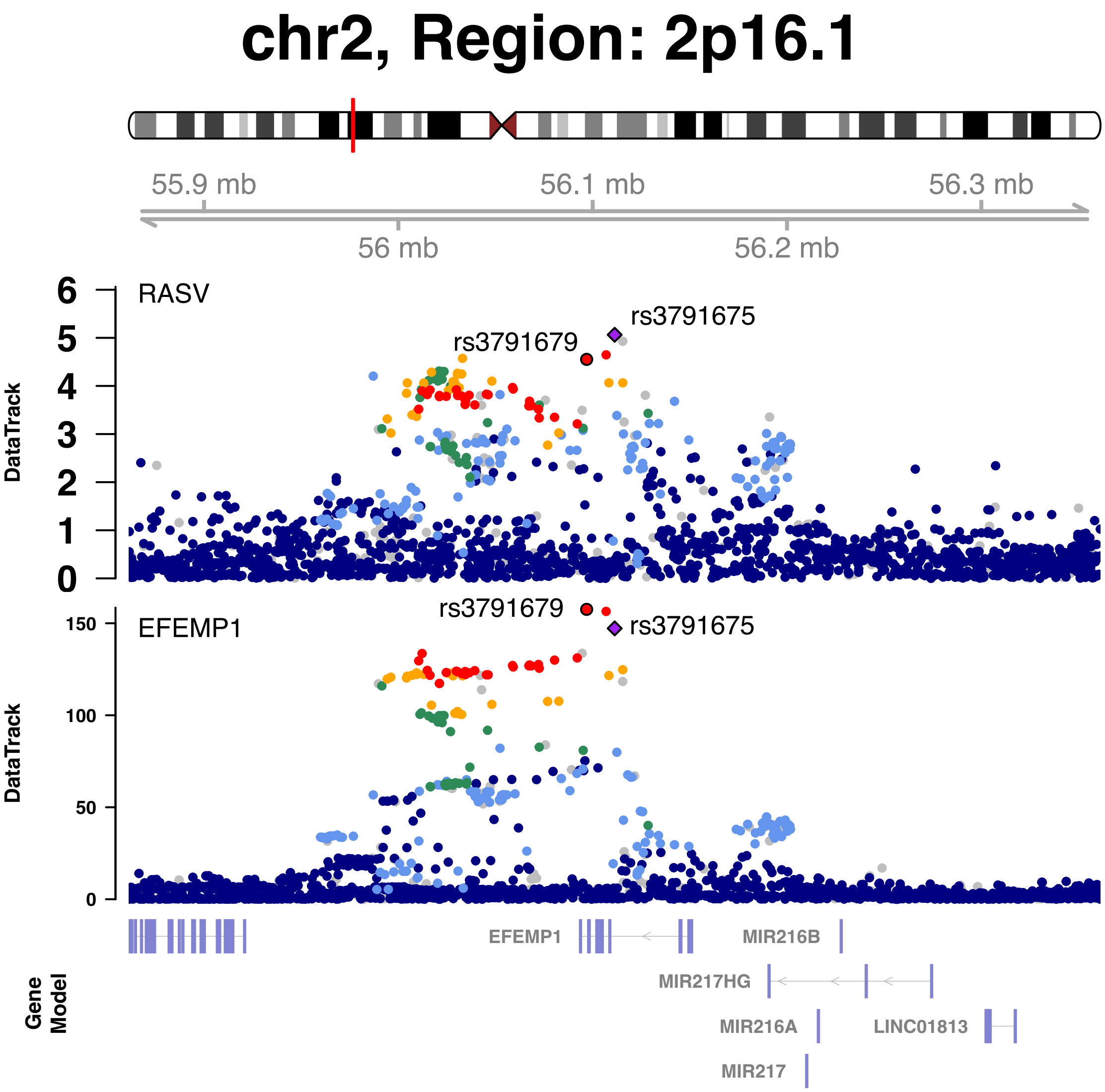


**Fig. S68 Genetic causal pairs between plasma proteins and cardiac MRI identified through Mendelian randomization.**

EFEMP1 was causally associated with right atrium stroke volume (RASV). The posterior probability of Bayesian colocalization analysis for the shared causal variant hypothesis (PPH4) is 93.57%.

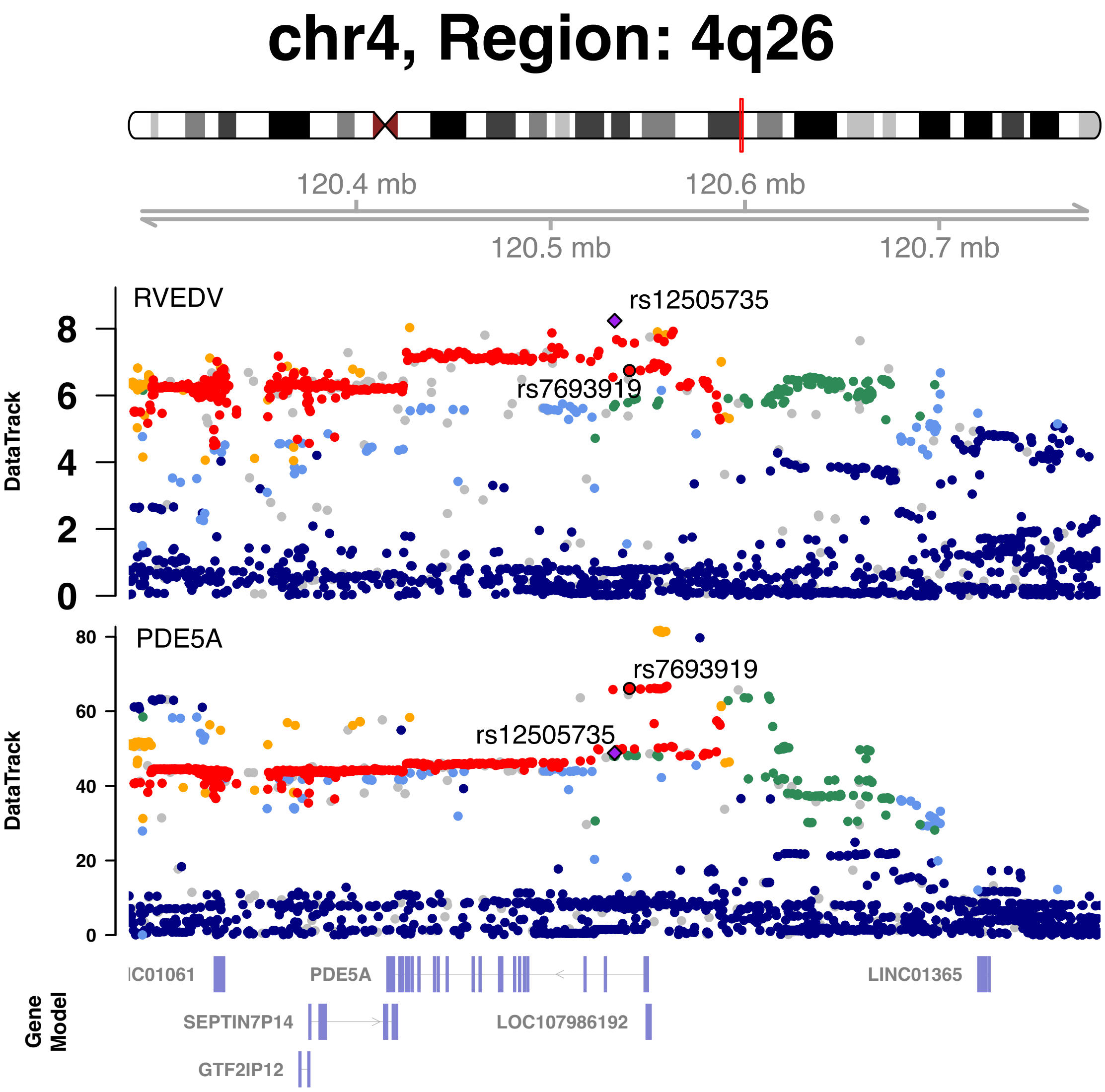


**Fig. S69 Genetic causal pairs between plasma proteins and cardiac MRI identified through Mendelian randomization.**

PDE5A was causally associated with right ventricular end-diastolic volume (RVEDV). The posterior probability of Bayesian colocalization analysis for the shared causal variant hypothesis (PPH4) is 92.62%.

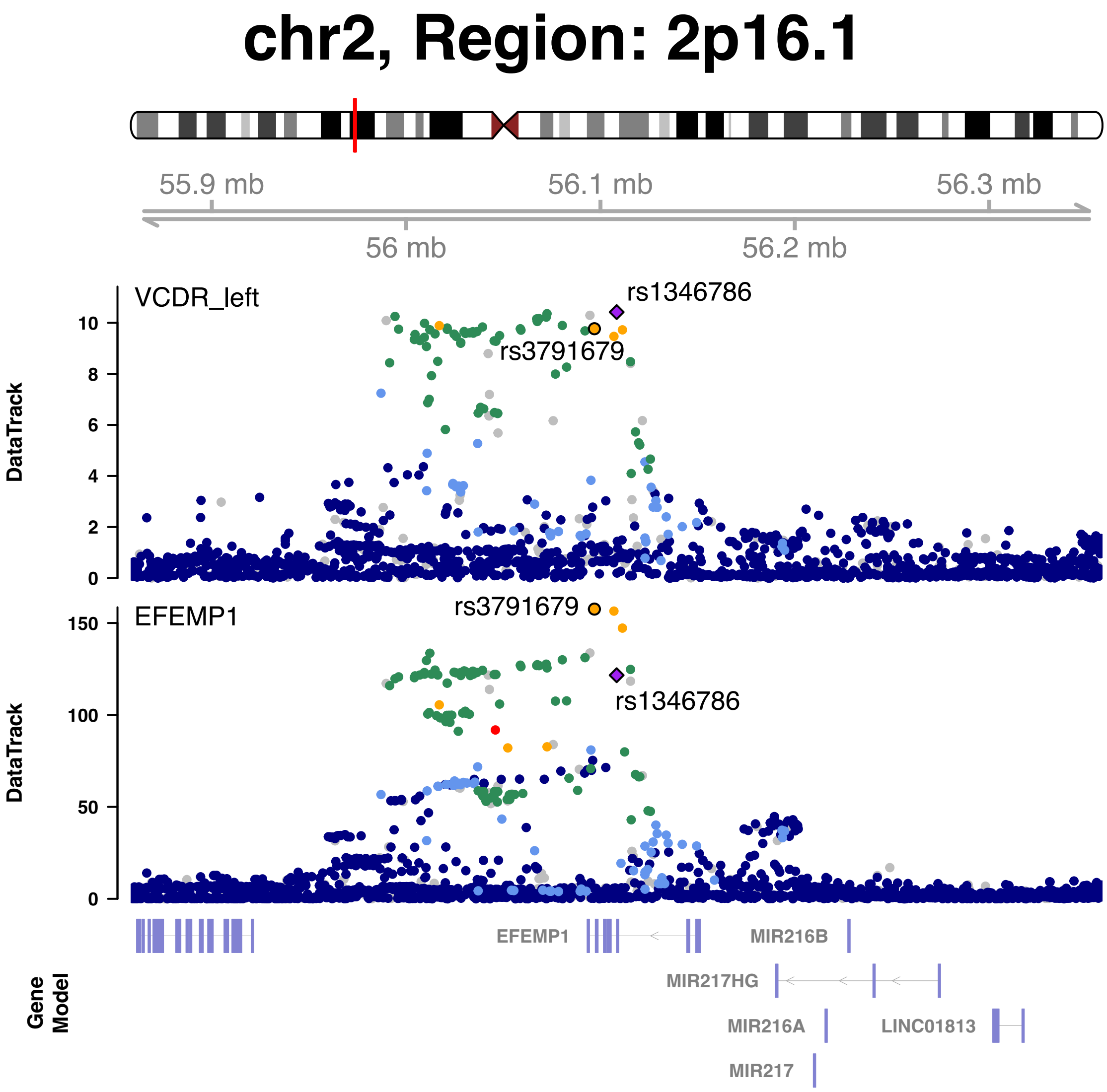


**Fig. S70 Genetic causal pairs between plasma proteins and eye imaging identified through Mendelian randomization.**

EFEMP1 was causally associated with the left vertical cup to disc ratio (VCDR). The posterior probability of Bayesian colocalization analysis for the shared causal variant hypothesis (PPH4) is 95.01%.

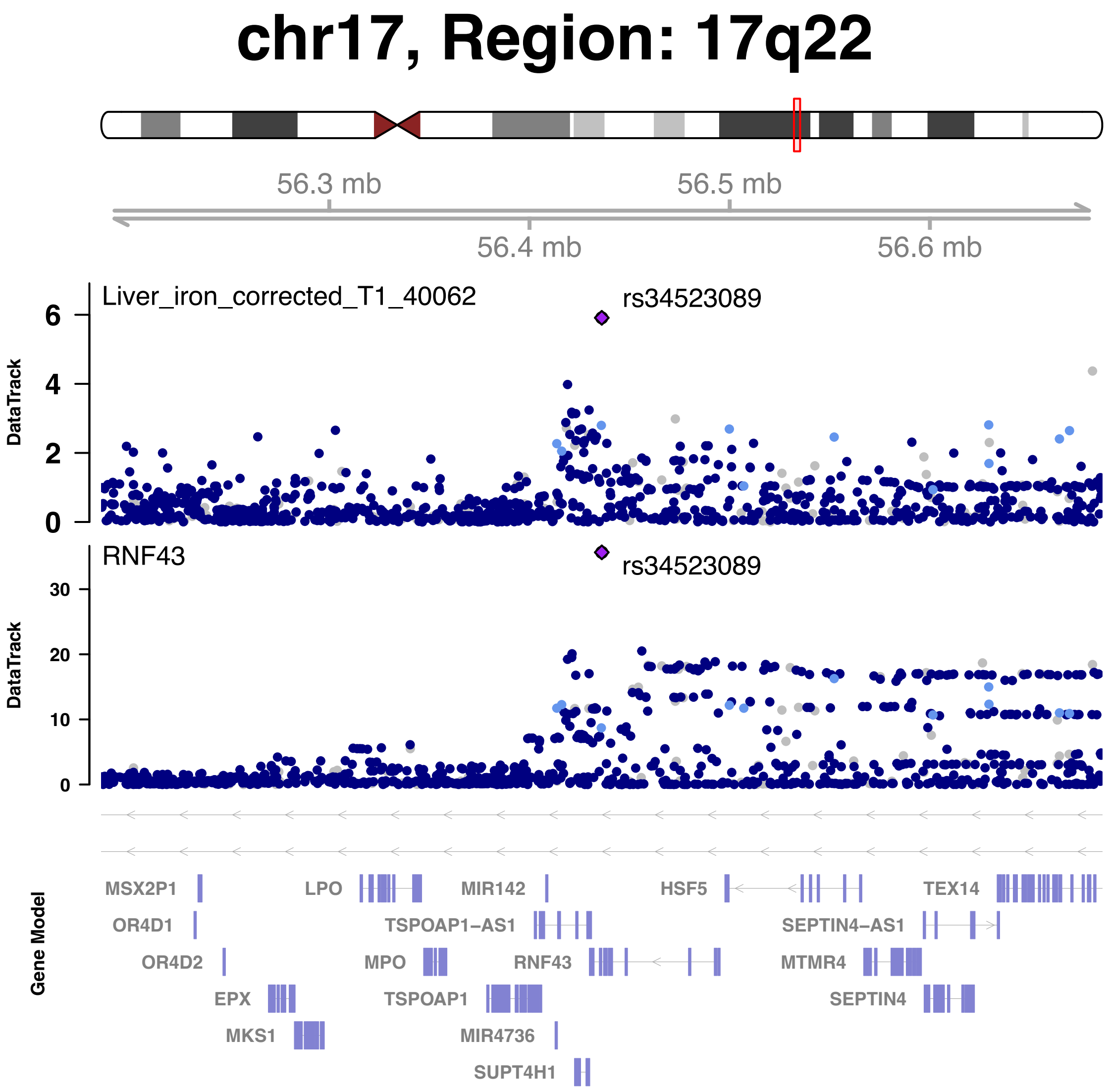


**Fig. S71 Genetic causal pairs between plasma proteins and abdominal MRI identified through Mendelian randomization.**

RNF43 was causally associated with liver iron corrected T1. The posterior probability of Bayesian colocalization analysis for the shared causal variant hypothesis (PPH4) is 99.84%.

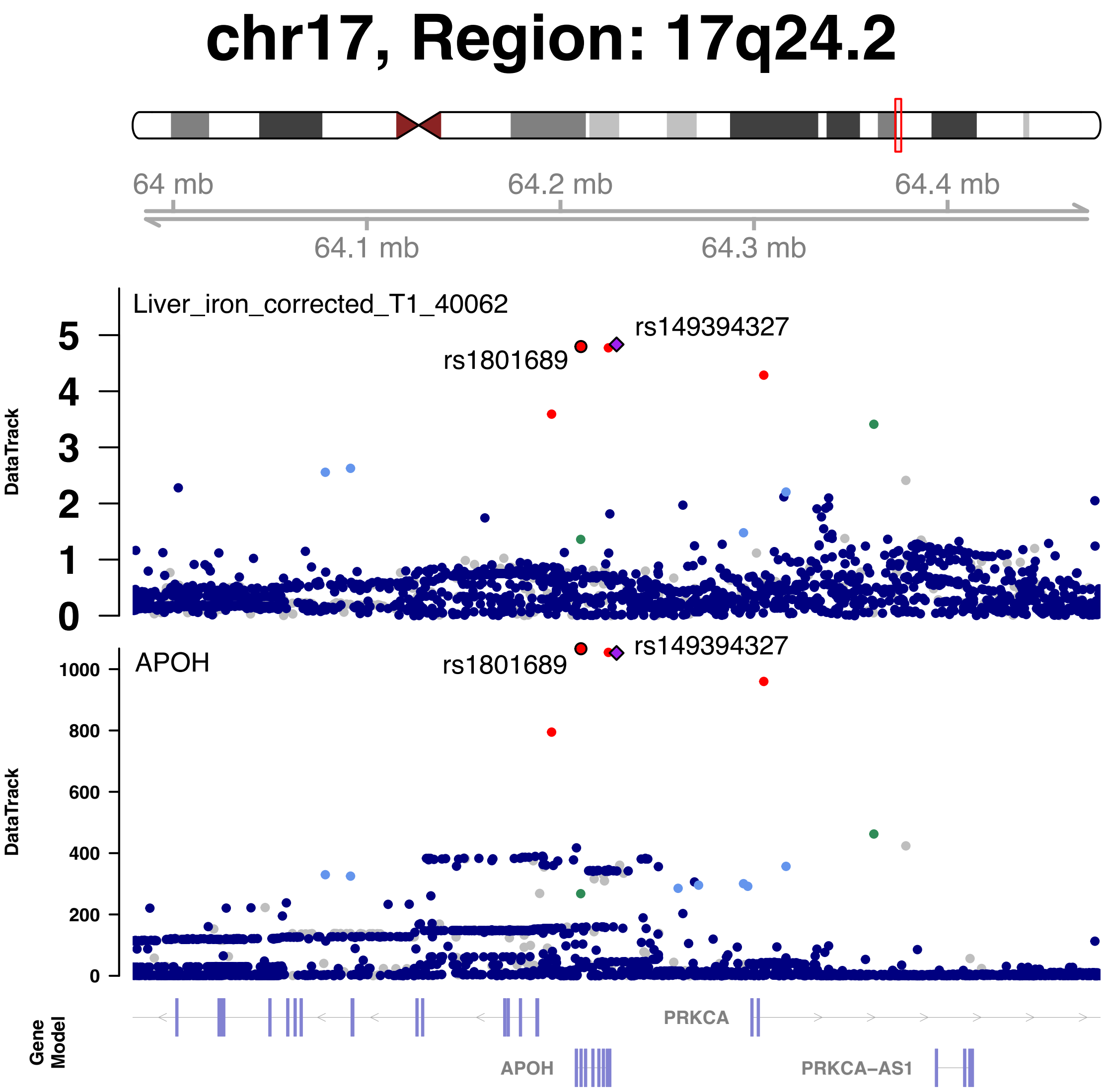


**Fig. S72 Genetic causal pairs between plasma proteins and abdominal MRI identified through Mendelian randomization.**

APOH was causally associated with liver iron corrected T1. The posterior probability of Bayesian colocalization analysis for the shared causal variant hypothesis (PPH4) is 98.87%.

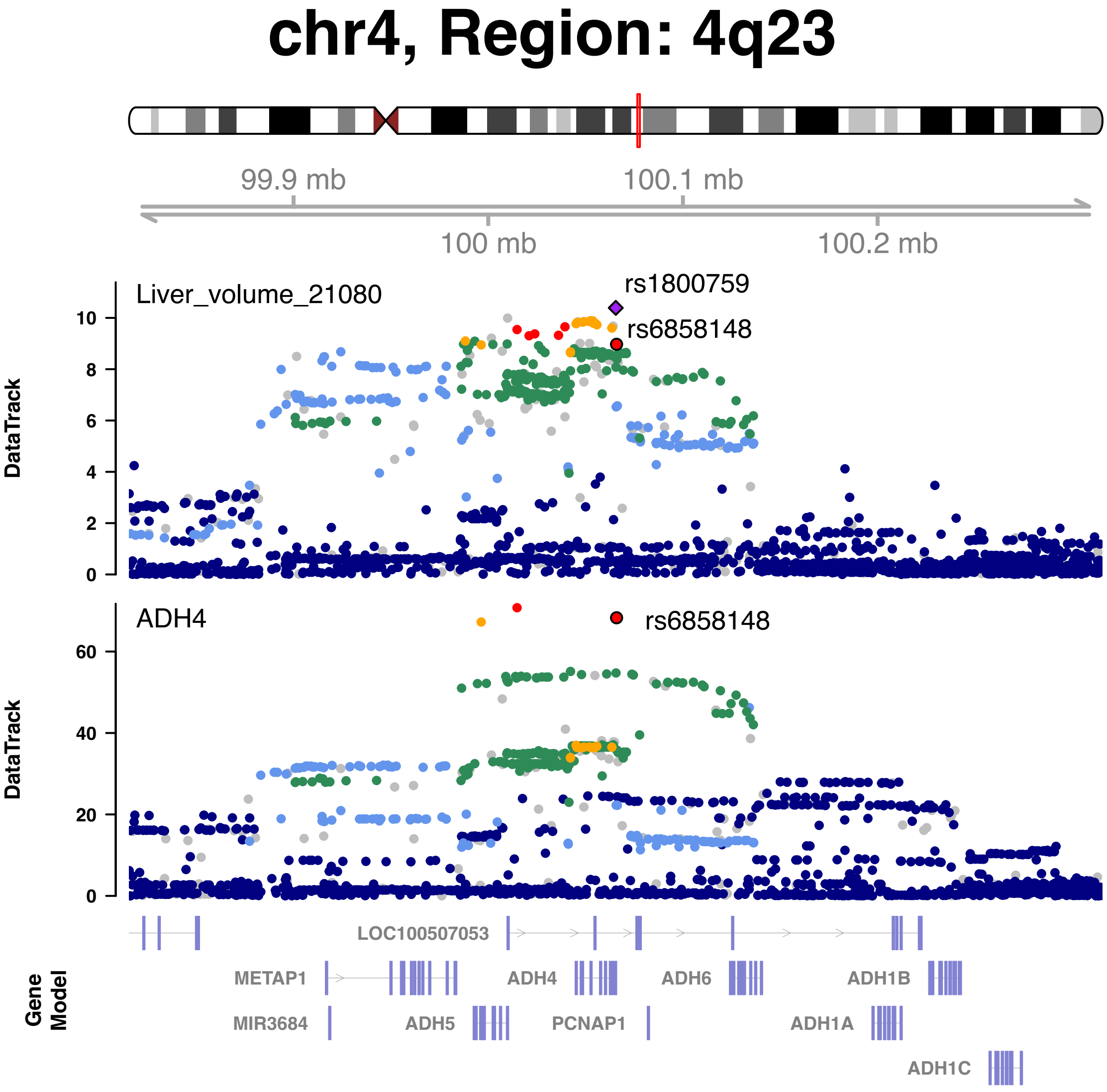


**Fig. S73 Genetic causal pairs between plasma proteins and abdominal MRI identified through Mendelian randomization.**

ADH4 was causally associated with liver volume. The posterior probability of Bayesian colocalization analysis for the shared causal variant hypothesis (PPH4) is 95.08%.

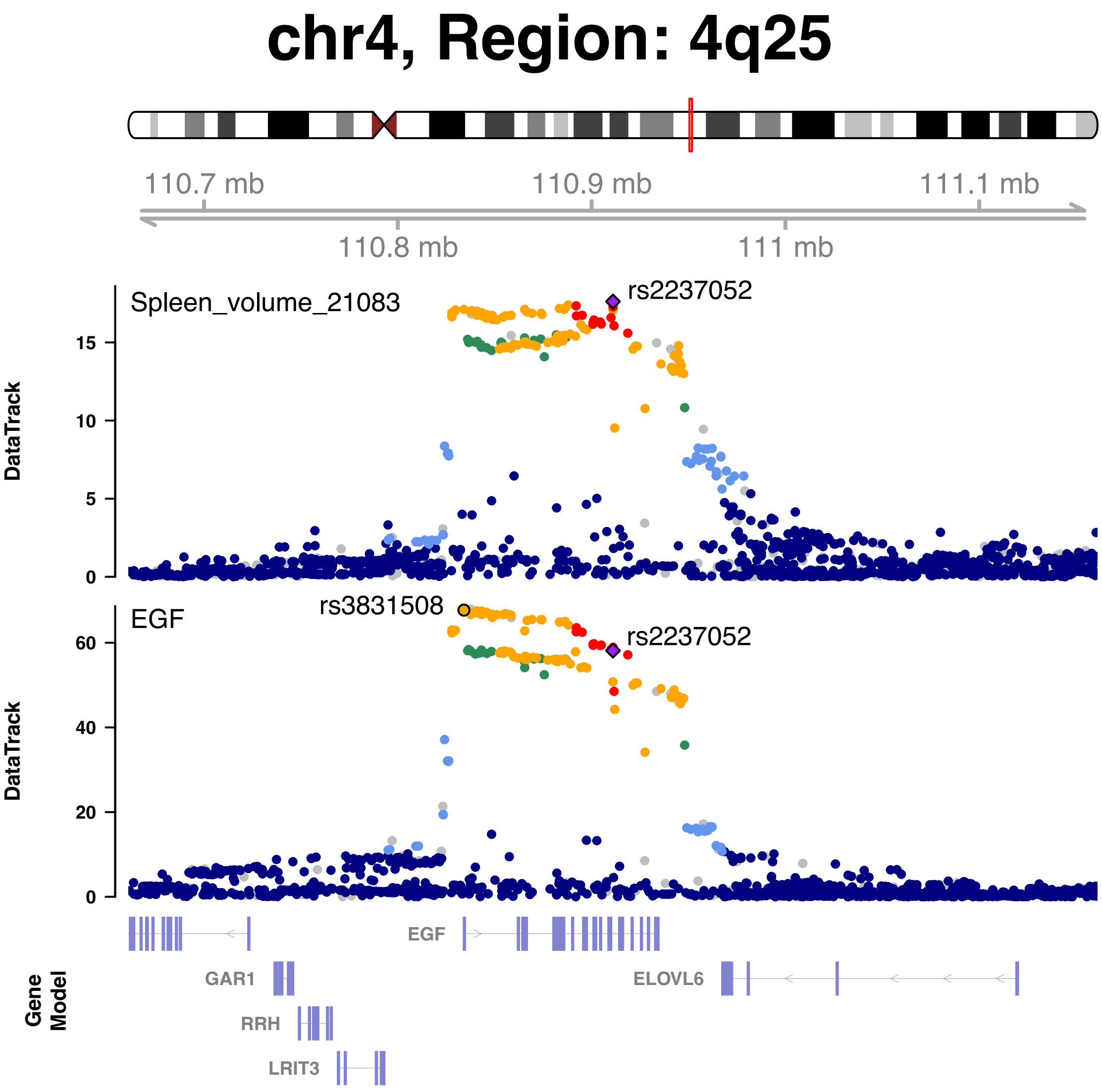


**Fig. S74 Genetic causal pairs between plasma proteins and abdominal MRI identified through Mendelian randomization.**

EGF was causally associated with spleen volume. The posterior probability of Bayesian colocalization analysis for the shared causal variant hypothesis (PPH4) is 94.52%.

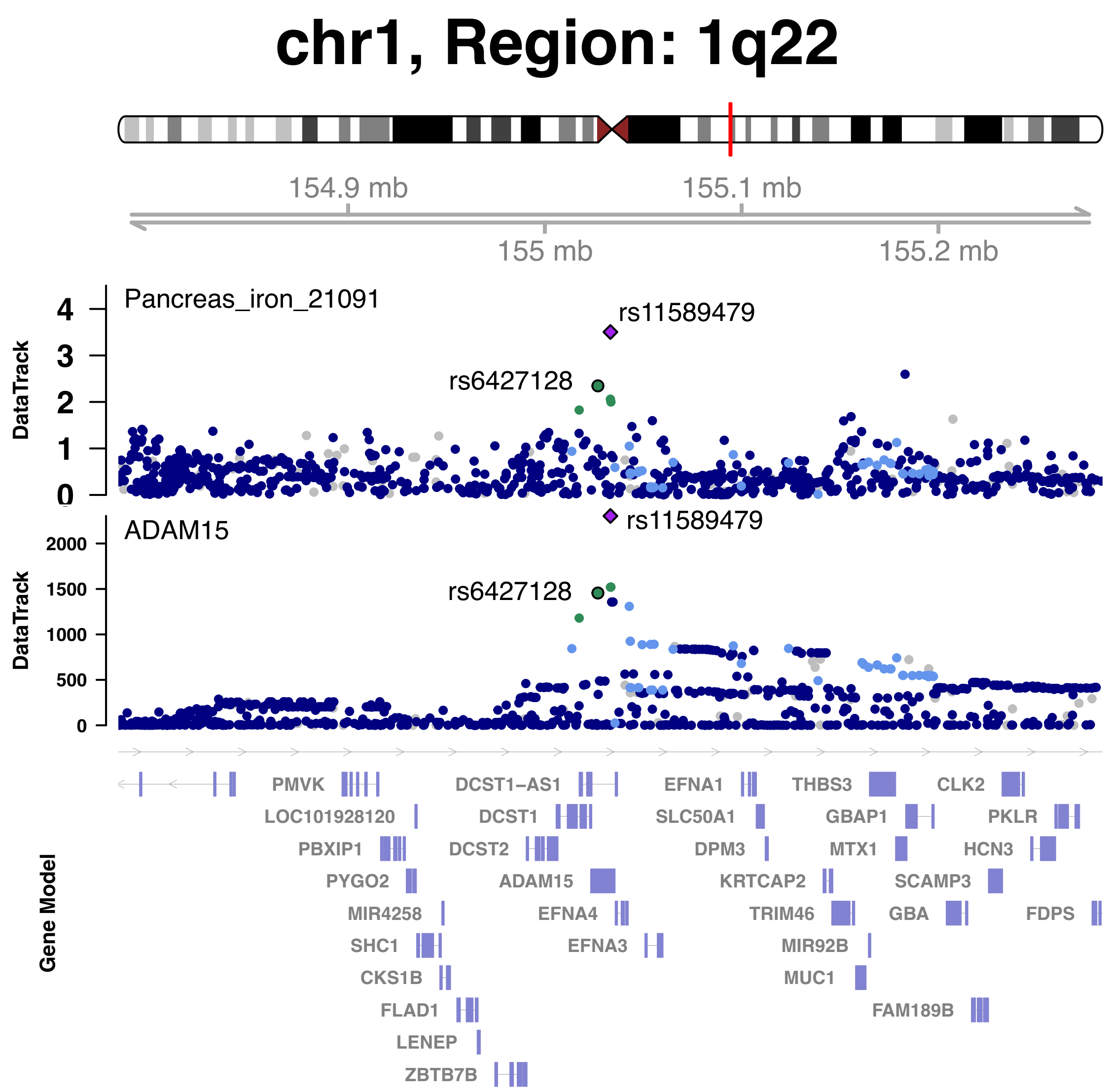


**Fig. S75 Genetic causal pairs between plasma proteins and abdominal MRI identified through Mendelian randomization.**

ADAM15 was causally associated with pancreas iron. The posterior probability of Bayesian colocalization analysis for the shared causal variant hypothesis (PPH4) is 81.22%.
